# How Sex, Age, Adiposity, and Smoking Shape the Human Rib Cage: Evidence from 26,275 Whole-Body MRIs across the German National Cohort (NAKO)

**DOI:** 10.64898/2026.09.01.26361964

**Authors:** Anatol Aicher, Robert Graf, Jan Kirschke, Thomas Frauenfelder, Falko Ensle, Bjoern Menze, Josua Decker, Thomas Kröncke, Johannes Haubold, Steffen Ringhof, Fabian Bamberg, Carsten Oliver Schmidt, Mark Wielpütz, Michael Leitzmann, Stefan N. Willich, Thomas Keil, Thoralf Niendorf, Tobias Pischon, Christopher Schlett, Hendrik Möller

## Abstract

Rib-cage morphology is a determinant of thoracic biomechanics, ventilation, and injury response, yet statistical shape models (SSMs) of the rib cage have relied on small cohorts (∼100s of individuals) imaged by clinical computed tomography, which over-represents injury and disease. We constructed a surface-based SSM of the complete 24-rib cage from 26,275 standardised whole-body magnetic resonance imaging (MRI) scans of adults aged 19–74 years from the population-based German National Cohort (NAKO). Ribs were segmented with a deep-learning pipeline (a rib-extended SPINEPS model), reconstructed as per-rib surface meshes, and brought into dense vertex-wise correspondence by Gaussian-process morphable registration in Scalismo; the aligned ensemble was summarised by generalised Procrustes analysis and principal component analysis (PCA). Fourteen per-rib geometric descriptors provided a quantitative cross- walk between the abstract PCA modes and named shape features, and associations with sex, age, body size and composition (including body-fat percentage), and smoking exposure were estimated by multivariable regression with Benjamini–Hochberg false-discovery-rate control. Shape variation was strongly concentrated: 28 modes captured 95% of the total variance, and the first three alone accounted for 69.4% (PC1, 42.6%; PC2, 16.3%; PC3, 10.5%) and admitted consistent anatomical readings – a sexually dimorphic axis (PC1), a slender-versus-stout body-habitus contrast (PC2), and a free-rib-size axis at ribs 11–12 (PC3). The sexes were nearly fully separated along PC1 (Cohen’s d = 2.52). Body mass and body-fat percentage were the dominant modifiable correlates of rib-cage shape, whereas the association with cumulative smoking exposure was comparatively small. The model is released as a population-representative geometric reference for benchmarking and morphing donor-derived finite-element human-body models and for further large-cohort shape analysis.

## 1. Introduction

Rib cage morphology is an important determinant of thoracic biomechanics and physiology, with implications for trauma, pulmonary medicine, and spinal biomechanics, making it a subject of longstanding interest in anatomy, anthropology, and biomechanics.

In injury biomechanics, rib fractures are the most common skeletal thoracic injury in blunt chest trauma (Liebsch et al., 2019) and a common chest-injury category in nationwide trauma registries (Peek, Ochen, et al., 2020; Peek, Beks, et al., 2020). The finite element human body models that underpin vehicle-safety development (such as THUMS, GHBMC, and related platforms) (Iwamoto et al., 2015; Vavalle et al., 2012) represent rib geometry in detail because shape features govern fracture prediction (S. Holcombe & Huang, 2023; Iraeus et al., 2020), yet that geometry is inherited from a small number of donor anatomies (S. A. Holcombe et al., 2020), motivating ongoing efforts toward parametric, morphable model families (Larsson et al., 2024; Shi et al., 2014; Wang et al., 2016) to better represent a heterogeneous population and their rib cage morphologies (Larsson et al., 2023). In pulmonary medicine, rib cage geometry is a mechanical determinant of ventilation: sex differences in rib inclination are associated with established differences in respiratory function between males and females (Bellemare et al., 2003); chronic obstructive pulmonary disease produces the characteristic “barrel chest” via chronic hyperinflation that remodels the rib cage (Lim et al., 2018; Sverzellati et al., 2013) – in accordance with the classically taught clinical image of barrel chest in smokers; and obesity imposes a restrictive load that scales with central adiposity (Mafort et al., 2016; Wehrmeister et al., 2012). Beyond ventilation, the rib cage is a substantial biomechanical stabiliser of the thoracic spine (Liebsch et al., 2017; Liebsch & Wilke, 2022; Watkins et al., 2005). Taken together, rib cage morphology is associated with various physiological and clinical determinants, and a well-founded model of rib morphology and its variance within a given population is of great interest.

Prior population-scale rib geometry studies have characterised rib geometry in approximately 1,000 trauma-CT subjects through low-dimensional parametric models of the centroidal path, enabling efficient regression against age, sex, height, and weight but constraining the shape representation to a pre-specified parametric family (S. A. Holcombe et al., 2016, 2017). Surface-based statistical shape models (SSMs) of the full rib cage have remained an order of magnitude smaller still (n = 89 (Shi et al., 2014); n = 101 (Wang et al., 2016)), limited by the cost of manual segmentation and establishing dense correspondence. A recent automated-pipeline study reached a substantially larger cohort (n = 1,719) by replacing manual segmentation with a deep-learning whole-body segmentation tool (Robinson et al., 2024), but characterised rib cage shape using ten scalar geometric measurements (bounding-box height, width and depth; convex-hull area in the axial, sagittal and coronal planes; convex-hull volume; and three sagittal angles for rib 1, rib 7 and the sternum) rather than a surface-correspondence shape model, and deliberately restricted the cohort to ages 25–45 years to remove ageing effects. For the latter study, the authors argued for these scalar measurements over principal component analysis on the grounds of interpretability of the results. Notably, all of these lines of work – parametric centerlines, small-cohort surface SSMs, and large-cohort scalar pipelines alike – have drawn almost exclusively on diagnostic computed tomography acquired in clinical settings, producing samples biased toward acute injury, advanced disease, or other clinical indications for chest imaging, and systematically underrepresenting the asymptomatic adult population.

We address these gaps by building an SSM of the full rib cage from standardised whole-body magnetic resonance imaging scans from the German National Cohort (NAKO), a population-based prospective cohort that recruited men and women aged 19–74 years across 18 study centers in Germany (Peters et al., 2022; Rach et al., 2025), with whole-body MRI on 3 T scanners in a sub-cohort of approximately 30,000 participants (Bamberg et al., 2015), of which 26,275 met inclusion and quality-control criteria. Rib segmentations are obtained using an extension of the SPINEPS deep-learning pipeline (Graf et al., 2023; Möller et al., 2024, 2026) retrained to include a rib class on NAKO whole-body MRI, and surface shape modelling is performed in Scalismo, an open-source Scala framework for image analysis and shape modelling that implements Gaussian Process Morphable Models (Luthi et al., 2018). We additionally extract fourteen geometric shape features per rib – thirteen PyRadiomics (Griethuysen et al., 2017) features as well as rib arc length; this provides a quantitative cross-walk between abstract PC modes and named geometric descriptors, recovering scalar interpretability without discarding the coupled multi-region variation that scalar descriptors do not jointly express. This approach combines the data-driven shape representation of an SSM with a cohort size that exceeds the largest prior studies by more than an order of magnitude, reduces the selection bias inherent to clinical CT cohorts, and broadens the metadata space to include body fat percentage and cumulative smoking exposure.

The resulting shape model accordingly serves as a detailed, highly parametrised reference for an adult population of predominantly central European ancestry. We use this model to characterise the principal modes of population-level rib cage shape variation and quantify their associations with sex, age, height, weight, body fat percentage, and smoking (ever-smoker status plus cumulative pack-years).

## 2. Methods

Figure 1 gives a graphical overview of the pipeline applied to one participant. Individual stages are described in the subsections that follow.

**Figure 1.**
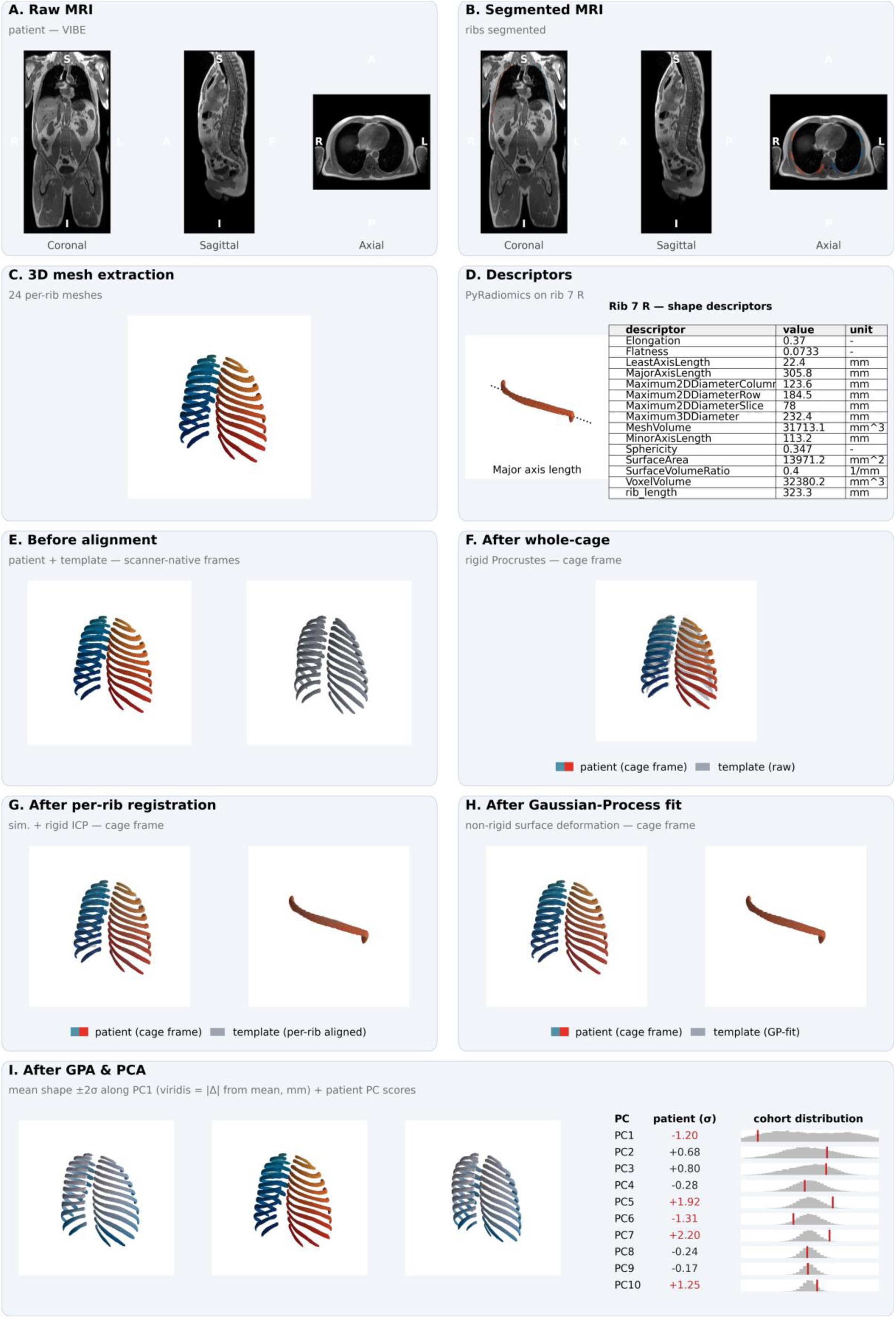
Methodology overview. One display participant walked through every stage of the pipeline. **(A)** Raw VIBE (Volumetric Interpolated Breath-hold Examination) whole-body MRI (coronal, sagittal, axial). **(B)** Rib segmentation overlaid on the same slices, with one colour gradient per side. **(C)** All 24 per-rib surface meshes after marching-cubes extraction, Taubin smoothing, and quadric-error decimation. **(D)** Per-rib PyRadiomics descriptors for the seventh right rib, tabulated alongside the mesh. **(E)** Participant and template ribs shown in their native scanner frames before alignment. **(F)** After rigid whole-cage Procrustes alignment to the template using 24 per-rib centroids. **(G)** After per-rib principal-frame alignment and similarity + rigid iterative-closest-point matching. **(H)** After non-rigid Gaussian-Process surface fitting in four coarse-to-fine passes. **(I)** Generalized Procrustes Analysis: a representative cohort sample (light grey) with the participant (coloured) in shape space. **(J)** Principal component analysis: the PC1 ±2σ surface deformation envelope plus a sparkline table locating the participant on the first ten PCs of the cohort distribution.

### 2.1 Study population and imaging

Whole-body magnetic resonance imaging data and accompanying baseline metadata were drawn from the German National Cohort (NAKO), a population-based prospective cohort of approximately 200,000 men and women aged 19–74 years recruited between 2014 and 2019 across 18 study centers in Germany (Peters et al., 2022; Rach et al., 2025). A sub-cohort of approximately 30,000 participants additionally underwent standardised 3 T whole-body MRI (Bamberg et al., 2015). The present analysis is based on the VIBE MRI sequence acquired within this sub-cohort. Three orthogonal slices of one representative VIBE volume are shown in Figure 1 A. After application of the inclusion and quality-control criteria defined below, the post-ingestion cohort comprised 26,291 participants (14,739 male, 11,552 female; age 48.03 ± 12.27 years, range 19–74 years). Baseline characteristics of the analytic cohort are summarised in Table 1.

**Table 1.**
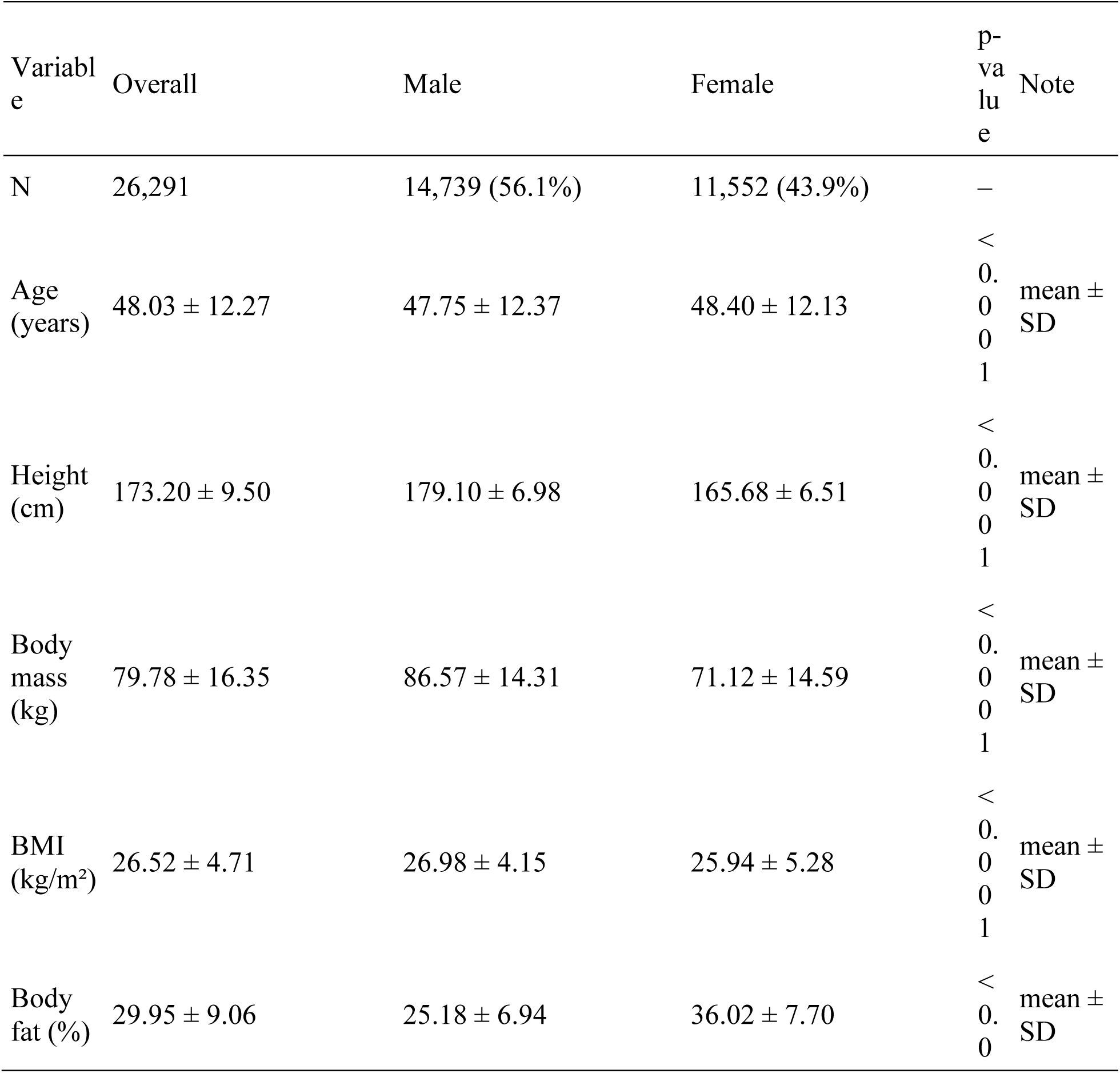
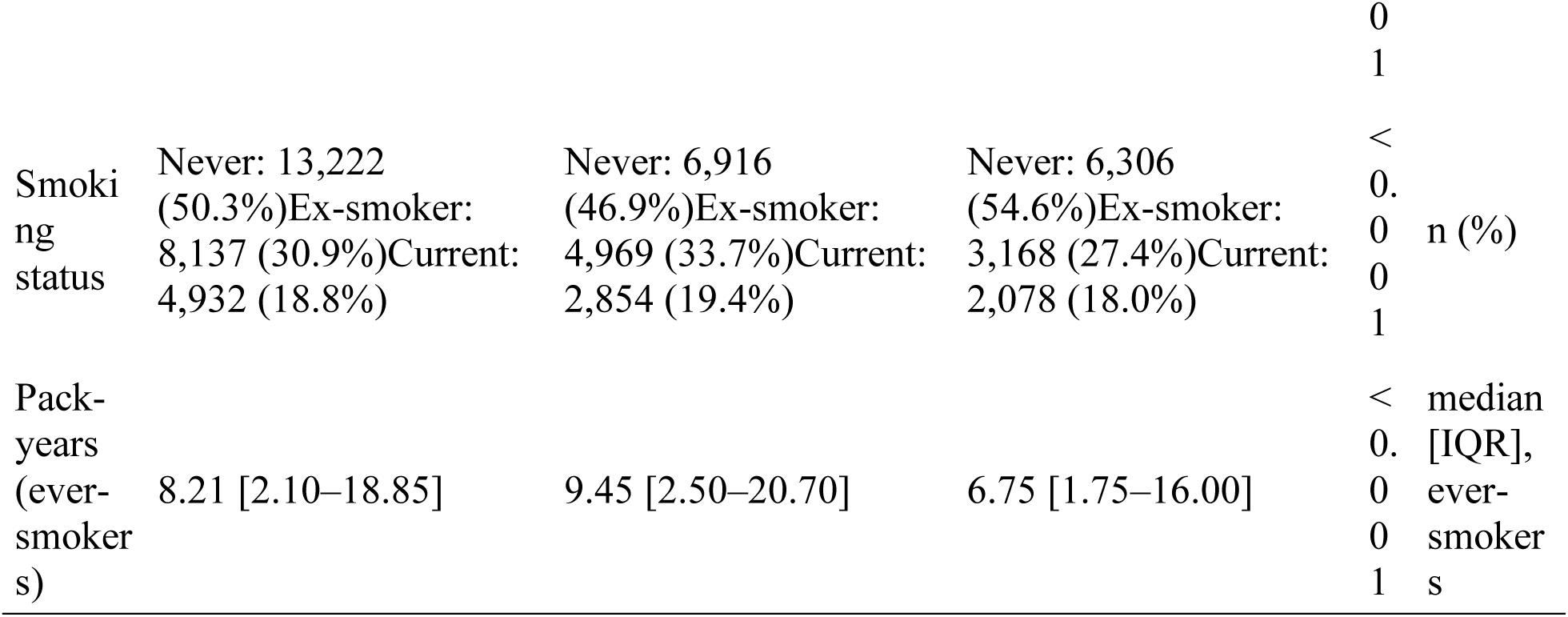
Baseline characteristics of the post-ingestion cohort after exclusions, overall and stratified by biological sex. Continuous variables are summarised as mean ± SD or as median [IQR] for right-skewed distributions. Pack-years are reported for ever-smokers only; never-smokers are coded as zero pack-years (see Cohort definition and quality control). Two-group comparisons use Welch’s t-test for continuous variables and the chi-squared test for categorical variables.

The NAKO is performed with the approval of the relevant local ethics committees and in accordance with national law and with the Declaration of Helsinki of 1975 in its current revised version; written informed consent was obtained from all participants. The present analysis used pseudonymised imaging and baseline data provided under the NAKO data-use and transfer regulations.

### 2.2 Rib segmentation

To achieve rib segmentation annotations, we trained a nnUNet model on 586 manually created VIBE annotations of the NAKO dataset. We reoriented them consistently and left the resolution untouched since it is already consistent across the data. We employed horizontal flipping (x2) and elastic deformation (x3) to increase the data by a factor of 6. We trained on the inphase, outphase, and water sequences of the VIBE images, resulting in roughly 10,000 training images. The model was trained for binary semantic segmentation, i.e., every voxel belonging to a rib has the same class label. We trained with the proposed parameters from nnUNet, other than setting the patch size to a cubic 192 voxels, using the large Residual Encoder Unet architecture (denoted nnUNetResEncUNetLPlans), and their heavy data augmentation strategy denoted DA5. We trained for 1000 epochs.

### 2.3 Per-rib mesh extraction

For each participant the segmentation volume was reoriented to canonical right-anterior-superior (RAS) orientation, and each of the rib labels was extracted. A three-dimensional connected-component analysis on the binary rib mask was used to separate the left and right rib at every level. The two components were assigned to anatomical sides by their centroid along the left–right axis; levels yielding a single component were assigned by comparison against the median per-side centroids of the same participant’s other levels, and levels yielding more than two components were reduced to the two largest. Per-side masks of fewer than 10 foreground voxels were discarded at mesh extraction. All such cases were recorded in a per-participant audit, and participant-level inclusion was decided downstream based on the count of valid rib-side meshes (see Cohort definition and quality control). A triangulated surface mesh was extracted from each binary rib mask by the marching-cubes algorithm at iso-value 0.5, with voxel spacing supplied so that the output vertex coordinates were in millimetres. Voxel-grid stair-stepping was attenuated by 40 iterations of Taubin smoothing (pass-band 0.1), which we used to avoid the volume loss of pure Laplacian filtering, and each mesh was decimated by quadric-error metric to a fixed budget of 1,000 triangular faces. This procedure yielded, for every participant, all individual rib surfaces at a uniform face count, written to STL files. The 24 per-rib meshes for one participant are shown in Figure 1 C.

### 2.4 Per-rib centerline extraction

To measure individual ribs, we utilized the open-source strategy depicted in (Möller et al., 2026). In short, we utilized vertebra spine segmentation to assign individual ribs to vertebrae, in order to obtain the info which rib has a rib on one or either side. Furthermore, an iterative algorithm is employed to derive points along the center of the rib from the start to end. Piece- wise linear interpolation is run afterwards to compute the length of a rib. For more details, see their implementation at https://github.com/Hendrik-code/rib-segmentation.

### 2.5 Cohort definition and quality control

Per-participant phenotype data were taken from the NAKO baseline assessment and comprised age (years), biological sex, standing height (cm), body mass (kg), body mass index, body fat percentage, self-reported smoking status (never, ex-smoker, current) and cumulative cigarette exposure in pack-years. NAKO sentinel codes were recoded as missing, with the exception that pack-years was set to zero for self-reported never-smokers.

Participants were excluded from all downstream analyses if (i) the per-rib audit yielded a non-canonical number of valid rib-side meshes (i.e. other than 24 ribs total); (ii) any rib was flagged as touching the imaging field-of-view boundary or as bifurcated – i.e. detected as branching into two components at the proximal or distal end during the extraction audit; or (iii) any required metadata variable was missing. Counts at each exclusion gate are tabulated in the Supplement.

### 2.6 Per-rib shape descriptors

In parallel to the surface meshes we computed fourteen scalar geometric descriptors per rib using PyRadiomics (Griethuysen et al., 2017), summarised with formal definitions and interpretations in Table 2. Due to its conceptual redundancy with mesh volume, voxel volume was dropped at the analysis stage, leaving thirteen PyRadiomics features. We additionally derived the rib arc length in millimetres from the per-rib centerline coordinates, yielding a final of fourteen descriptors total as shape variables (13 PyRadiomics + rib length). An example descriptor set for rib 7 R is laid out in Figure 1 D. The descriptors furnish a low-dimensional, named representation of each rib that is interpretable independently of the surface- correspondence model; they are used both as direct outcomes in the tabular association analyses and to attach anatomical meaning to the principal modes of the surface shape model in §2.8.4.

**Table 2.**
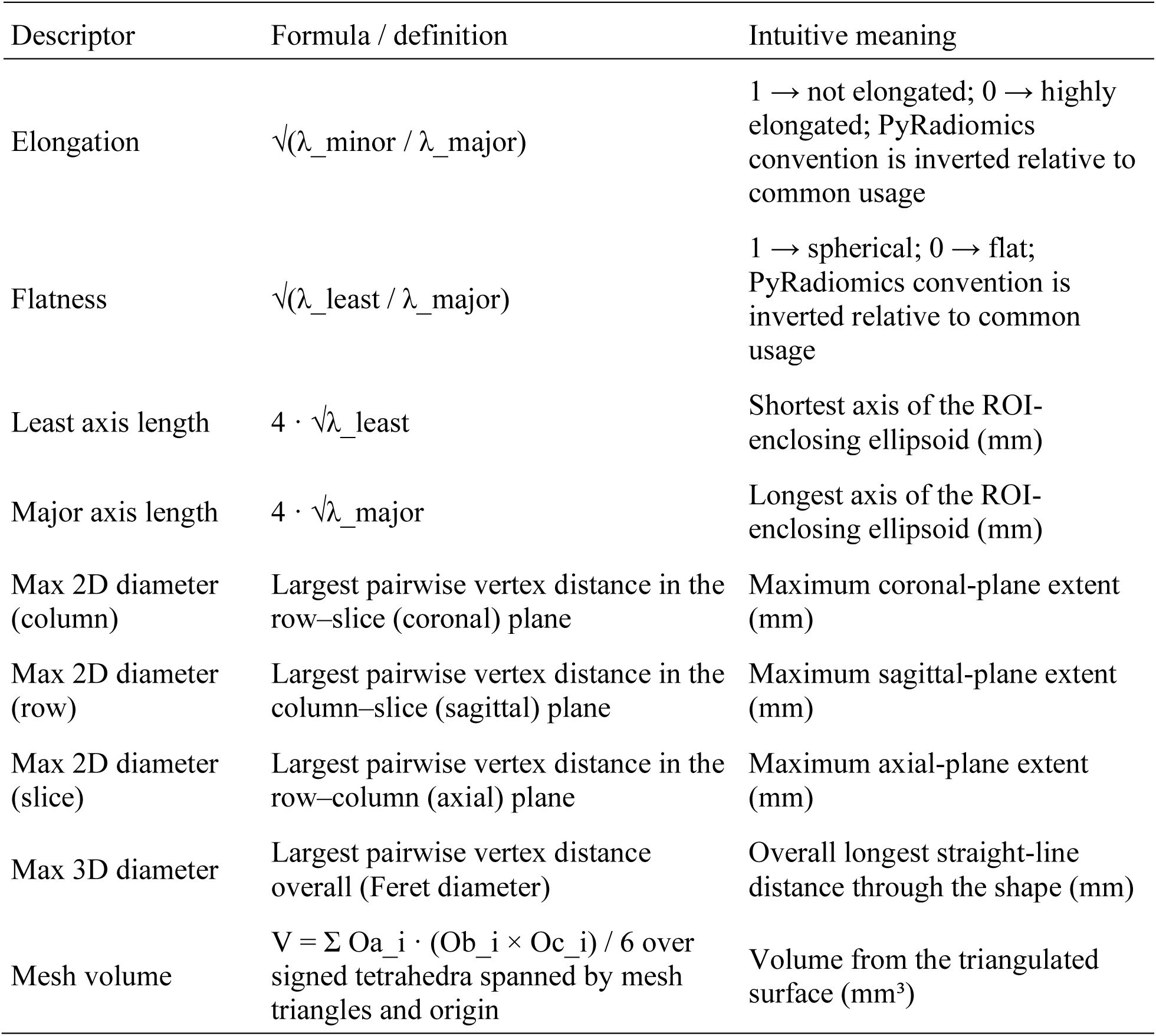
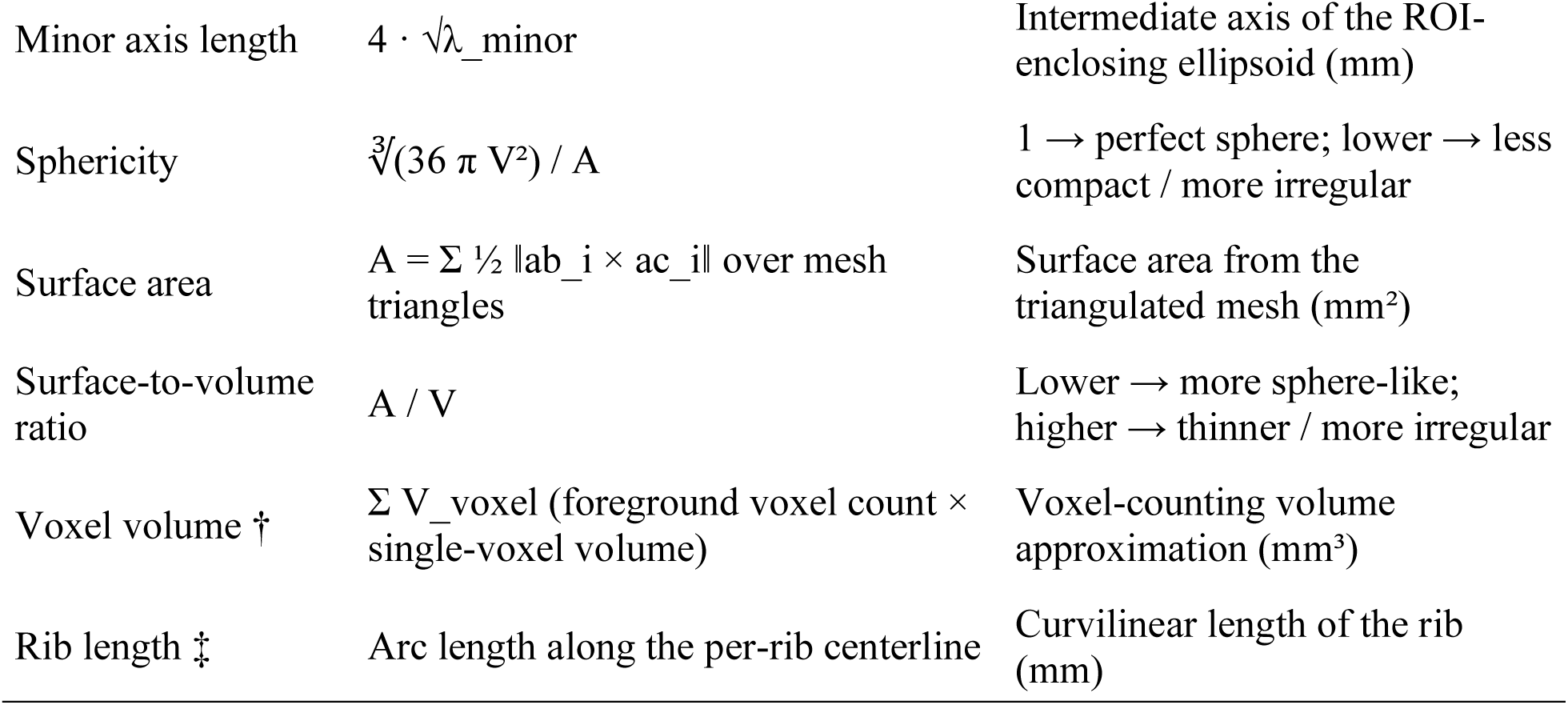
Per-rib shape descriptors used in the analyses. Formal definitions are paraphrased from the PyRadiomics documentation (https://pyradiomics.readthedocs.io/en/latest/features.html, accessed 2026-05-28); the software is described by Griethuysen et al. (2017). λ_major ≥ λ_minor ≥ λ_least denote the eigenvalues of the ROI’s principal axes obtained by principal-component decomposition. † Extracted but dropped at the analysis stage due to its conceptual redundancy. ‡ Project-derived from the per-rib centerline; not produced by PyRadiomics.

### 2.7 Statistical shape model

#### 2.7.1 Template selection

A single participant was designated as the registration template by a centrality-based procedure. From the cohort of participants with a complete standard set of 24 per-rib meshes we drew a random subsample of *n* = 500 participants S. For each rib identity *r* ∈ ℛ (the 12 levels × 2 sides) and every pair of subsample participants *p*, *q* ∈ S we computed the symmetric mean closest-point surface distance between their meshes *M*: *d*(*M_p_*_,*r*_, *M_q_*_,*r*_). The per-rib centrality score of participants *p* at rib identity *r* is the mean distance from that rib to all other subsample ribs of the same identity,

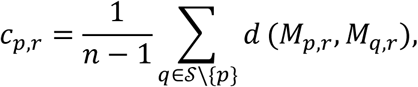

and the aggregate centrality score *C_p_* = ∑*_r_*_∈ℛ_ *c_p_*_,*r*_ sums these per-rib scores across the 24 rib identities. The participant minimising *C_p_*, i.e. the one whose 24 ribs lay, on average across the full thoracic cage, closest to all other subsample participants, was returned as the candidate template.

The candidate template was then rendered as an interactive three-dimensional viewer of its 24 per-rib meshes and visually inspected by A.A. and H.M. for anatomical typicality and segmentation quality. The first candidate returned by the procedure exhibited a near-bifid morphology for one rib and the procedure was therefore re-run with a different random seed; the resulting second candidate, with anatomically unremarkable morphology across all 24 ribs and no visible segmentation artefacts, was adopted as the template. The chosen identifier is recorded with the model and reused in all downstream registrations.

#### 2.7.2 Mesh-to-template registration

Dense vertex-wise correspondence between every rib of every participant and the corresponding rib of the template was established by a multi-stage registration pipeline built on the Scalismo framework (Luthi et al., 2018). The pipeline proceeds in three stages, illustrated in Figure 1 F–H.

First, a rigid Procrustes alignment of the whole rib cage to the template was performed using the centroid of each of the 24 ribs as a correspondence point; a bilateral-symmetry safeguard reset the rotation to the identity whenever its trace fell below 1 – equivalent to a rotation angle exceeding 90°. Second, each rib’s template counterpart was independently aligned to the target rib by matching principal frames: the template’s two in-plane principal axes were rotated onto the target’s, with each target axis’s sign disambiguated by requiring a positive dot product with the corresponding template axis (well-defined given the prior whole-cage alignment, and constructed so that anatomical orientation is preserved by construction rather than chosen post hoc); the third axis was set as the cross product of the first two to guarantee a right- handed frame. This pose alignment was followed by 30 iterations of similarity (rigid plus uniform scale) iterative closest-point matching with an 80%-trimmed mean criterion and 25 iterations of rigid-only ICP with an 85%-trimmed mean criterion, each with a closed-form per-iteration update – Umeyama’s algorithm (Umeyama, 1991) for the similarity case and orthogonal Procrustes (Schönemann, 1966) for the rigid case. Third, non-rigid registration was performed by Gaussian Process surface deformation in four coarse-to-fine passes *π* = 1, …,4 with shrinking outer length scales; at each pass the kernel was rebuilt and proposed correspondences (*s*, *t*) – where *s* is a sampled vertex of the current deformed reference and *t* its closest point on the target surface – were retained for the posterior update only when all three of the following held: a closest-point distance threshold 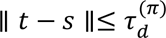, exclusion of boundary points on the target, and a surface-normal agreement criterion **n***_s_* ⋅ 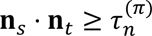, where **n** and **n***_t_* are the unit surface normals at *s* and *t*. Correspondences were generated in the source-to-target direction. The pass- dependent schedule – ICP iterations, observation noise variance, distance and normal thresholds, and the number of sampled reference vertices – is given in Table 3. Finally, the registered template surface was pushed back through the per-rib similarity transform and the inverse whole-cage Procrustes transform so that every output mesh was returned to the participant’s native scanner coordinate frame while preserving template-derived vertex correspondence.

**Table 3.** Coarse-to-fine non-rigid registration schedule. D denotes the bounding-box diagonal (mm) of the per-rib template mesh. “Vertex samples” is the number of source vertices uniformly sampled from the reference mesh that anchor the GP posterior at each ICP iteration of the pass. Across passes the Gaussian-process outer length-scale additionally shrinks by a factor of 0.8 per pass relative to the pass-1 value (initialised at max(D/4, 30) mm); a fixed inner kernel of 25 mm is summed with it to capture local thickness variation.

| Pass $\pi$ | ICP iters | Noise var.<br>(mm<sup>2</sup>) | $\tau_d^{(\pi)}$ (mm) | Vertex<br>samples | $\tau_n^{(\pi)}$ |
| --- | --- | --- | --- | --- | --- |
| 1 | 15 | 5.00 | $\max(0.10 D, 30)$ | 1,500 | 0.30 |
| 2 | 20 | 1.00 | $\max(0.05 D, 15)$ | 2,500 | 0.50 |
| 3 | 25 | 0.25 | $\max(0.02 D, 8)$ | 4,000 | 0.60 |
| 4 | 30 | 0.05 | $\max(0.01 D, 4)$ | 6,000 | 0.70 |

#### 2.7.3 Generalized Procrustes Analysis and principal component analysis

Registered meshes were stacked across participants in a canonical anatomical order (rib 1 left, rib 1 right, …, rib 12 right) into a three-dimensional tensor of shape *N* × *P* × 3, where *N* is the number of included participants and *P* is the total number of correspondence points across the 24 ribs. The ensemble was aligned by Generalized Procrustes Analysis (GPA): each shape was iteratively rotated and translated towards the running ensemble mean using a singular-value-decomposition solution corrected for reflections (Gower, 1975). Scale was retained, because overall body size is among the explanatory variables of interest. Figure 1 I shows the GPA-aligned ensemble with one participant superposed.

Principal component analysis was applied to the GPA-aligned, mean-centered shape ensemble via a randomised singular value decomposition. The number of retained modes was determined by retaining the smallest number of modes such that cumulative explained variance reached 95%, yielding *K* = 28 principal modes. Per-participant PC scores were computed by projecting each shape onto the retained basis. Figure 1 J shows the PC1 ±2σ surface deformation and the participant’s per-PC score table.

### 2.8 Association analyses

We related rib-cage shape to participant metadata through three complementary regression estimands – unadjusted, adjusted, and targeted – each applied to two outcome families: the fourteen per-rib shape descriptors of Methods §2.6 and the *K* principal-component scores of the surface shape model. Descriptor models were fit at the rib level on within-rib *z*- standardised outcomes – each descriptor centered and scaled to mean 0 and unit standard deviation within each rib position across participants, which absorbs the rib-position level term – with standard errors clustered at the participant level (HC1-style small-sample correction) to account for the within-participant correlation of the 24 rib-sides. Principal-component models treated each participant’s score on a mode as a single outcome and used HC3 heteroscedasticity-robust standard errors. Throughout, sex was encoded as a binary is_female indicator (male reference) and smoking as an ever_smoker indicator together with cumulative pack-years; the three-level smoking-status variable was retained only as the source for ever_smoker. The three estimands are defined below. The targeted causal models (§2.8.3) are our primary specification; the unadjusted and adjusted models are reported alongside them for reference.

#### 2.8.1 Unadjusted (marginal) associations

Each metadata variable was related to each outcome one predictor at a time, capturing the total marginal association. Predictors comprised sex, age, standing height, body mass, body mass index, body fat percentage, ever-smoker status, and pack-years; body mass index enters at this layer only (it is redundant with height and body mass once both are in a model), and pack-years was fit on ever-smokers alone. For every (predictor, outcome) pair we report the raw and standardised slope, the marginal *R*^2^, and the *p*-value; for the binary sex contrast we additionally report Cohen’s *d* as a standardised effect size.

#### 2.8.2 Adjusted (multivariable) associations

We fit a single multivariable model on the full analytic cohort entering all seven core predictors simultaneously – sex, age, standing height, body mass, body fat percentage, ever-smoker status, and pack-years. Body mass index was excluded as redundant with height and body mass; never-smokers enter as ever-smoker = 0, pack-years = 0, so the never/ever contrast and the within-smoker dose are carried in one model. For every (predictor, outcome) pair we report the raw and standardised slope and a Frisch–Waugh–Lovell partial *R*^2^ quantifying the unique variance share attributable to that predictor net of the rest of the model. Because every coefficient is mutually adjusted, these are associations net of the measured covariates. We note that these are adjusted associations rather than the causal effect of any single predictor (see the “Table 2 fallacy” (Westreich & Greenland, 2013)). The targeted models below address this.

#### 2.8.3 Targeted causal models

Our primary analysis estimates one causal contrast per exposure under an explicit directed acyclic graph (DAG) of the assumed relationships among the metadata variables and rib-cage shape (Figure 2). For each exposure we fit a separate model that conditions only on that exposure’s minimal back-door adjustment set, so that each coefficient targets a defined estimand rather than a mutually-adjusted partial association. Sex and age are treated as exogenous, so their effects are estimated as total effects, each adjusted only for the other; height, body fat percentage, ever-smoker status, and pack-years are taken to be influenced by sex and age, so their adjusted associations condition on sex and age; body mass lies downstream of body size and composition, so its adjusted association additionally conditions on height and body fat percentage. Each model was fit to both outcome families under the same standard-error scheme as above (participant-clustered for descriptors, HC3 for principal-component scores), and reported with the raw and standardised slope and the Frisch–Waugh–Lovell partial *R*^2^. The targeted estimates are reported alongside the unadjusted (total marginal) and adjusted (mutually-adjusted) coefficients, so that the movement of an effect across the three specifications exposes its confounding and mediation structure.

**Figure 2.**
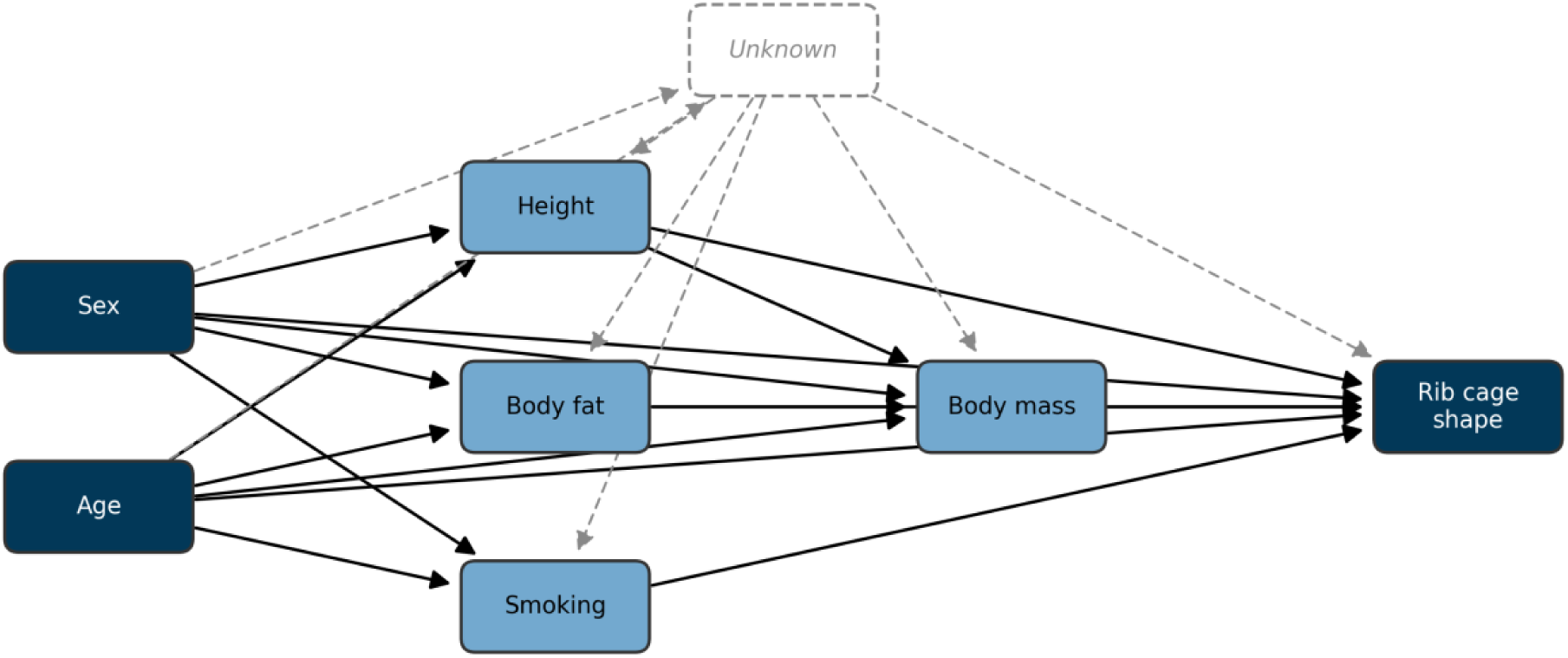
Assumed causal structure for the targeted analysis. Sex and age are exogenous; height, body fat percentage and smoking are children of sex and age; body mass is additionally downstream of height and body fat; every exposure acts on rib-cage shape. Each targeted estimate conditions only on the exposure’s back-door set. Dashed edges mark an unmeasured confounder (U), itself influenced by sex and age, acting on the non-exogenous exposures and on shape: the total effects of sex and age remain identified (U lies on a path descending from them), whereas the body- composition and smoking associations are not point-identified under such confounding.

The identification this affords is asymmetric. Because sex and age are exogenous, their total effects remain valid even if unmeasured factors influence the downstream metadata variables, as such a factor would lie on a causal path descending from sex and age rather than on a back-door path. The adjusted associations of the body-composition and smoking exposures, by contrast, assume no unmeasured confounding with rib-cage shape: any unmeasured common cause of, for example, body composition and thoracic shape would open a back-door path that the measured adjustment sets cannot close, so these coefficients should be read as adjusted associations rather than causal effects.

#### 2.8.4 Shape-mode interpretation via radiomics descriptors

To attach anatomical meaning to the principal modes of the surface shape model we computed, for every triplet of (PC, rib position, per-rib shape descriptor), a univariate OLS model in which the cohort-z-scored PC score predicted the descriptor at that rib position, and report both native-unit and standardised slopes. Effects were summarised as per-PC heatmaps (24 rib positions × fourteen descriptors) and per-descriptor heatmaps (24 rib positions × *K* PCs).

This analysis provides a quantitative cross-walk between the abstract shape-model basis and named geometric descriptors at every rib.

#### 2.8.5 Multiple-comparison correction

Each model layer was corrected separately for multiple comparisons by the Benjamini– Hochberg false discovery rate procedure (Benjamini & Hochberg, 1995) at a nominal *q*-value threshold of 0.05: the unadjusted, adjusted, and targeted families on each of the descriptor and principal-component sides, together with the PC × rib × descriptor cross-walk. Correction was applied within each family separately so that adjusted *p*-values remain interpretable within their grouping.

#### 2.8.6 Geometry prediction model

To turn these associations into a generative tool, we additionally fit a model that maps demographics directly to rib-cage geometry. For each retained mode the per-participant PC score was regressed on the seven core predictors (continuous predictors centered at their cohort means) by ordinary least squares, and the resulting coefficient matrix, intercepts, and predictor means were stored as a compact generator bundled with the PCA basis. Given any covariate profile the generator predicts the leading PC scores and reconstructs a full surface as the cohort-mean shape plus those scores projected back through the PCA modes; an interactive version of the generator is available online (SSM Viewer). Predictive fidelity was assessed by 10-fold cross-validation over participants: holding out each fold in turn, we fit the generator on the remaining participants, predicted the held-out participants’ PC scores from their covariates alone, reconstructed each surface, and measured its per-vertex distance to that participant’s GPA-aligned shape. We report the mean and 95th-percentile of this demographics-only prediction error (mm), overall and by sex, as a function of the number of reconstructed modes. The error quantifies how much of rib-cage geometry is recoverable from demographics alone; being a prediction measure it carries no false-discovery correction, and it is distinct from the Styner triad (§2.9), which characterises the PCA basis rather than its predictability from demographics.

### 2.9 Shape-model quality assessment

The fitted shape model was characterised by the three classical SSM quality metrics following Styner et al. (Styner et al., 2003). *Compactness* is the cumulative explained variance as a function of the number of retained modes. *Generalisation* was estimated by 100-fold cross-validation: the cohort was randomly partitioned into 100 disjoint folds and, for each fold, a PCA was refit on the remaining shapes and the held-out shapes projected onto the first *k* modes and reconstructed; we report the resulting per-vertex root-mean-square reconstruction error averaged over folds, as a function of *k*. *Specificity* was estimated by drawing 200 random samples from the Gaussian shape distribution defined by the fitted model and reporting, as a function of *k*, the mean per-vertex distance from each sample to its nearest training-set neighbour. Each metric was computed both on the whole-cage shape vector and on each rib stack separately, allowing model performance to be localised.

### 2.10 Software and reproducibility

The complete pipeline – mesh extraction, descriptor computation, surface registration, statistical shape modelling, association analyses, and figure generation – is implemented in Python (Version 3.13, Python Software Foundation) and Scala (Version 3.3.1, EPFL). Python stages rely on NumPy (Version 2.4.4) (Harris et al., 2020), pandas (Version 3.0.2) (McKinney, 2010), scikit-learn (Version 1.8.0) (Pedregosa et al., 2011), statsmodels (Version 0.14.6) (Seabold & Perktold, 2010), SciPy (Version 1.16.3) (Virtanen et al., 2020), PyVista (Version 0.47.1) (Sullivan & Kaszynski, 2019), Plotly (Version 6.7.0, Plotly Technologies Inc.), and Altair (Version 6.1.0) (Satyanarayan et al., 2017; VanderPlas et al., 2018); the registration stage is implemented on top of Scalismo (Version 1.0-RC1) (Luthi et al., 2018) running on JDK 17 (Oracle Corporation). Full source code, configuration for the canonical run, and the figure bundle reproduced in this manuscript are available at https://github.com/AnatolAicher/NAKO-Ribcage-SSM. An interactive version of this manuscript is available at https://anatolaicher.github.io/NAKO-Ribcage-Manuscript/.

## 3. Results

**3.1 Cohort, segmentation yield, exclusions**

Of the 30,218 participants in whom both a complete VIBE rib-segmentation output and the full set of baseline metadata variables were available, 26,275 met all quality-control and registration requirements and entered the analytic cohort used in the SSM-based analyses that follow (Table 4).

**Table 4.** Exclusion flow for the analytic cohort. Note that exclusions are not mutually exclusive: a participant may fail multiple gates, and the counts at each gate reflect the total number of participants failing that gate regardless of other failures. The final analytic cohort is the subset of participants passing all gates.

| Step | n | % |
| --- | --- | --- |
| Joined cohort | 30,218 | 100.0% |
| Excluded – rib-side count outside 24 | 2,716 | 9.0% |
| Excluded – missing baseline metadata | 1,350 | 4.5% |
| Excluded – segmentation touching FOV boundary | 1 | <0.1% |
| Excluded – bifurcated proximal/distal rib end | 0 | 0.0% |
| Post-ingestion cohort | 26,291 | 87.0% |
| Excluded – incomplete or failed surface registration | 16 | <0.1% |
| Analytic cohort | 26,275 | 87.0% |

Ingestion-stage exclusions were dominated by missing baseline metadata and by participants with a rib-count outside of 24. A small residual of participants with sufficient rib coverage failed surface registration. 27,502 participants (91.0% of the joined participants) yielded the full complement of 24 rib sides; the remainder distribute approximately symmetrically around this mode (full distribution: Supplement S1.1).

Baseline demographic and anthropometric characteristics are reported on the post-ingestion cohort (n = 26,291) in Table 1 (§2.1). The analytic cohort spans the full adult age range (19–74 years), with a slight male predominance.

Thus, the cohort exceeds the largest prior surface-correspondence SSM of the human rib cage by approximately two orders of magnitude, and is distinguished from prior shape model cohorts assembled from clinical CT (Robinson et al., 2024; Shi et al., 2014; Wang et al., 2016) and from the trauma-CT centerline study of S. A. Holcombe et al. (2016) in terms of population representativeness.

### 3.2 Principal modes of shape variation

The GPA-aligned cohort-mean rib cage is shown from three orthogonal viewpoints in Figure 3 and provides the anatomical reference frame against which all subsequent shape variation is described. All principal-component scores reported in the remainder of the Results section quantify per-participant deviations from this mean shape.

**Figure 3.**
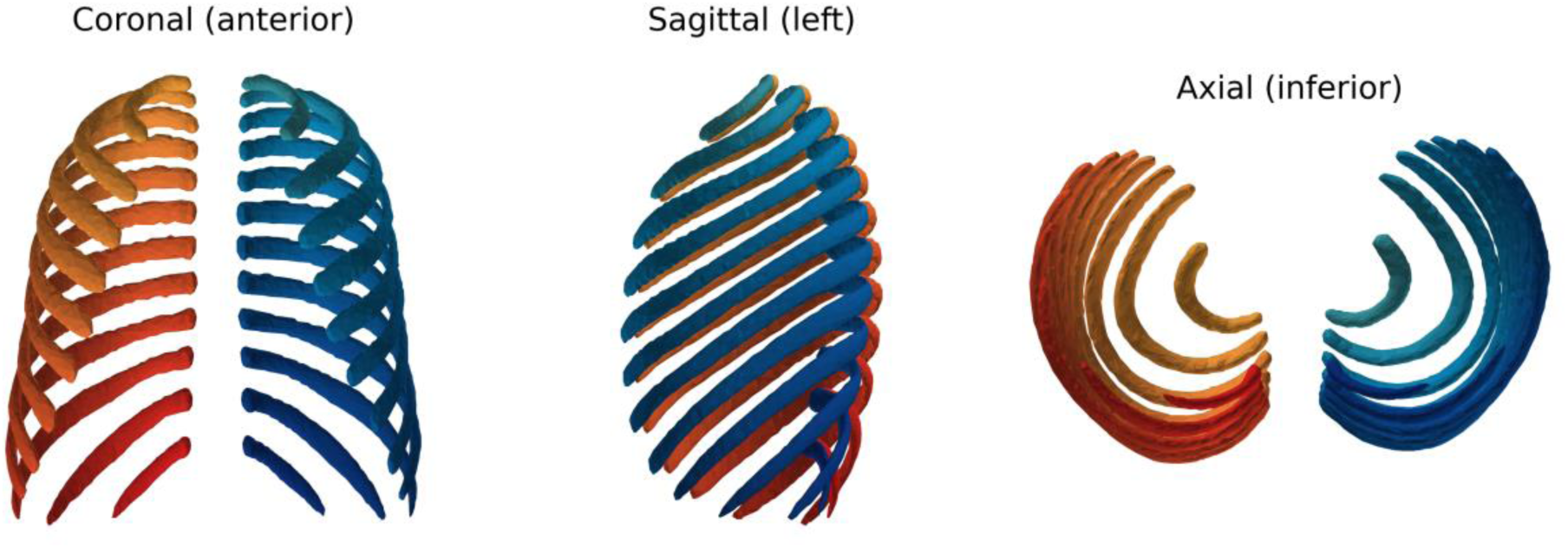
Cohort-mean rib cage. Coronal (anterior view), sagittal (right view), and axial (superior view) renderings of the GPA-aligned mean shape; left ribs are shaded blue, right ribs red. The mean is the reference shape from which the principal-component deformations of Figure 4 are computed.

Retaining the smallest number of modes such that cumulative explained variance reached 95% yielded 28 modes; the leading three modes together account for 69.4% of the total shape variance (PC1: 42.6%; PC2: 16.3%; PC3: 10.5%), with no subsequent mode individually exceeding 5.4%. The corresponding ±2σ surface deformations of the leading modes are shown in Figure 4.

**Figure 4.**
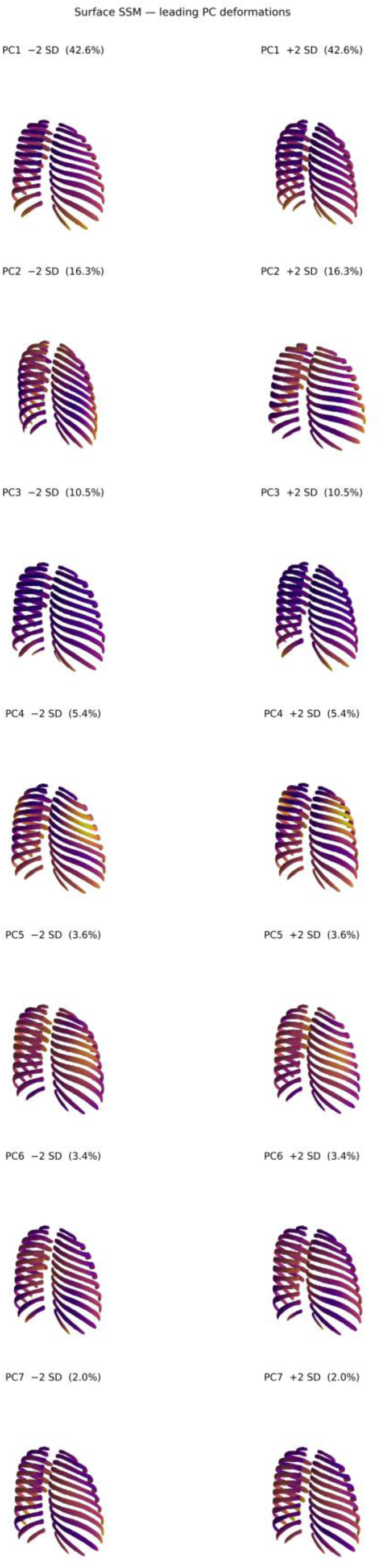
Leading principal-component modes. ±2σ surface deformations of the top modes of cohort rib- cage shape variation, expressed as displacements from the mean shape (Figure 3). An interactive viewer with continuous per-PC sliders and metadata controls is available on the SSM Viewer page.

### 3.3 Shape-model fidelity

The fitted SSM was characterised by the three standard quality metrics of Styner et al. (2003) – compactness (cumulative explained variance), generalisation (held-out reconstruction error), and specificity (mean distance from samples drawn from the fitted Gaussian shape distribution to their nearest training-set neighbour) – evaluated for k = 1 to 25 modes on both the whole-cage shape vector and each of the 24 per-rib subspaces (Figure 5).

**Figure 5.**
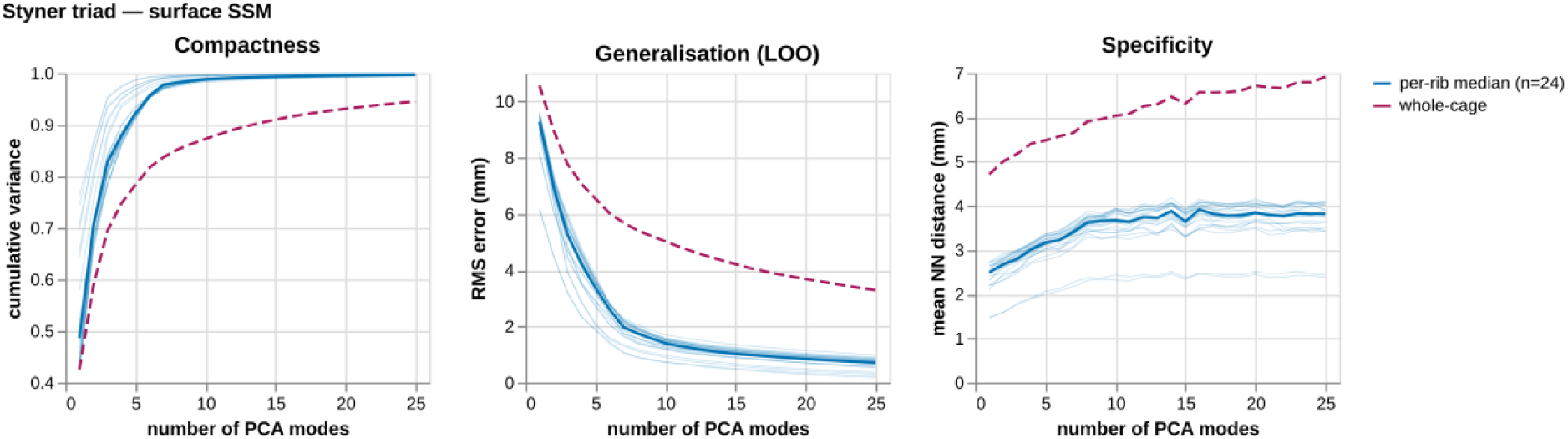
Styner triad of shape-model quality. Compactness, generalisation (100-fold cross-validation), and specificity (200 random samples from the fitted Gaussian shape distribution) as functions of the number of retained PCA modes. Faint blue lines: each of the 24 per-rib subspaces; solid blue: per-rib median; dashed red: whole-cage shape vector.

At k = 25 modes – the upper bound of the evaluation range – the whole-cage SSM achieves a compactness of 94.5%, a generalisation error of 3.29 mm per vertex, and a specificity of 6.92 mm per vertex. The per-rib subspaces reach comparable compactness and reconstruction error in substantially fewer modes than the joint 24-rib shape vector (Figure 5), reflecting their lower intrinsic dimensionality.

Separately, we quantified how much of an individual’s rib-cage geometry is recoverable from demographics alone, using the held-out geometry-prediction model (Methods §2.8.6). Demographics predicted held-out shape to a mean per-vertex error of 8.82 mm (95th percentile 12.90 mm), against a 3.29 mm basis reconstruction error at k = 25. The held-out error changed little with the number of reconstructed modes (8.93 mm at three modes; 8.82 mm at K), and demographic predictive *R*^2^ was concentrated in PC1 (0.78) and PC2 (0.62), below 0.10 thereafter. This means that while demographics explain a substantial portion of the total shape variance (approximately 45%), this is concentrated in the two leading body-size modes, and the remaining modes of shape variation are largely unpredictable from demographics alone. A pure demographics-based generator thus produces an “average” shape for the given covariate profile. To capture the whole shape distribution, including the unpredictable modes, the generator can be supplemented with random sampling from the residual shape distribution (the Gaussian defined by the PCA basis and eigenvalues) to produce shapes that are both demographically typical and realistically variable.

### 3.4 Descriptor-level associations

Each of the 14 per-rib shape descriptors (Methods §2.6) was related to the seven exposures under the three estimands of Methods §2.8, with the targeted causal models as the primary specification. Of the 98 targeted (exposure, descriptor) pairs, 81 reached Benjamini– Hochberg FDR significance (q < 0.05); the largest targeted partial R² values were mesh volume × sex (female) (partial R² = 0.351, β = −0.59); surface area × sex (female) (partial R² = 0.348, β = −0.59); rib length × sex (female) (partial R² = 0.297, β = −0.54). The targeted standardised slopes (Panel A) and Frisch–Waugh–Lovell partial R² (Panel B) are mapped in Figure 6; the adjusted and unadjusted descriptor maps are in the Supplement.

**Figure 6.**
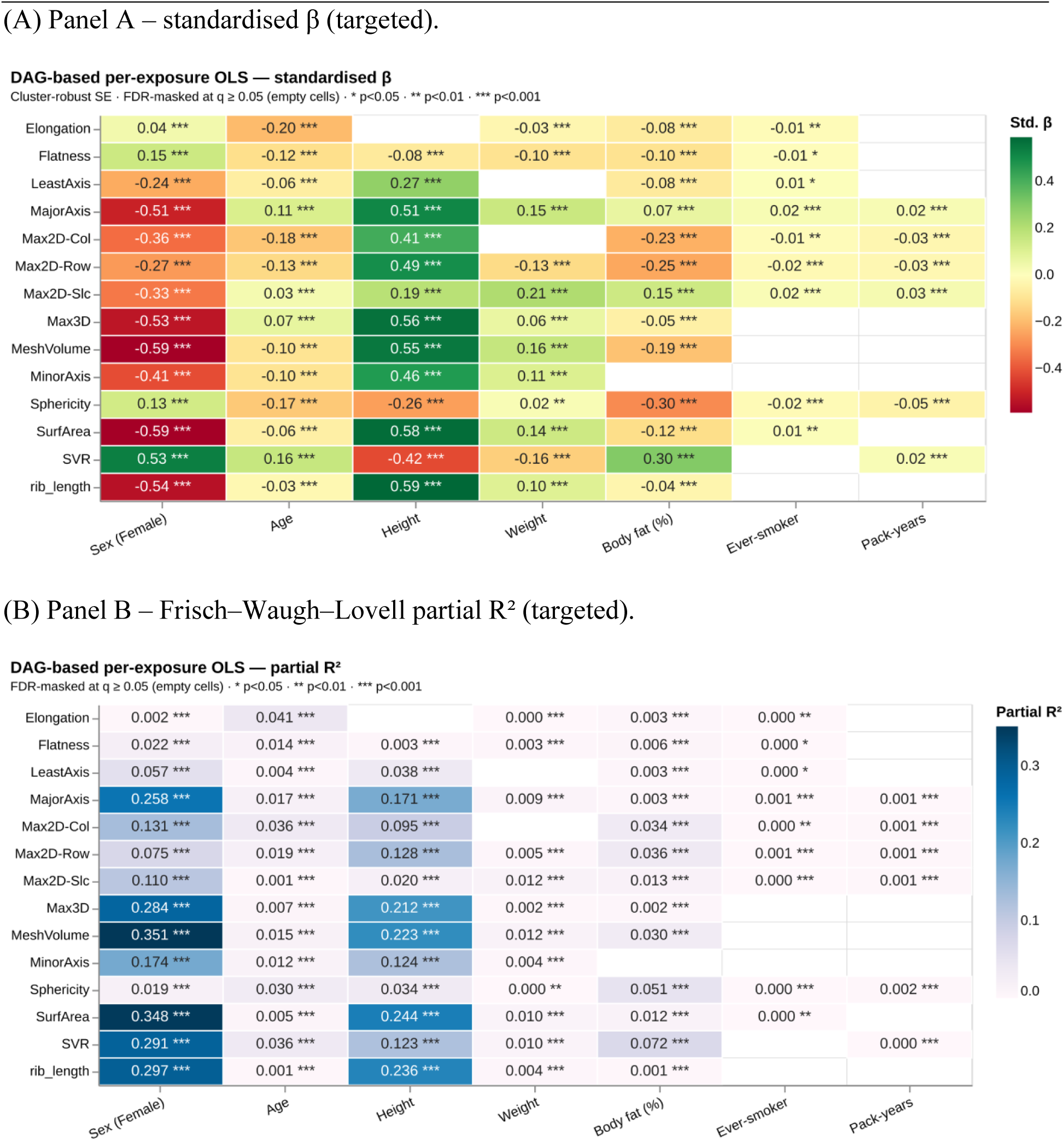
Targeted causal (DAG) descriptor effect maps. **(A)** Standardised β per (shape descriptor × exposure) under each exposure’s targeted estimand – a total effect for sex and age, an adjusted association under the back-door set for the other five exposures (Methods §2.8.3). **(B)** Frisch– Waugh–Lovell partial R² for the same pairs. Both panels are FDR-masked at q ≥ 0.05 (empty cells). Cluster-robust standard errors at the participant level. The adjusted and unadjusted equivalents and the standardised-β forest plot are in the Supplement.

Overall, sex and height are the strongest shape predictors, both in terms of effect size and determination; smoking is the weakest.

The complementary principal-component view, where covariate signal concentrates into a few modes, follows in §3.5.

### 3.5 PC-score associations

Per-participant PC scores were regressed on the metadata under the three estimands of Methods §2.8, with the targeted causal models as the primary specification; the targeted standardised β across PC1–PC7 are mapped in Figure 7. PC1 is the most strongly explained mode (model R² = 0.78) and PC2 the next (R² = 0.62), with R² below 0.10 from PC3 onward. Sex strongly separated the cohort along PC1 (Cohen’s d = 2.52, female − male; d = 0.37 for PC2), whereas ever-smoker status reaches FDR significance on 21 of the 28 modes, all at small standardised effect sizes.

**Figure 7.**
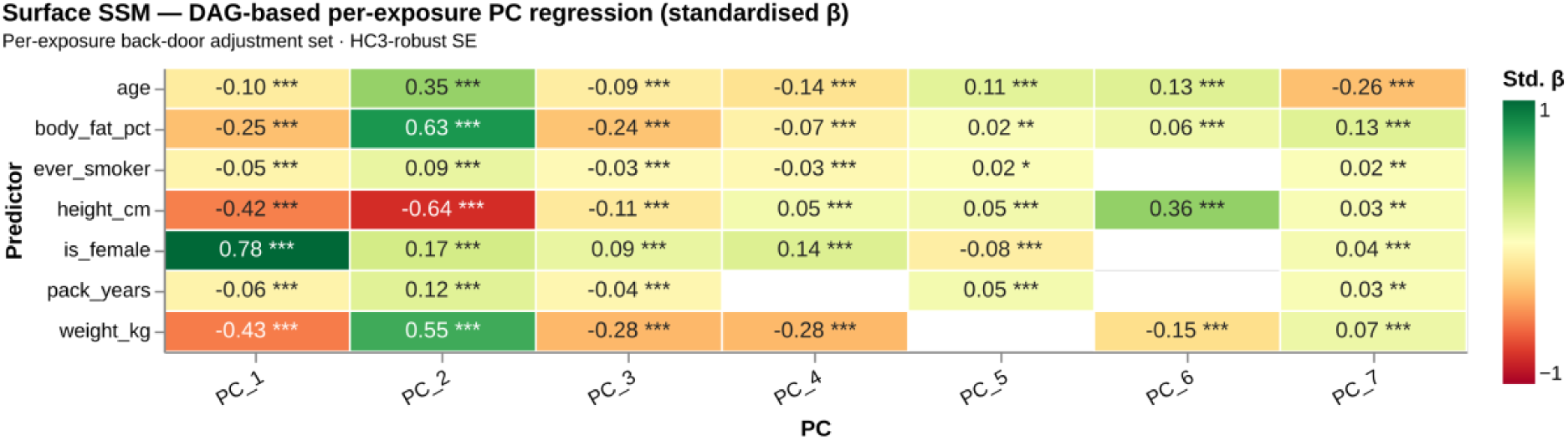
Targeted causal (DAG) associations with the leading principal-component scores. Standardised β from the per-PC targeted models against the seven exposures (rows) – a total effect for sex and age, an adjusted association for the other five (Methods §2.8.3) – for PC1– PC7 (columns). Cells are FDR-masked at q ≥ 0.05 (empty). The adjusted β map, the β-vector field, and the per-rib PC loadings are in the Supplement.

Table 5, Table 6 and Table 7 set the targeted coefficient for each exposure beside its unadjusted and adjusted counterparts on PC1 and PC2.

**Table 5.** Standardised β of each exposure on PC1 under the three estimands. The targeted (DAG) estimate (bold) is the primary specification: a total effect for sex and age, an adjusted association under each remaining exposure’s back-door set.

| Predictor | Unadjusted $\beta$ | Adjusted $\beta$ | Targeted $\beta$ (DAG) | Estimand |
| --- | --- | --- | --- | --- |
| sex (female) | +0.78 | +0.34 | **+0.78** | Total effect |
| age | -0.07 | -0.13 | **-0.10** | Total effect |
| height | -0.73 | -0.23 | **-0.42** | Adjusted assoc. |
| body mass | -0.69 | -0.43 | **-0.43** | Adjusted assoc. |
| body fat (%) | +0.29 | +0.13 | **-0.25** | Adjusted assoc. |
| ever-smoker | -0.12 | -0.02 | **-0.05** | Adjusted assoc. |
| pack-years | -0.17 | -0.03 | **-0.06** | Adjusted assoc. |

**Table 6.** Standardised β of each exposure on PC2 under the three estimands (targeted estimate in bold).

| Predictor | Unadjusted $\beta$ | Adjusted $\beta$ | Targeted $\beta$ (DAG) | Estimand |
| --- | --- | --- | --- | --- |
| sex (female) | +0.18 | -0.23 | **+0.17** | Total effect |
| age | +0.35 | +0.10 | **+0.35** | Total effect |
| height | -0.49 | -0.87 | **-0.64** | Adjusted assoc. |
| body mass | +0.23 | +0.54 | **+0.55** | Adjusted assoc. |
| body fat (%) | +0.56 | +0.10 | **+0.63** | Adjusted assoc. |
| ever-smoker | +0.10 | +0.03 | **+0.09** | Adjusted assoc. |
| pack-years | +0.19 | +0.05 | **+0.12** | Adjusted assoc. |

**Table 7.** Standardised β of each exposure on PC3 under the three estimands (targeted estimate in bold).

| Predictor | Unadjusted $\beta$ | Adjusted $\beta$ | Targeted $\beta$ (DAG) | Estimand |
| --- | --- | --- | --- | --- |
| sex (female) | +0.09 | -0.04 | **+0.09** | Total effect |
| age | -0.08 | -0.06 | **-0.09** | Total effect |
| height | -0.10 | +0.02 | **-0.11** | Adjusted assoc. |
| body mass | -0.26 | -0.28 | **-0.28** | Adjusted assoc. |
| body fat (%) | -0.11 | +0.01 | **-0.24** | Adjusted assoc. |
| ever-smoker | -0.04 | -0.00 | **-0.03** | Adjusted assoc. |
| pack-years | -0.07 | -0.02 | **-0.04** | Adjusted assoc. |

Westreich and Greenland (2013) caution us against the “Table 2 fallacy” of interpreting adjusted associations as causal effects without reference to the underlying DAG and the movement of an effect across unadjusted, adjusted, and targeted specifications. Based on the assumptions laid out in the DAG of Figure 2, sex and age are the only exposures whose targeted estimates can be read as total causal effects. Nevertheless, comparing their targeted estimates to the unadjusted and adjusted counterparts is instructive. For sex’s impact on PC1, comparing the various β coefficients implies that a large portion of the effect is mediated by the other metadata variables, notably height, body mass and body fat percentage. Still, even adjusting for these variables, it remains a strong predictor of PC1. PC2, in the meantime, is a more direct body-size and -shape mode, largely independent of sex.

The near-complete separation of the sexes along PC1 is shown in the targeted pair-plot of Figure 8.

**Figure 8.**
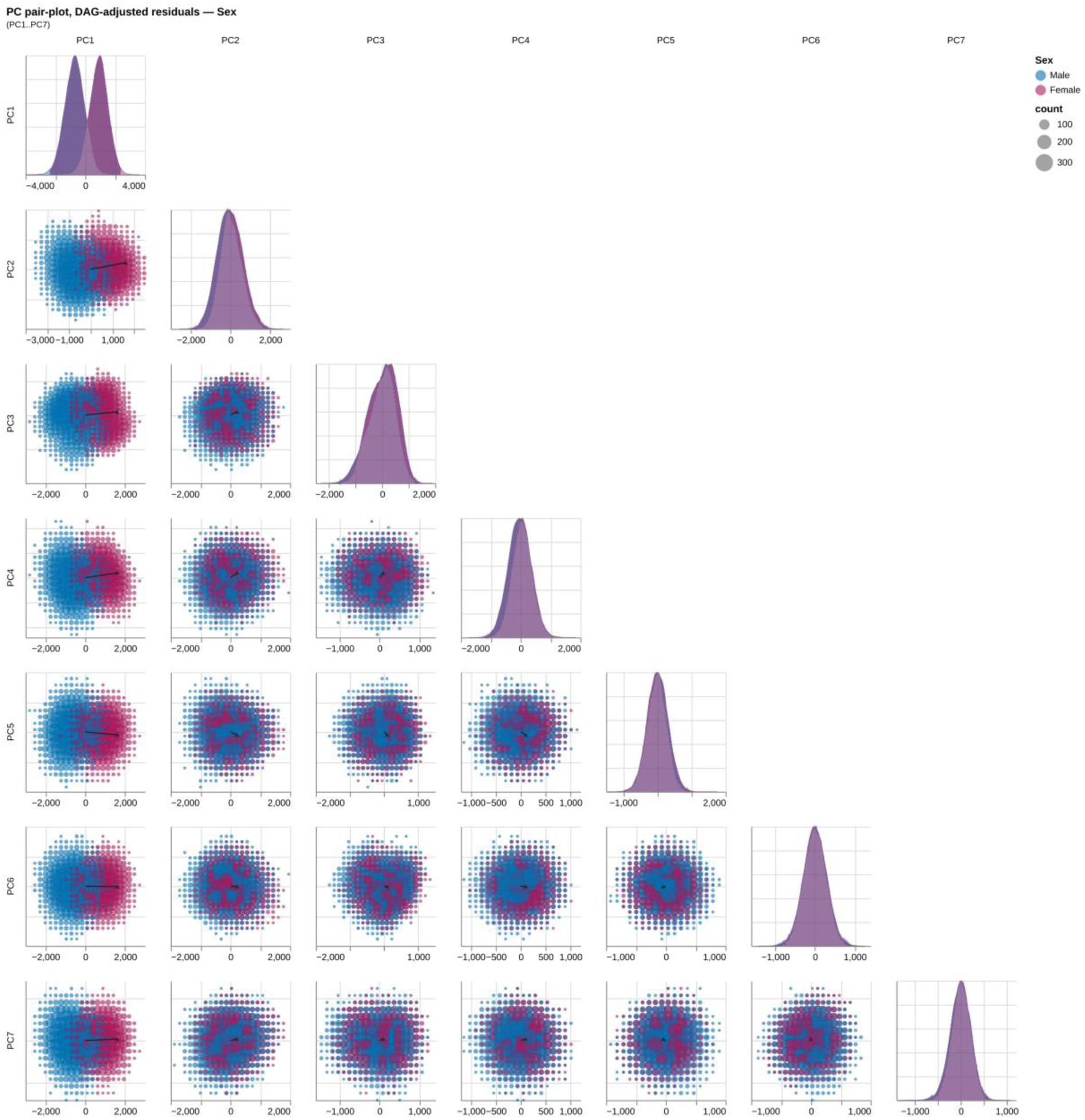
PC score pair-plot stratified by biological sex (targeted). Lower-triangular scatter of PC1–PC7 scores under the targeted total-effect specification for sex, male (blue) versus female (red); diagonal density panels show the per-PC distribution by sex. The marginal and adjusted companions are in the Supplement.

The same pair-plot construction coloured by each continuous exposure under the targeted specification is shown as a 2×2 grid in Figure 9.

**Figure 9.**
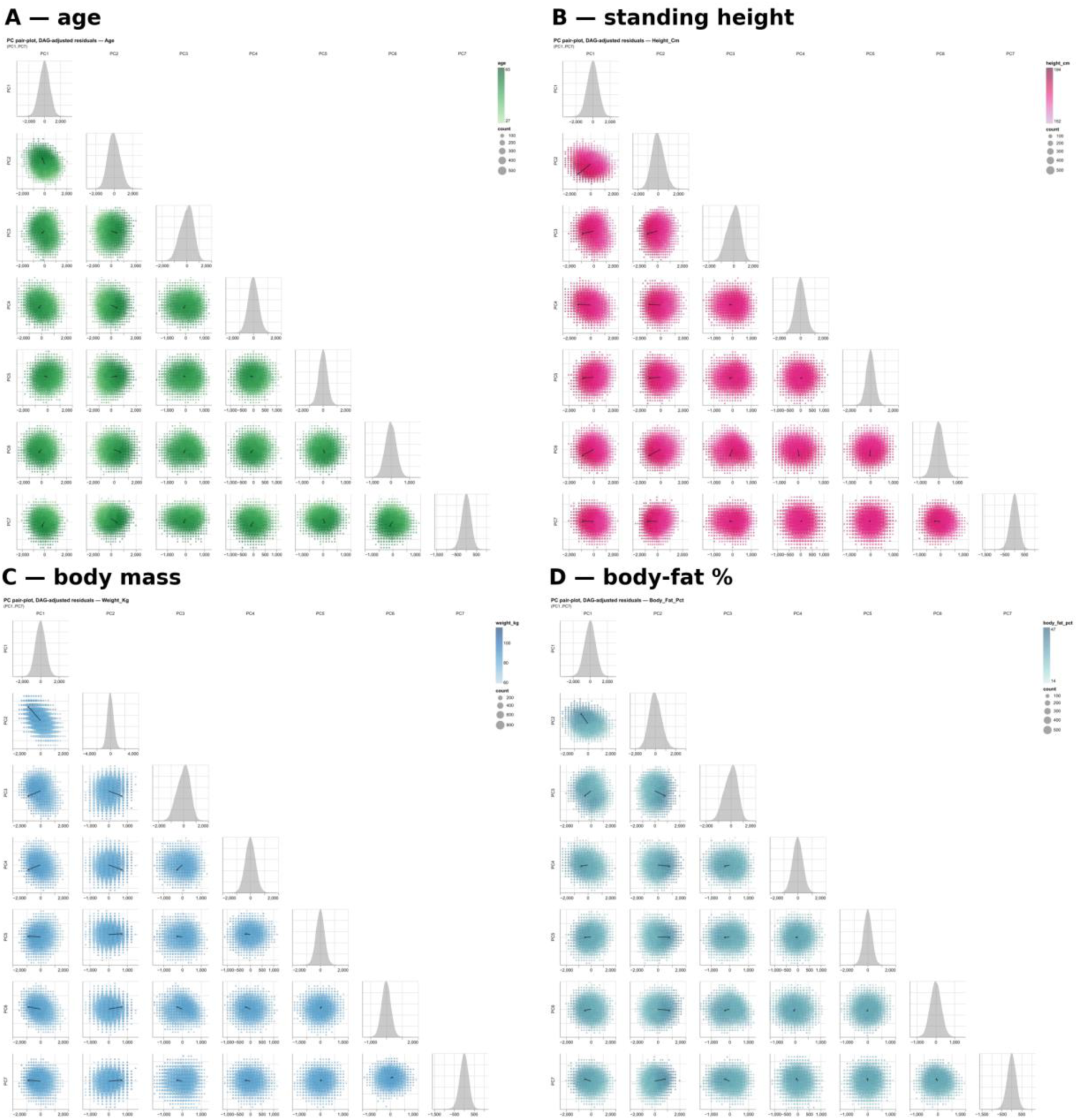
PC score pair-plots by continuous exposure (targeted). Off-diagonal bins coloured by the mean of each exposure under its targeted estimand: **(A)** age, **(B)** standing height, **(C)** body mass, **(D)** body-fat percentage. Ever-smoker and pack-years are omitted, their effect on the leading modes being negligible (Figure 7).

An anatomical reading of the principal modes – which per-rib shape descriptors load on which PC at which level – follows in §3.6.

### 3.6 Anatomical interpretation of principal modes

To attach anatomical meaning to the abstract principal modes characterised quantitatively in §3.2 and §3.5, we cross-walked each PC against the 14 per-rib shape descriptors of Methods §2.6, regressing each (rib position × descriptor) value against the cohort-z-scored PC score and tabulating standardised β coefficients per Methods §2.8.4. The resulting per-PC anatomical maps for the leading three modes – to be read against the ±2σ surface deformations of Figure 4, with continuous per-PC slider exploration available on the SSM Viewer page – are shown in Figure 10; analogous maps for PC4 through PC28 and the descriptor-anchored companion view are reported in Supplement S5. PC1 – the mode carrying most of the sex separation observed across PC1–PC7 and the one most strongly predicted by height and body mass – shows a strong positive association with the surface-to-volume ratio (SVR) across most ribs and negative associations with most other descriptors, most strongly with mesh volume, surface area, rib length and major axis

**Figure 10.**
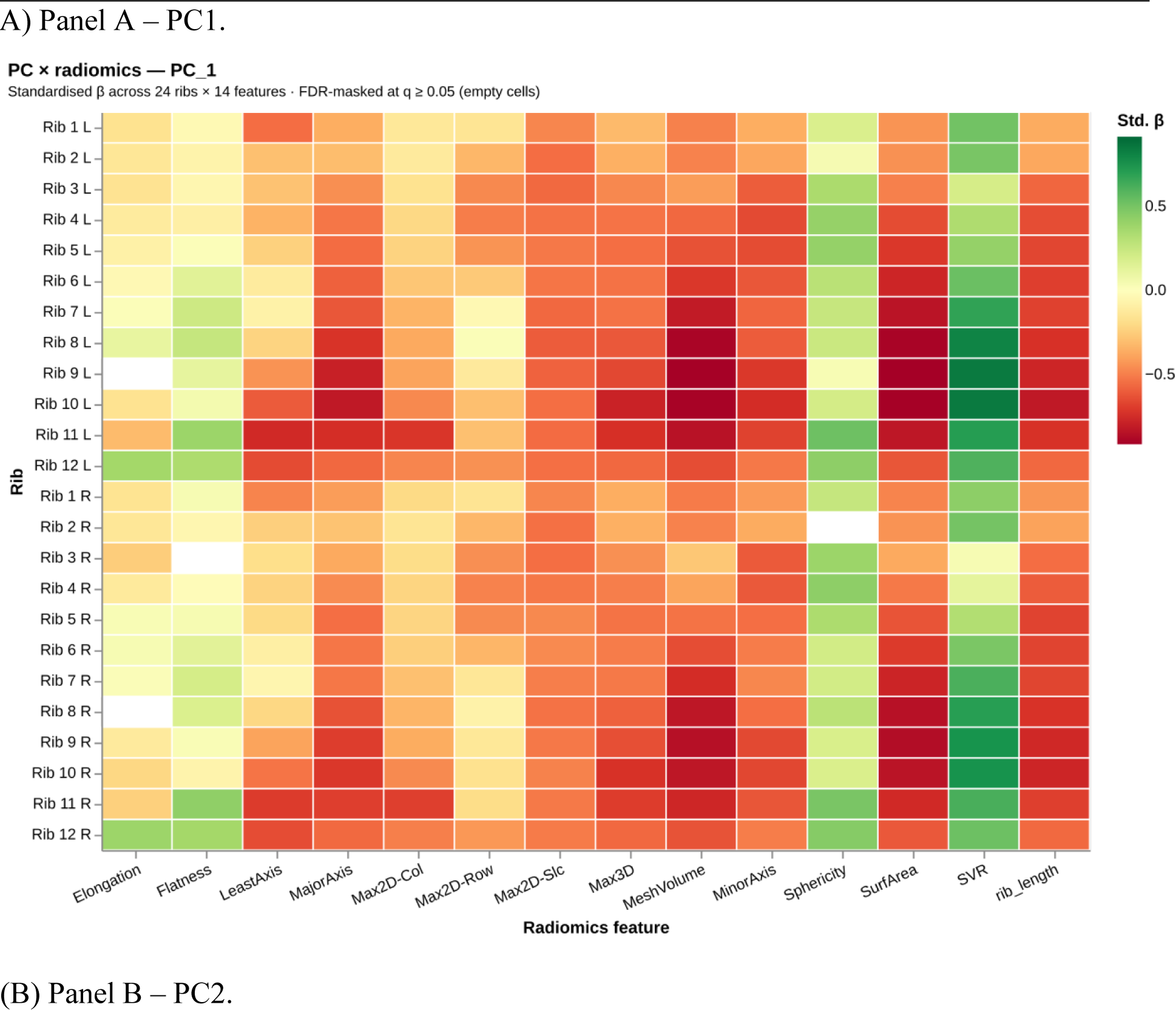
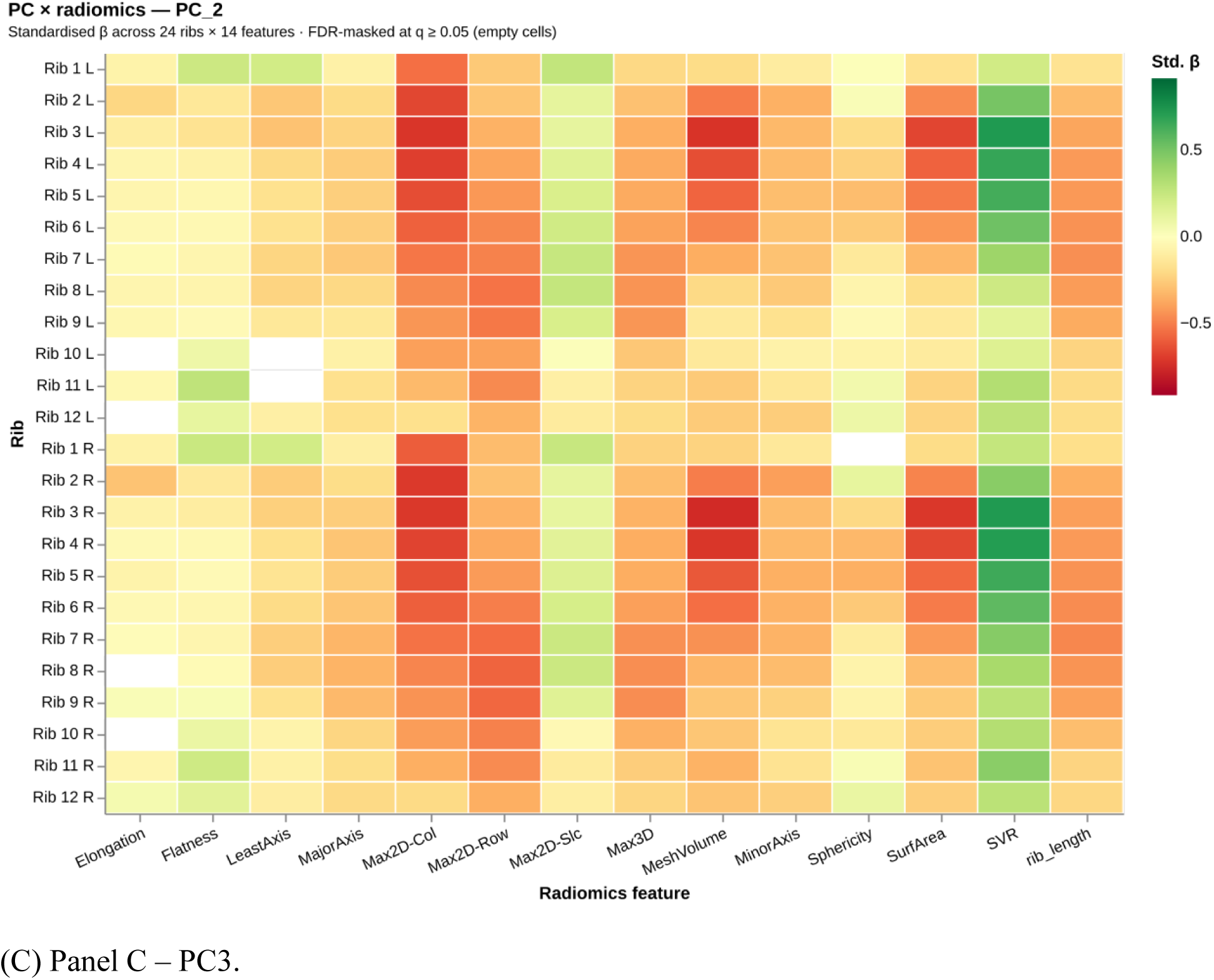
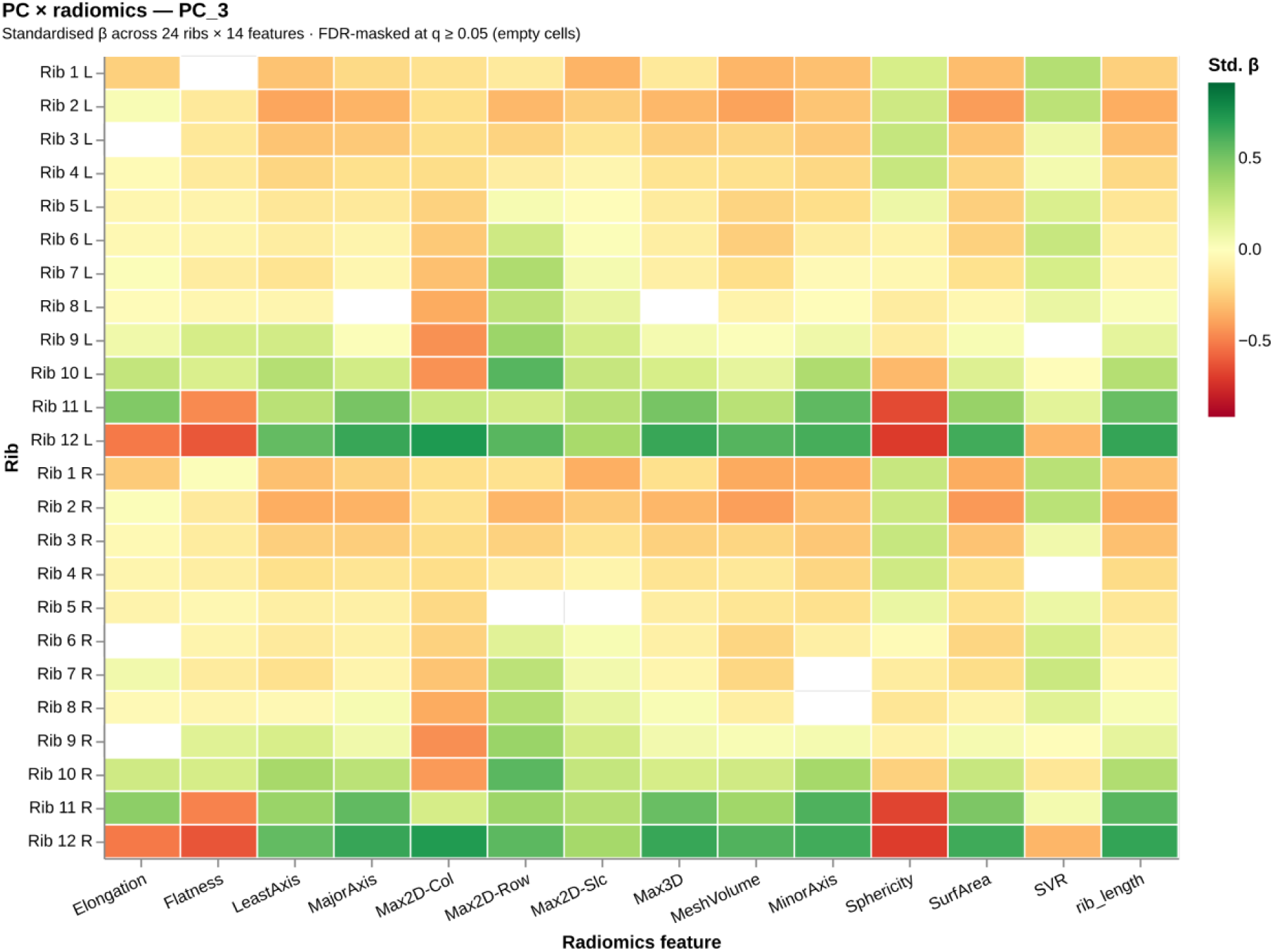
Per-PC anatomical cross-walk for the leading three modes. Each panel shows the standardised slope of every (rib position × shape descriptor) pair on the corresponding PC score (cohort-z-scored), rendered as a 24-row × 14-column heatmap; rows index rib positions (rib levels × side) and columns index the per-rib shape descriptors defined in Methods §2.6. Cells are FDR-masked at q ≥ 0.05 (empty). The panels are designed to be read alongside the ±2σ surface deformations of Figure 4: where the deformation moves rib N inward or outward, the corresponding row of this map identifies which descriptors carry that geometric change.

(Figure 10A). The effect peaks at ribs 8–10, the false but non-floating ribs. Although these descriptor signatures could be read as a uniform size increase, the corresponding ±2σ deformation in Figure 4 – or the same mode on the SSM Viewer – shows that PC1 is not a pure isotropic scaling: increasing PC1 also widens the cage, makes it more barrel-shaped, opens it anteriorly, and decreases the cranio-caudal inclination of the rib base, similar to the results of (Bellemare et al., 2003). Overall, the sexual dimorphism of these shape features fits the modes described in (Bellemare et al., 2003; S. A. Holcombe et al., 2017; S. Holcombe & Huang, 2023). Qualitatively, PC1 may be interpreted as a sexual-dimorphism axis.

PC2 – most strongly negatively predicted by height, and positively by body fat percentage and body mass – has its largest negative descriptor associations with Max2D-Col, Max2D-Row, the maximum 2D extents in the coronal and sagittal planes, and mesh volume (Figure 10B). The corresponding deformation in Figure 4 – or the SSM Viewer – shows the ribs becoming more horizontal, contracting vertically more than horizontally, and the overall ribcage becoming shorter. Qualitatively, PC2 may be interpreted as a slender-versus-stout body-shape axis.

PC3 – most strongly negatively predicted by body mass and body-fat percentage – concentrates its descriptor associations at the free ribs 11 and 12 (Figure 10C): negative on sphericity and positive on every other feature (recall that PyRadiomics defines Flatness and Elongation inversely; see Methods §2.6 and Table 2). The corresponding deformation in Figure 4 – or the SSM Viewer – is largely confined to those two ribs – low-PC3 cages have small free ribs, high-PC3 cages have large ones. Qualitatively, PC3 may be interpreted as a free-rib axis.

The inverse reading – how each metadata predictor reshapes the rib cage – is less easily expressed in per-rib descriptors, because a single covariate typically loads on several PC modes simultaneously and the resulting compound deformation is hard to summarise by any one geometric feature. The SSM Viewer’s per-predictor controls, which translate each covariate directly into its compound shape deformation, are more informative for this purpose. One noteworthy observation from such an exploration is that cumulative smoking exposure (pack-years) has a comparatively small effect on overall rib-cage shape, substantially smaller than that of body mass (cf. §3.5; Figure 7); the interpretation of this contrast is taken up in the Discussion.

Per-PC anatomical maps for PC4 onwards are reported in Supplement S5, and the SSM Viewer supports continuous exploration of any combination of the modes.

## 4. Discussion

### 4.1 Principal findings

Three principal findings frame the analyses of §3.

First, population-level rib-cage geometry is low-dimensional: a handful of orthogonal PC axes capture the majority of the shape variance observed across 26,275 adults aged 19–74.

Second, the leading modes admit consistent, named anatomical readings rather than abstract directions in shape space: PC1 a sexually dimorphic axis that widens the cage, opens it anteriorly, and reduces cranio-caudal inclination of the rib base; PC2 a slender-versus-stout body-habitus contrast on which height and body composition load in opposite directions; PC3 a free-rib-scale axis largely confined to ribs 11–12. That PC1 captures this coupled, multi-region deformation in a single component is itself a result – the surface representation expresses what lower-dimensional parametric families (S. A. Holcombe et al., 2017; Robinson et al., 2024) cannot recover from one mode.

Third, at this anatomical level, shape change is dominated by sex, about half of the sex difference on PC1 being attenuated after adjusting for body size and composition. When it comes to modifiable variables, shape change is modulated more by body mass and body fat percentage than by smoking exposure – a population-level association rather than a causal claim. Caveats on this reading are deferred to Strengths and limitations.

### 4.2 Comparison with prior work

The two anchor surface-correspondence rib SSMs, Shi et al. (2014) (n = 89, left hemithorax) and Wang et al. (2016) (n = 101, full cage), established the PCA-plus-demographic- regression formulation but were limited by sample size. The parametric centerline pipelines of S. A. Holcombe et al. (2016) and S. A. Holcombe et al. (2017) (n = 100 and 1,042 participants respectively, trauma-screened CT) traded surface correspondence for a logarithmic-spiral plus orientation representation and identified independent effects of age, sex, height and weight. Robinson et al. (2024) reached n = 1,719 with a fully automated CT pipeline but characterised shape through ten holistic scalar measurements, on the explicit argument that scalar descriptors are more interpretable than PCA modes, and deliberately restricted recruitment to ages 25–45 to remove ageing effects. The present model combines bilateral 24-rib surface correspondence with a cohort roughly two orders of magnitude larger than the prior surface SSMs and approximately an order of magnitude larger than the largest prior automated pipeline. The PyRadiomics cross- walk (Methods §2.6, §2.8.4) directly addresses the Robinson et al. (2024) interpretability concern: PC modes are readable as compound deformations whose loadings on named geometric descriptors can be tabulated rib-by-rib (see §3.6).

The imaging substrate also differs. NAKO is a population-based prospective cohort recruited on a standardised 3 T whole-body MRI protocol across multiple study centers (Bamberg et al., 2015; Peters et al., 2022; Rach et al., 2025), spanning the full adult age range and mitigating the acute-injury and clinical-indication selection bias that pervades prior (CT-based) rib-cage cohorts. Cohort-representativeness caveats are treated under Strengths and limitations.

The adiposity-over-pack-years reading of the barrel-chest signal sits naturally alongside the clinical literature: obesity is a documented restrictive thoracic load with measurable mechanical consequences (Mafort et al., 2016; Wehrmeister et al., 2012), whereas the classical barrel-chest of Sverzellati et al. (2013) and Lim et al. (2018) is a hyperinflation phenotype of established pulmonary disease rather than a smoking-exposure signal as such. The present model sees rib-cage geometry rather than lung parenchyma or airway calibre, so this calls into question the imagery of the barrel-chested smoker without refuting it.

Methodologically, the pipeline rests on established components – Gaussian Process Morphable Models in Scalismo (Luthi et al., 2018), SPINEPS-derived segmentation extended to a rib class on NAKO MRI (Graf et al., 2023; Möller et al., 2024), PyRadiomics descriptors as a cross-walk to named geometric features (Griethuysen et al., 2017). The contribution lies in their application at population scale on a non-clinical imaging substrate.

### 4.3 Strengths and limitations

The principal strengths follow from the combination set out above: a population-based prospective MRI cohort more than an order of magnitude larger than prior automated-pipeline work, dense per-vertex surface correspondence across all 24 ribs, and a PyRadiomics cross-walk that restores anatomical readability without collapsing the coupled multi-region variation that motivates the surface representation. Model fidelity is reported using the standard Styner triad of compactness, generalisation and specificity, and a held-out demographics-to-geometry prediction benchmark. An interactive viewer and scripts for generating rib cage models are provided to support downstream reuse as a population reference.

Several limitations should be read alongside these strengths.

The design is cross-sectional: the associations with body composition, age, and smoking exposure (§3.5) describe population-level structure rather than within-individual trajectories. NAKO recruitment carries a modest overall 15.6% baseline response rate with documented under-representation of lower-educated and non-German residents (Rach et al., 2025), so the resulting reference is appropriate for an adult population of predominantly central European ancestry and should not be extrapolated to other ancestries without appropriate measures.

MRI affords lower direct cortical-bone contrast and resolution than CT, and rib labels are produced by an automated deep-learning segmentation pipeline (Graf et al., 2023; Möller et al., 2024); the rib-count and registration-quality gates filter many but not all of its failure modes, and the residual error is propagated into the shape model rather than separately quantified against a manual reference.

The exclusion rate for non-canonical rib counts is higher than the prevalence of numerical variations reported for European cohorts (Brewin et al., 2009; Osiowski et al., 2024), and likely also reflects segmentation errors at the thoracolumbar (under-segmenting rib 12, or over-segmenting a pronounced L1 transverse process) and cervico-thoracic transition, as well as fused segmentations across multiple ribs. Excluding non-canonical rib counts is itself a limitation: the present SSM-PCA framework captures shape variation but not numerical variation. Further, direct numerical comparison of cross-sectional cortical metrics with CT-derived references (S. A. Holcombe et al., 2020; S. Holcombe & Huang, 2023) is out of scope of this work.

At the descriptor level, individual partial R² values remain modest (§3.4); per-rib effect-size statements are best interpreted alongside the PC-level analyses where signal concentrates. The small smoking signal at the level of overall rib-cage geometry does not refute the established COPD-associated barrel chest: the present model does not stratify by pulmonary phenotype, and an emphysema-positive subgroup may show a clearer signal. Nor does it argue against the well-established consequences of smoking for pulmonary function, which lie outside what overall rib- cage geometry can capture. Finally, the model represents rib-cage morphology only; other thoracic factors – notably thoracic-spine curvature – enter only through their effect on rib pose.

### 4.4 Conclusion

We present a population-scale statistical shape model of the complete 24-rib cage built from 26,275 standardised NAKO whole-body MRI scans, with dense per-vertex correspondence and a per-rib radiomics cross-walk that recovers named geometric descriptors from the principal modes. For example, PC1–PC3 admit consistent anatomical readings – a sexually dimorphic axis, a slender-versus-stout body-habitus contrast, and a free-rib axis at ribs 11–12. We show that the population-level signature of cumulative smoking exposure is substantially smaller than that of body mass or body-fat percentage. The model is intended as a population-representative geometric reference against which donor-derived finite-element human-body models can be benchmarked and morphed (S. A. Holcombe et al., 2020; Iwamoto et al., 2015; Vavalle et al., 2012), as a substrate for simulation work that links external rib geometry to biomechanical response (S. Holcombe & Huang, 2023; Iraeus et al., 2020), and as a foundation for further large-cohort statistical shape modelling. Mean shape, PC modes, and metadata loadings are released alongside an interactive browser (see the SSM Viewer page and the Data and code availability section).

## 5. Data and code availability

Full source code, configuration for the canonical run, and the figure bundle reproduced in this manuscript are available at https://github.com/AnatolAicher/NAKO-Ribcage-SSM. The mean shape model, PC modes, and metadata loadings are released with that repository and archived on Zenodo at https://doi.org/10.5281/zenodo.22230693 (version 1.1.0: https://doi.org/10.5281/zenodo.22237859). Due to data privacy regulations in the context of the NAKO cohort, the individual-level metadata and imaging data used in this project cannot be released publicly. Specific data may be available on request from the authors, subject to NAKO data access policies.

## 6. Acknowledgements

### 6.1 Funding

This project was conducted with data from the German National Cohort (NAKO) (www.nako.de). The NAKO is funded by the Federal Ministry of Research, Technology and Space (BMFTR) [project funding reference numbers: 01ER1301A/B/C, 01ER1511D, 01ER1801A/B/C/D and 01ER2301A/B/C], Federal States of Germany and the Helmholtz Association, the participating universities and the institutes of the Leibniz Association.

### 6.3 Contributions

Conceptualization: [A.A.]; Methodology: [A.A., H.M.]; Software: [A.A., H.M.]; Validation: [H.M., M.L., B.M., F.E., R.G.]; Formal analysis: [A.A.]; Investigation: [A.A.]; Writing – original draft: [A.A.]; Writing – review and editing: [all authors]; Visualization: [A.A.]; Supervision: [H.M.]; Resources: [J.K., T.F., T.Kr., S.W., C.S., C.O.S., M.W., S.R., J.H., J.D., T.Ke., T.N., T.P., F.B.]; Data curation: [H.M.]; Funding acquisition: [J.K., T.F., T.P., T.N.].

All authors have read and approved the final version of the manuscript.

## Data Availability

Full source code, the configuration of the canonical run, and the figure bundle reproduced in the manuscript are available at https://github.com/AnatolAicher/NAKO-Ribcage-SSM. The mean shape, principal modes, and metadata loadings of the statistical shape model are released with that repository and archived on Zenodo (concept DOI 10.5281/zenodo.22230693; version 1.1.0, 10.5281/zenodo.22237859). An interactive version of the manuscript with a 3D model viewer is available at https://anatolaicher.github.io/NAKO-Ribcage-Manuscript/. Individual-level NAKO imaging and phenotype data cannot be shared publicly because of data-protection regulations; they can be requested by researchers through the NAKO application portal (www.nako.de/transferhub), subject to NAKO data-access policies.

https://doi.org/10.5281/zenodo.22230693

https://doi.org/10.5281/zenodo.22237859

https://anatolaicher.github.io/NAKO-Ribcage-Manuscript/

https://www.nako.de/transferhub

## 6.4 Acknowledgements

We thank all participants of the German National Cohort (NAKO) and the staff of this research initiative. Members and affiliations of the NAKO Investigator Consortium can be accessed via www.nako.de/principal-investigators.

## References

Bamberg, F., Kauczor, H.-U., Weckbach, S., Schlett, C. L., Forsting, M., Ladd, S. C., Greiser, K. H., Weber, M.-A., Schulz-Menger, J., Niendorf, T., Pischon, T., Caspers, S., Amunts, K., Berger, K., Bülow, R., Hosten, N., Hegenscheid, K., Kröncke, T., Linseisen, J., … Völzke, H. and. (2015). Whole-body MR imaging in the german national cohort: Rationale, design and technical background. Radiology, 277(1), 206–220. 10.1148/radiol.2015142272

Bellemare, F., Jeanneret, A., & Couture, J. (2003). Sex differences in thoracic dimensions and configuration. American Journal of Respiratory and Critical Care Medicine, 168(3), 305–312. 10.1164/rccm.200208-876oc

Benjamini, Y., & Hochberg, Y. (1995). Controlling the false discovery rate: A practical and powerful approach to multiple testing. Journal of the Royal Statistical Society Series B: Statistical Methodology, 57(1), 289–300. 10.1111/j.2517-6161.1995.tb02031.x

Brewin, J., Hill, M., & Ellis, H. (2009). The prevalence of cervical ribs in a london population. Clinical Anatomy, 22(3), 331–336. 10.1002/ca.20774

Gower, J. C. (1975). Generalized procrustes analysis. Psychometrika, 40(1), 33–51. 10.1007/bf02291478

Graf, R., Schmitt, J., Schlaeger, S., Möller, H. K., Sideri-Lampretsa, V., Sekuboyina, A., Krieg, S. M., Wiestler, B., Menze, B., Rueckert, D., & Kirschke, J. S. (2023). Denoising diffusion-based MRI to CT image translation enables automated spinal segmentation. European Radiology Experimental, 7(1). 10.1186/s41747-023-00385-2

Griethuysen, J. J. M. van, Fedorov, A., Parmar, C., Hosny, A., Aucoin, N., Narayan, V., Beets-Tan, R. G. H., Fillion-Robin, J.-C., Pieper, S., & Aerts, H. J. W. L. (2017). Computational radiomics system to decode the radiographic phenotype. Cancer Research, 77(21), e104–e107. 10.1158/0008-5472.can-17-0339

Harris, C. R., Millman, K. J., Walt, S. J. van der, Gommers, R., Virtanen, P., Cournapeau, D., Wieser, E., Taylor, J., Berg, S., Smith, N. J., Kern, R., Picus, M., Hoyer, S., Kerkwijk, M. H. van, Brett, M., Haldane, A., Río, J. F. del, Wiebe, M., Peterson, P., … Oliphant, T. E. (2020). Array programming with NumPy. Nature, 585(7825), 357–362. 10.1038/s41586-020-2649-2

Holcombe, S. A., Agnew, A. M., Derstine, B., & Wang, S. C. (2020). Comparing FE human body model rib geometry to population data. Biomechanics and Modeling in Mechanobiology, 19(6), 2227–2239. 10.1007/s10237-020-01335-2

Holcombe, S. A., Wang, S. C., & Grotberg, J. B. (2016). Modeling female and male rib geometry with logarithmic spirals. Journal of Biomechanics, 49(13), 2995–3003. 10.1016/j.jbiomech.2016.07.021

Holcombe, S. A., Wang, S. C., & Grotberg, J. B. (2017). The effect of age and demographics on rib shape. Journal of Anatomy, 231(2), 229–247. 10.1111/joa.12632

Holcombe, S., & Huang, Y. (2023). Cross-sectional properties of rib geometry from an adult population. Frontiers in Bioengineering and Biotechnology, 11. 10.3389/fbioe.2023.1158242

Iraeus, J., Brolin, K., & Pipkorn, B. (2020). Generic finite element models of human ribs, developed and validated for stiffness and strain prediction – to be used in rib fracture risk evaluation for the human population in vehicle crashes. Journal of the Mechanical Behavior of Biomedical Materials, 106, 103742. 10.1016/j.jmbbm.2020.103742

Iwamoto, M., Nakahira, Y., & Kimpara, H. (2015). Development and validation of the total HUman model for safety (THUMS) toward further understanding of occupant injury mechanisms in precrash and during crash. Traffic Injury Prevention, 16(sup1), S36–S48. 10.1080/15389588.2015.1015000

Larsson, K.-J., Iraeus, J., Holcombe, S., & Pipkorn, B. (2023). Influences of human thorax variability on population rib fracture risk prediction using human body models. Frontiers in Bioengineering and Biotechnology, 11. 10.3389/fbioe.2023.1154272

Larsson, K.-J., Östh, J., Iraeus, J., & Pipkorn, B. (2024). A first step toward a family of morphed human body models enabling prediction of population injury outcomes. Journal of Biomechanical Engineering, 146(3). 10.1115/1.4064033

Liebsch, C., Graf, N., Appelt, K., & Wilke, H.-J. (2017). The rib cage stabilizes the human thoracic spine: An in vitro study using stepwise reduction of rib cage structures. PLOS ONE, 12(6), e0178733. 10.1371/journal.pone.0178733

Liebsch, C., Seiffert, T., Vlcek, M., Beer, M., Huber-Lang, M., & Wilke, H.-J. (2019). Patterns of serial rib fractures after blunt chest trauma: An analysis of 380 cases. PLOS ONE, 14(12), e0224105. 10.1371/journal.pone.0224105

Liebsch, C., & Wilke, H.-J. (2022). How does the rib cage affect the biomechanical properties of the thoracic spine? A systematic literature review. Frontiers in Bioengineering and Biotechnology, 10. 10.3389/fbioe.2022.904539

Lim, S. J., Kim, J.-Y., Lee, S. J., Lee, G. D., Cho, Y. J., Jeong, Y. Y., Jeon, K. N., Lee, J. D., Kim, J. R., & Kim, H. C. (2018). Altered thoracic cage dimensions in patients with chronic obstructive pulmonary disease. Tuberculosis and Respiratory Diseases, 81(2), 123. 10.4046/trd.2017.0095

Luthi, M., Gerig, T., Jud, C., & Vetter, T. (2018). Gaussian process morphable models. IEEE Transactions on Pattern Analysis and Machine Intelligence, 40(8), 1860–1873. 10.1109/tpami.2017.2739743

Mafort, T. T., Rufino, R., Costa, C. H., & Lopes, A. J. (2016). Obesity: Systemic and pulmonary complications, biochemical abnormalities and impairment of lung function. Multidisciplinary Respiratory Medicine, 11(1). 10.1186/s40248-016-0066-z

McKinney, W. (2010). Data structures for statistical computing in python. Proceedings of the 9th Python in Science Conference, 56–61. 10.25080/majora-92bf1922-00a

Möller, H., Dima, A., Keinert-Weth, B., Graf, R., Atad, M., Paetzold, J., Jungmann, F., Braren, R., Kofler, F., Menze, B., Rueckert, D., Kirschke, J. S., & Schön, H. (2026). Automated thoracolumbar stump rib detection and analysis in a large CT cohort. AI, 7(6), 224. 10.3390/ai7060224

Möller, H., Graf, R., Schmitt, J., Keinert, B., Schön, H., Atad, M., Sekuboyina, A., Streckenbach, F., Kofler, F., Kroencke, T., Bette, S., Willich, S. N., Keil, T., Niendorf, T., Pischon, T., Endemann, B., Menze, B., Rueckert, D., & Kirschke, J. S. (2024). SPINEPS–automatic whole spine segmentation of T2-weighted MR images using a two-phase approach to multi-class semantic and instance segmentation. European Radiology, 35(3), 1178–1189. 10.1007/s00330-024-11155-y

Osiowski, M., Osiowski, A., Preinl, M., Stolarz, K., Klepinowski, T., Jasiewicz, B., & Taterra, D. (2024). Prevalence and characteristics of lumbar ribs: A meta-analysis with anatomical and clinical considerations. Surgical and Radiologic Anatomy, 46(12), 2057– 2066. 10.1007/s00276-024-03504-9

Pedregosa, F., Varoquaux, G., Gramfort, A., Michel, V., Thirion, B., Grisel, O., Blondel, M., Prettenhofer, P., Weiss, R., Dubourg, V., Vanderplas, J., Passos, A., Cournapeau, D., Brucher, M., Perrot, M., & Duchesnay, E. (2011). Scikit-learn: Machine learning in Python. Journal of Machine Learning Research, 12, 2825–2830.

Peek, J., Beks, R. B., Hietbrink, F., De Jong, M. B., Heng, M., Beeres, F. J. P., IJpma, F. F. A., Leenen, L. P. H., Groenwold, R. H. H., & Houwert, R. M. (2020). Epidemiology and outcome of rib fractures: A nationwide study in the netherlands. European Journal of Trauma and Emergency Surgery, 48(1), 265–271. 10.1007/s00068-020-01412-2

Peek, J., Ochen, Y., Saillant, N., Groenwold, R. H. H., Leenen, L. P. H., Uribe-Leitz, T., Houwert, R. M., & Heng, M. (2020). Traumatic rib fractures: A marker of severe injury. A nationwide study using the national trauma data bank. Trauma Surgery & Acute Care Open, 5(1), e000441. 10.1136/tsaco-2020-000441

Peters, A., Peters, A., Greiser, K. H., Göttlicher, S., Ahrens, W., Albrecht, M., Bamberg, F., Bärnighausen, T., Becher, H., Berger, K., Beule, A., Boeing, H., Bohn, B., Bohnert, K., Braun, B., Brenner, H., Bülow, R., Castell, S., Damms-Machado, A., … Zschocke, J. (2022). Framework and baseline examination of the german national cohort (NAKO). European Journal of Epidemiology, 37(10), 1107–1124. 10.1007/s10654-022-00890-5

Rach, S., Sand, M., Reineke, A., Becher, H., Greiser, K. H., Wolf, K., Wirkner, K., Schmidt, C. O., Schipf, S., Jöckel, K.-H., Krist, L., Ahrens, W., Brenner, H., Castell, S., Gastell, S., Harth, V., Holleczek, B., Ittermann, T., Janisch-Fabian, S., … Günther, K. (2025). The baseline examinations of the german national cohort (NAKO): Recruitment protocol, response and weighting. European Journal of Epidemiology, 40(4), 475–489. 10.1007/s10654-025-01219-8

Robinson, A., Zheng, B., Kleeck, B. W. von, Tan, J., & Gayzik, F. S. (2024). Holistic shape variation of the rib cage in an adult population. Frontiers in Bioengineering and Biotechnology, 12. 10.3389/fbioe.2024.1432911

Satyanarayan, A., Moritz, D., Wongsuphasawat, K., & Heer, J. (2017). Vega-lite: A grammar of interactive graphics. IEEE Transactions on Visualization and Computer Graphics, 23(1), 341–350. 10.1109/tvcg.2016.2599030

Schönemann, P. H. (1966). A generalized solution of the orthogonal procrustes problem. Psychometrika, 31(1), 1–10. 10.1007/bf02289451

Seabold, S., & Perktold, J. (2010). Statsmodels: Econometric and statistical modeling with python. Proceedings of the 9th Python in Science Conference, 92–96. 10.25080/majora-92bf1922-011

Shi, X., Cao, L., Reed, M. P., Rupp, J. D., Hoff, C. N., & Hu, J. (2014). A statistical human rib cage geometry model accounting for variations by age, sex, stature and body mass index. Journal of Biomechanics, 47(10), 2277–2285. 10.1016/j.jbiomech.2014.04.045

Styner, M. A., Rajamani, K. T., Nolte, L.-P., Zsemlye, G., Székely, G., Taylor, C. J., & Davies, R. H. (2003). Evaluation of 3D correspondence methods for model building. In Information processing in medical imaging (pp. 63–75). Springer Berlin Heidelberg. 10.1007/978-3-540-45087-0_6

Sullivan, C., & Kaszynski, A. (2019). PyVista: 3D plotting and mesh analysis through a streamlined interface for the visualization toolkit (VTK). Journal of Open Source Software, 4(37), 1450. 10.21105/joss.01450

Sverzellati, N., Colombi, D., Randi, G., Pavarani, A., Silva, M., Walsh, S. L., Pistolesi, M., Alfieri, V., Chetta, A., Vaccarezza, M., Vitale, M., & Pastorino, U. (2013). Computed tomography measurement of rib cage morphometry in emphysema. PLoS ONE, 8(7), e68546. 10.1371/journal.pone.0068546

Umeyama, S. (1991). Least-squares estimation of transformation parameters between two point patterns. IEEE Transactions on Pattern Analysis and Machine Intelligence, 13(4), 376– 380. 10.1109/34.88573

VanderPlas, J., Granger, B., Heer, J., Moritz, D., Wongsuphasawat, K., Satyanarayan, A., Lees, E., Timofeev, I., Welsh, B., & Sievert, S. (2018). Altair: Interactive statistical visualizations for python. Journal of Open Source Software, 3(32), 1057. 10.21105/joss.01057

Vavalle, N. A., Moreno, D. P., Rhyne, A. C., Stitzel, J. D., & Gayzik, F. S. (2012). Lateral impact validation of a geometrically accurate full body finite element model for blunt injury prediction. Annals of Biomedical Engineering, 41(3), 497–512. 10.1007/s10439-012-0684-3

Virtanen, P., Gommers, R., Oliphant, T. E., Haberland, M., Reddy, T., Cournapeau, D., Burovski, E., Peterson, P., Weckesser, W., Bright, J., Walt, S. J. van der, Brett, M., Wilson, J., Millman, K. J., Mayorov, N., Nelson, A. R. J., Jones, E., Kern, R., Larson, E., … Vázquez-Baeza, Y. (2020). SciPy 1.0: Fundamental algorithms for scientific computing in python. Nature Methods, 17(3), 261–272. 10.1038/s41592-19-0686-2

Wang, Y., Cao, L., Bai, Z., Reed, M. P., Rupp, J. D., Hoff, C. N., & Hu, J. (2016). A parametric ribcage geometry model accounting for variations among the adult population. Journal of Biomechanics, 49(13), 2791–2798. 10.1016/j.jbiomech.2016.06.020

Watkins, R., Watkins, R., Williams, L., Ahlbrand, S., Garcia, R., Karamanian, A., Sharp, L., Vo, C., & Hedman, T. (2005). Stability provided by the sternum and rib cage in the thoracic spine. Spine, 30(11), 1283–1286. 10.1097/01.brs.0000164257.69354.bb

Wehrmeister, F. C., Menezes, A. M. B., Muniz, L. C., Martínez-Mesa, J., Domingues, M. R., & Horta, B. L. (2012). Waist circumference and pulmonary function: A systematic review and meta-analysis. Systematic Reviews, 1(1). 10.1186/2046-4053-1-55

Westreich, D., & Greenland, S. (2013). The table 2 fallacy: Presenting and interpreting confounder and modifier coefficients. American Journal of Epidemiology, 177(4), 292– 298. 10.1093/aje/kws412

