## Supplement for "How Sex, Age, Adiposity, and Smoking Shape the Human Rib Cage: Evidence from 26,275 Whole-Body MRIs across the German National Cohort (NAKO)"

### Supplementary Methods and Figures

#### S1 Cohort and ingestion diagnostics

##### S1.1 Per-participant rib-side count distribution

*Supplementary Table 1: Distribution of per-participant rib-side counts among the candidate cohort. The inclusion criterion (Methods §2.5) requires exactly 24 rib sides; participants with different counts are excluded. Low counts (≤ 23) are consistent with transitional thoracolumbar vertebrae, missing segmentations or merged segmentations; high counts (≥ 25) with supernumerary cervical or lumbar ribs.*

| Rib sides per participant | n participants | % |
|---|---|---|
| 8 | 1 | 0.0% |
| 12 | 1 | 0.0% |
| 17 | 1 | 0.0% |
| 18 | 7 | 0.0% |
| 19 | 6 | 0.0% |
| 20 | 38 | 0.1% |
| 21 | 28 | 0.1% |
| 22 | 551 | 1.8% |
| 23 | 795 | 2.6% |
| 24 | 27,502 | 91.0% |
| 25 | 693 | 2.3% |
| 26 | 595 | 2.0% |

#### S1.2 Per-variable missingness across the joined cohort

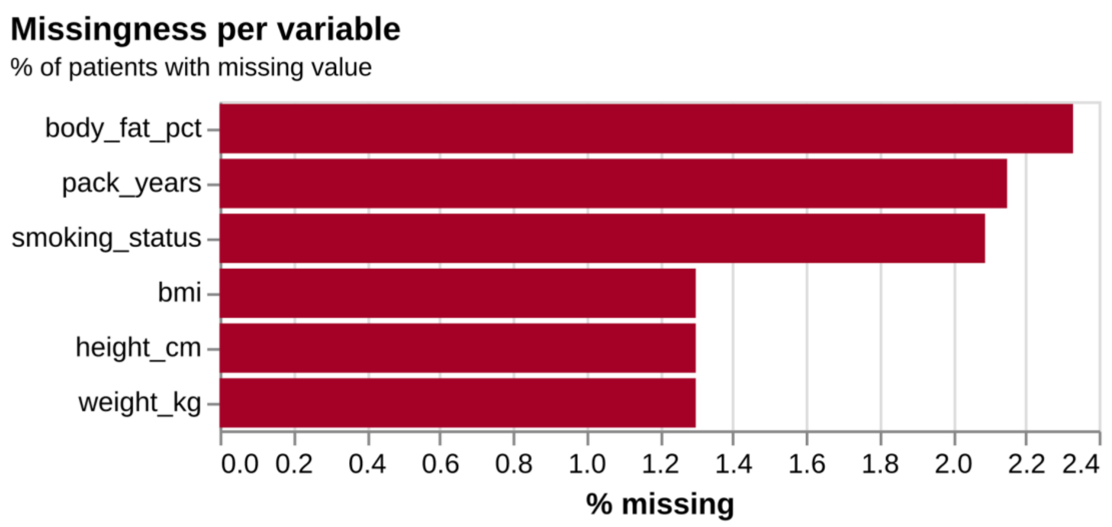


*Supplementary Figure 1:* ***Missingness across baseline metadata variables.*** *Computed on the inner-joined cohort (n = 30,218) prior to quality-control exclusions.*

#### S1.3 Predictor collinearity

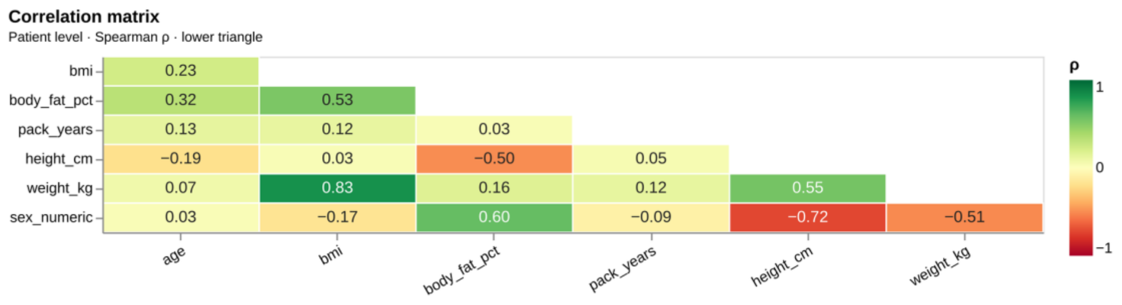


*Supplementary Figure 2:* ***Pairwise Pearson correlations between continuous metadata predictors.*** *Body mass, BMI, and body-fat percentage cluster tightly, motivating the multivariable adjusted regression design of Methods §2.8.2 rather than relying on bivariate associations.*

#### S1.4 Descriptor collinearity

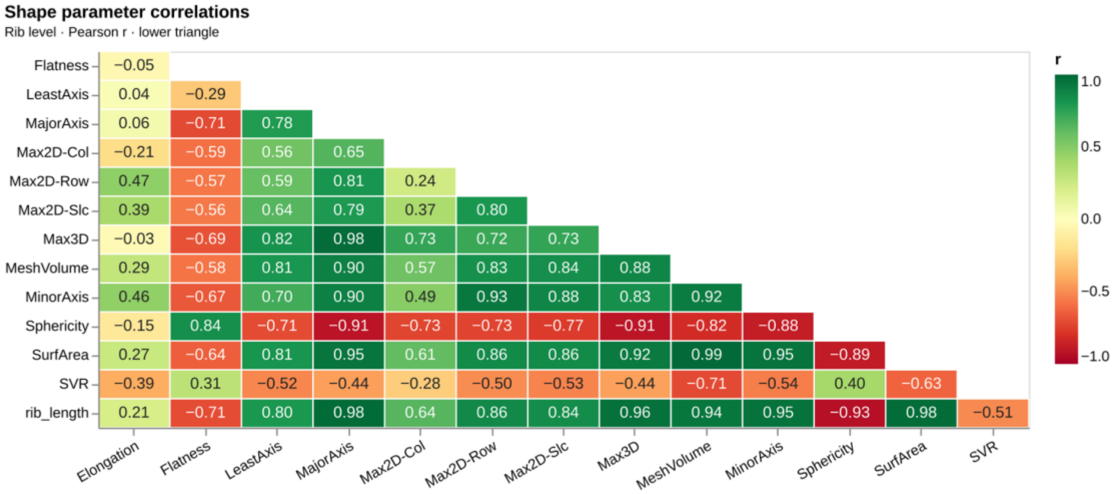


|  | Elongation | Flatness | LeastAxis | MajorAxis | Max2D-Col | Max2D-Row | Max2D-Slc | Max3D | MeshVolume | MinorAxis | Sphericity | SurfArea | SVR |
|---|---|---|---|---|---|---|---|---|---|---|---|---|---|
| Flatness | −0.05 | | | | | | | | | | | | |
| LeastAxis | 0.04 | −0.29 | | | | | | | | | | | |
| MajorAxis | 0.06 | −0.71 | 0.78 | | | | | | | | | | |
| Max2D-Col | −0.21 | −0.59 | 0.56 | 0.65 | | | | | | | | | |
| Max2D-Row | 0.47 | −0.57 | 0.59 | 0.81 | 0.24 | | | | | | | | |
| Max2D-Slc | 0.39 | −0.56 | 0.64 | 0.79 | 0.37 | 0.80 | | | | | | | |
| Max3D | −0.03 | −0.69 | 0.82 | 0.98 | 0.73 | 0.72 | 0.73 | | | | | | |
| MeshVolume | 0.29 | −0.58 | 0.81 | 0.90 | 0.57 | 0.83 | 0.84 | 0.88 | | | | | |
| MinorAxis | 0.46 | −0.67 | 0.70 | 0.90 | 0.49 | 0.93 | 0.88 | 0.83 | 0.92 | | | | |
| Sphericity | −0.15 | 0.84 | −0.71 | −0.91 | −0.73 | −0.73 | −0.77 | −0.91 | −0.82 | −0.88 | | | |
| SurfArea | 0.27 | −0.64 | 0.81 | 0.95 | 0.61 | 0.86 | 0.86 | 0.92 | 0.99 | 0.95 | −0.89 | | |
| SVR | −0.39 | 0.31 | −0.52 | −0.44 | −0.28 | −0.50 | −0.53 | −0.44 | −0.71 | −0.54 | 0.40 | −0.63 | |
| rib_length | 0.21 | −0.71 | 0.80 | 0.98 | 0.64 | 0.86 | 0.84 | 0.96 | 0.94 | 0.95 | −0.93 | 0.98 | −0.51 |

*Supplementary Figure 3:* ***Pairwise Pearson correlations between per-rib shape descriptors.*** *Computed on participant-level per-rib means. Voxel volume was dropped at the analysis stage (Methods §2.6) due to conceptual identity (and perfect collinearity) with mesh volume.*

##### S1.5 Distribution and normality of shape descriptors

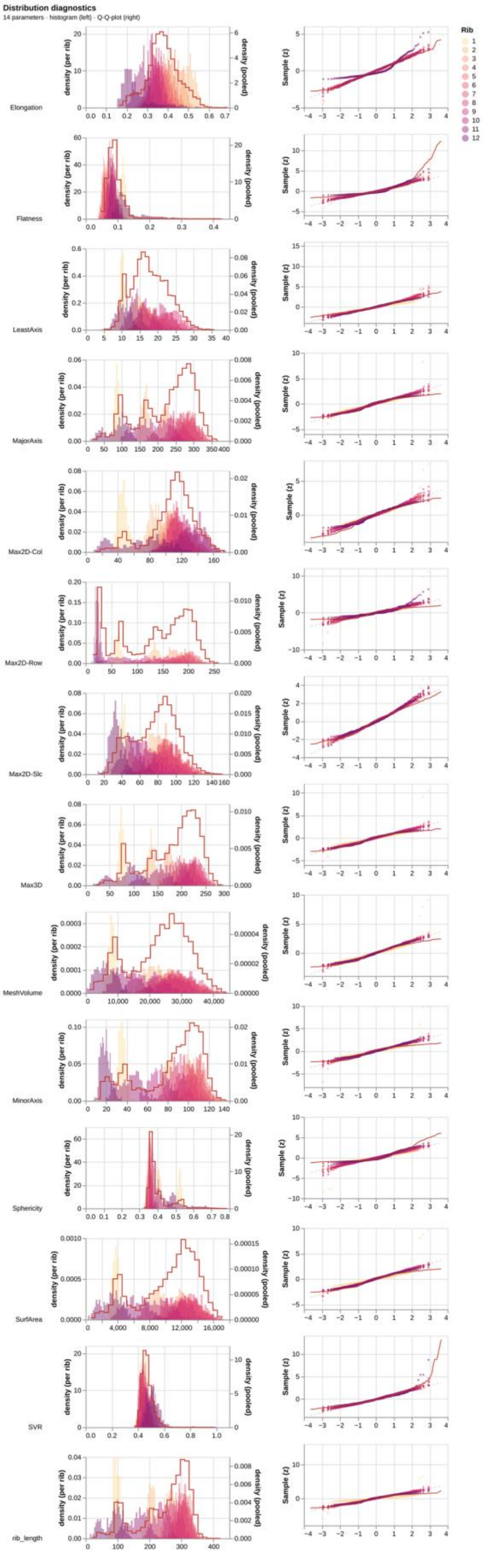


*Supplementary Figure 4:* ***Per-descriptor histograms with Q-Q overlays.*** *Q-Q axes are z-standardised so that the y =* [illegible]

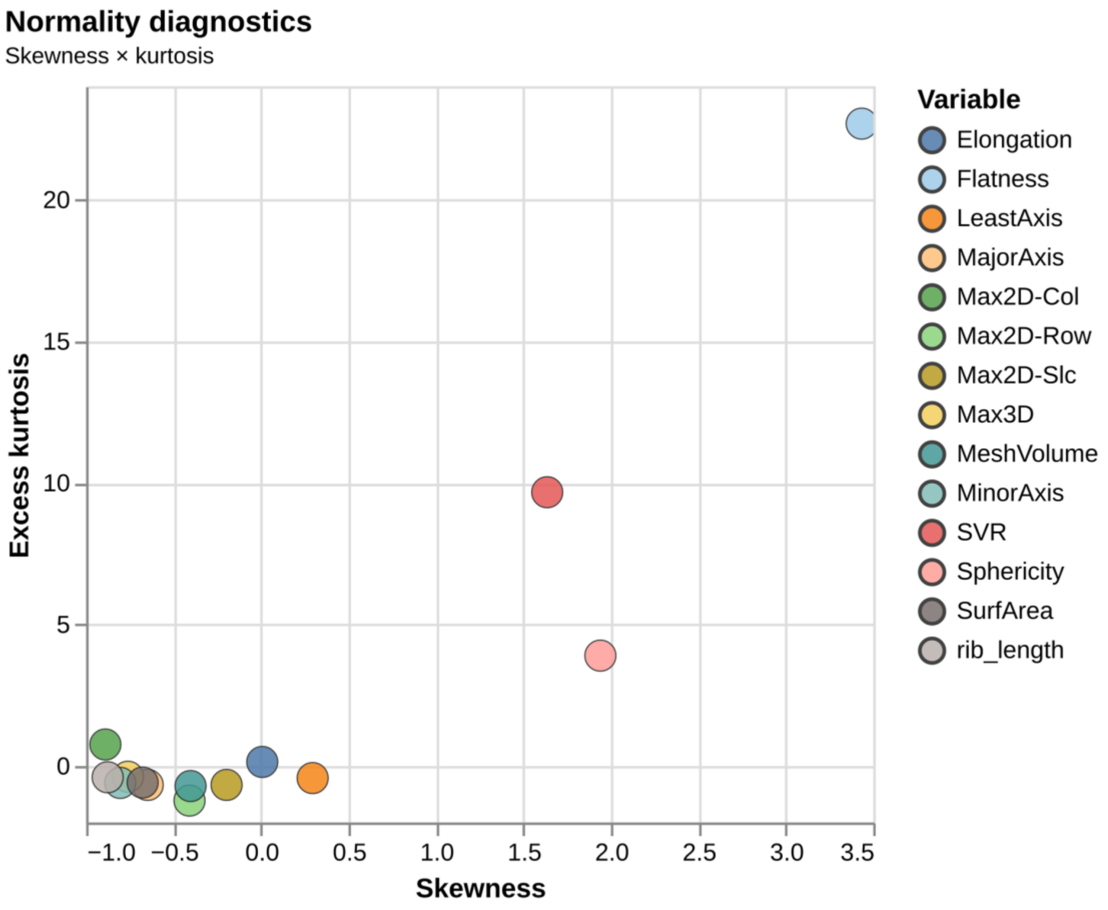


*Supplementary Figure 5:* ***Shapiro–Wilk normality summary across shape descriptors.***

#### S1.6 Predictor distributions by sex

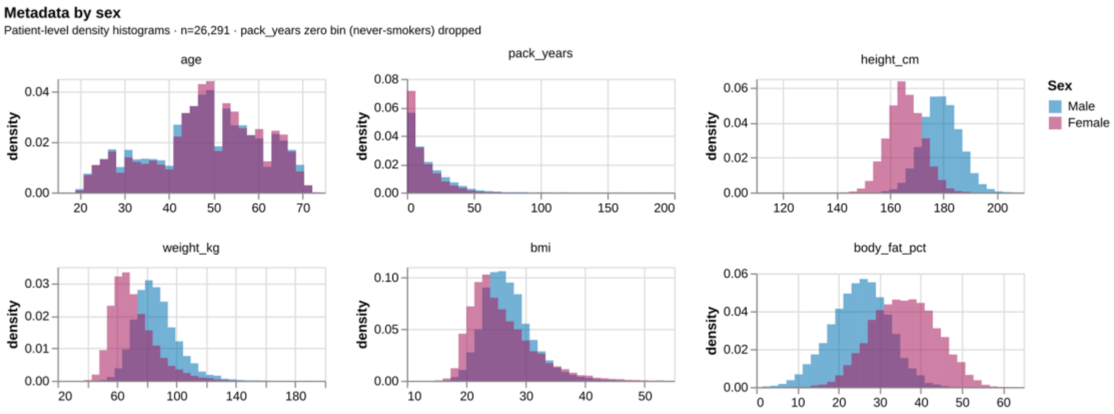


*Supplementary Figure 6:* ***Distribution of each continuous metadata predictor stratified by biological sex.***

*Companion view to [Table 1](#).*

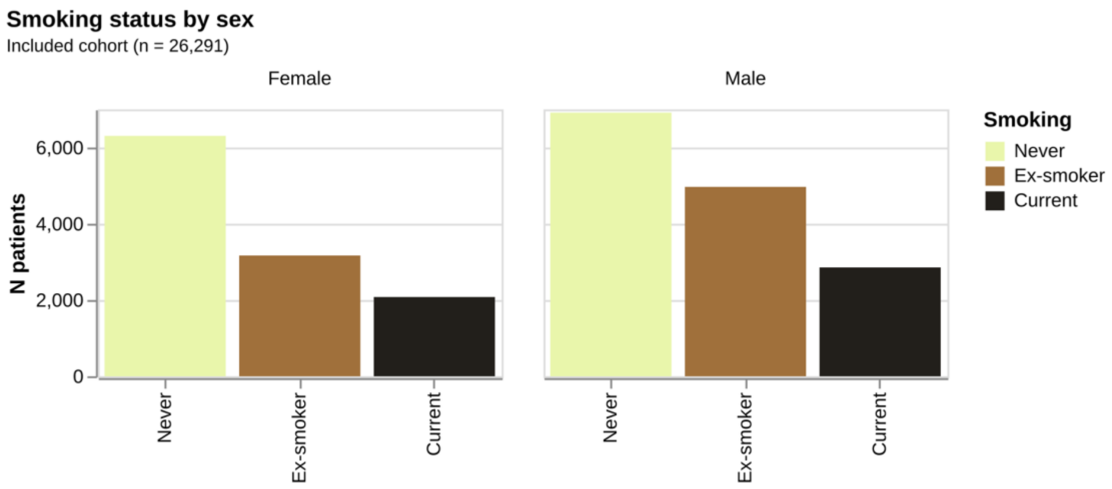


*Supplementary Figure 7:* ***Smoking-status composition stratified by biological sex.*** *Companion view to Table 1.*

### S2 Shape-model variance and convergence

#### S2.1 Scree plot

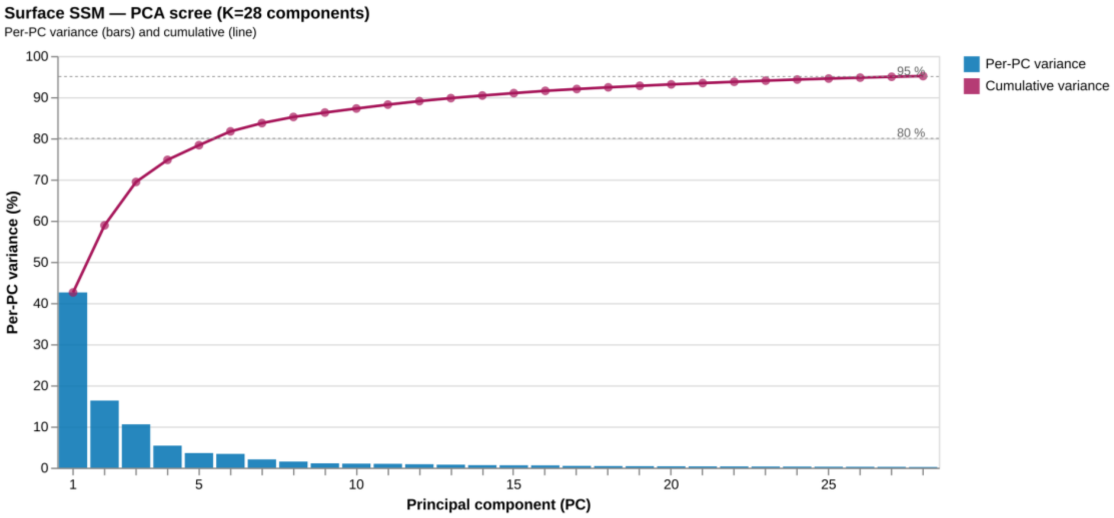


*Supplementary Figure 8:* ***Per-mode and cumulative explained variance.*** *Variance retained at the truncation threshold (95%) is annotated; 28 modes were carried forward into all downstream PC-score analyses.*

#### S2.2 Generalized Procrustes Analysis convergence

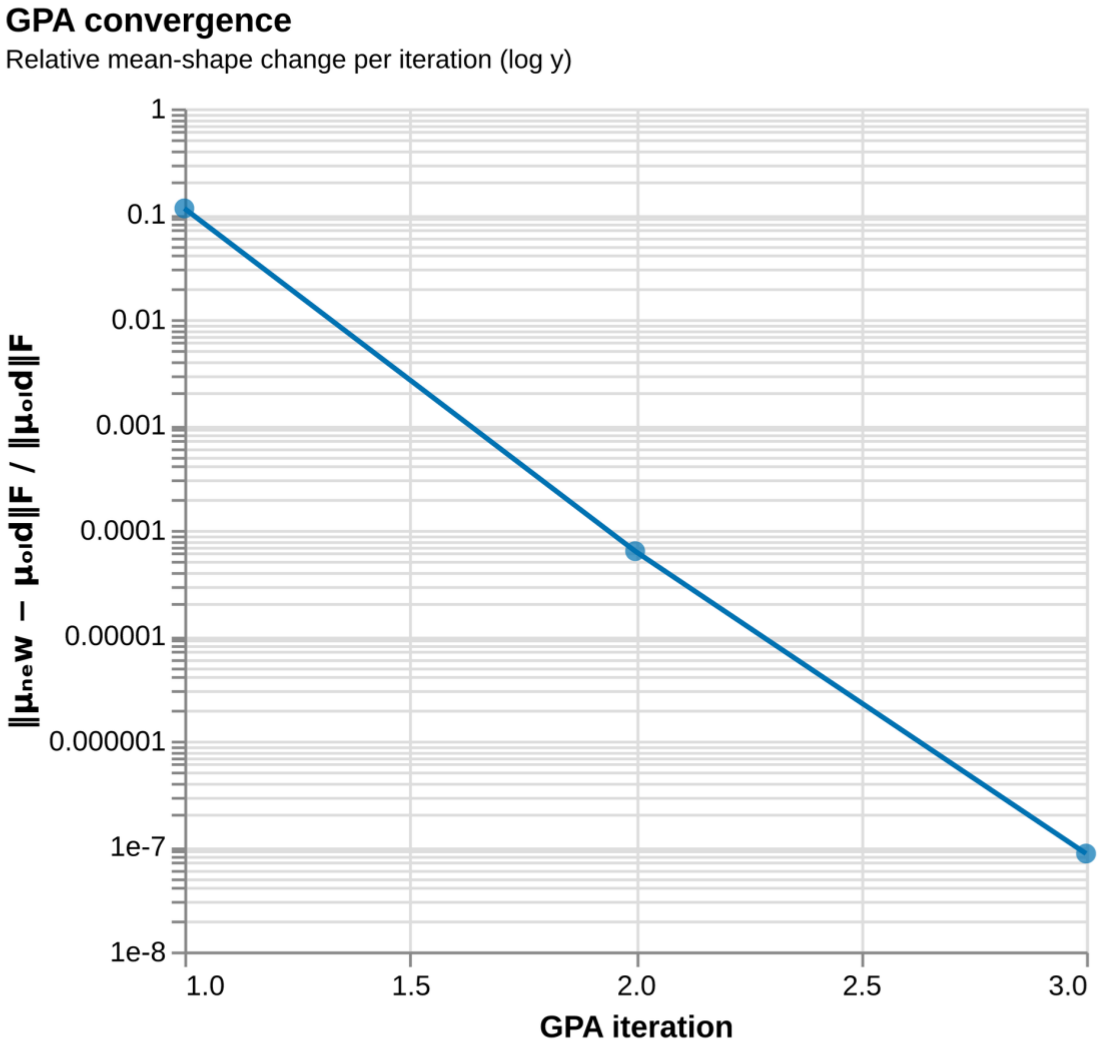


*Supplementary Figure 9:* ***GPA convergence trace.*** *Mean-shape change per iteration of the Generalized Procrustes Analysis described in Methods §2.7.3, with the convergence threshold marked.*

#### S3 Descriptor-level associations

##### S3.1 Adjusted OLS effect maps

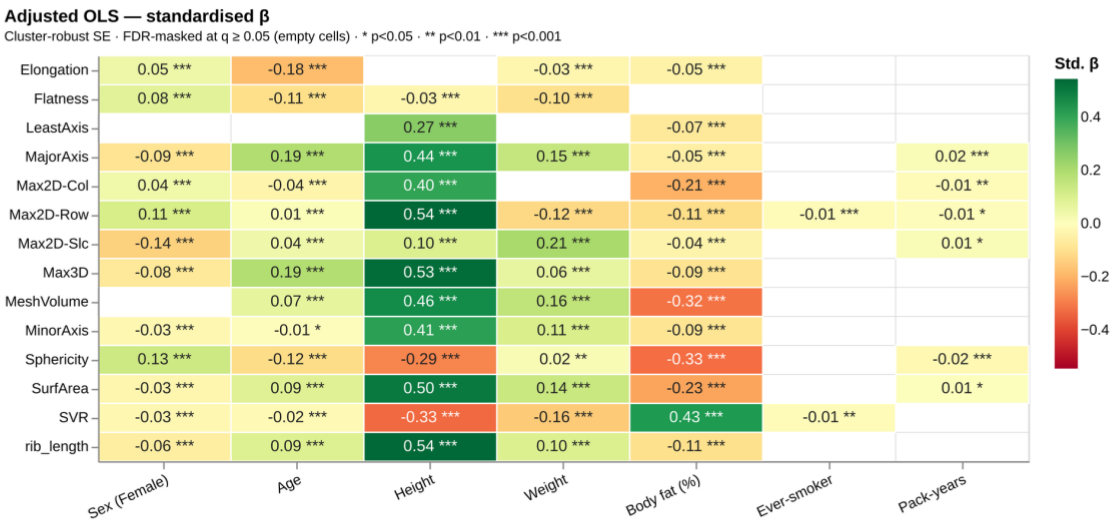


*(a) Panel A – standardised β.*

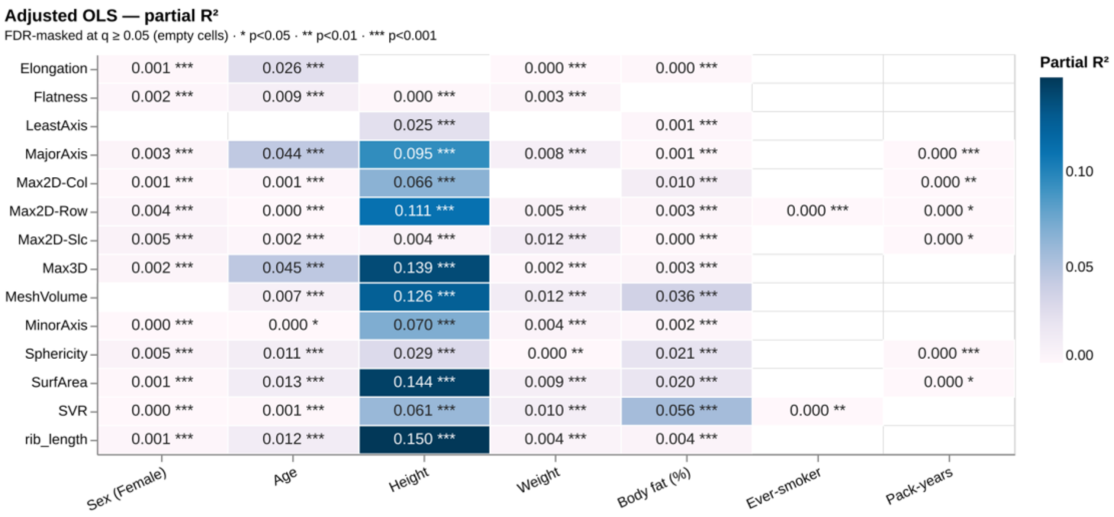


*(b) Panel B – Frisch–Waugh–Lovell partial $R^2$.*

*Supplementary Figure 10:* ***Adjusted (multivariable) OLS effect maps. (A)*** *Standardised β and* ***(B)*** *Frisch–Waugh–Lovell partial $R^2$ per (shape descriptor × predictor) pair from the single adjusted model (Methods §2.8.2) – the mutually-adjusted counterpart to the main-text targeted maps. FDR-masked at $q \geq 0.05$; cluster-robust standard* *errors at the participant level.*

##### S3.2 Adjusted OLS standardised β forest plot

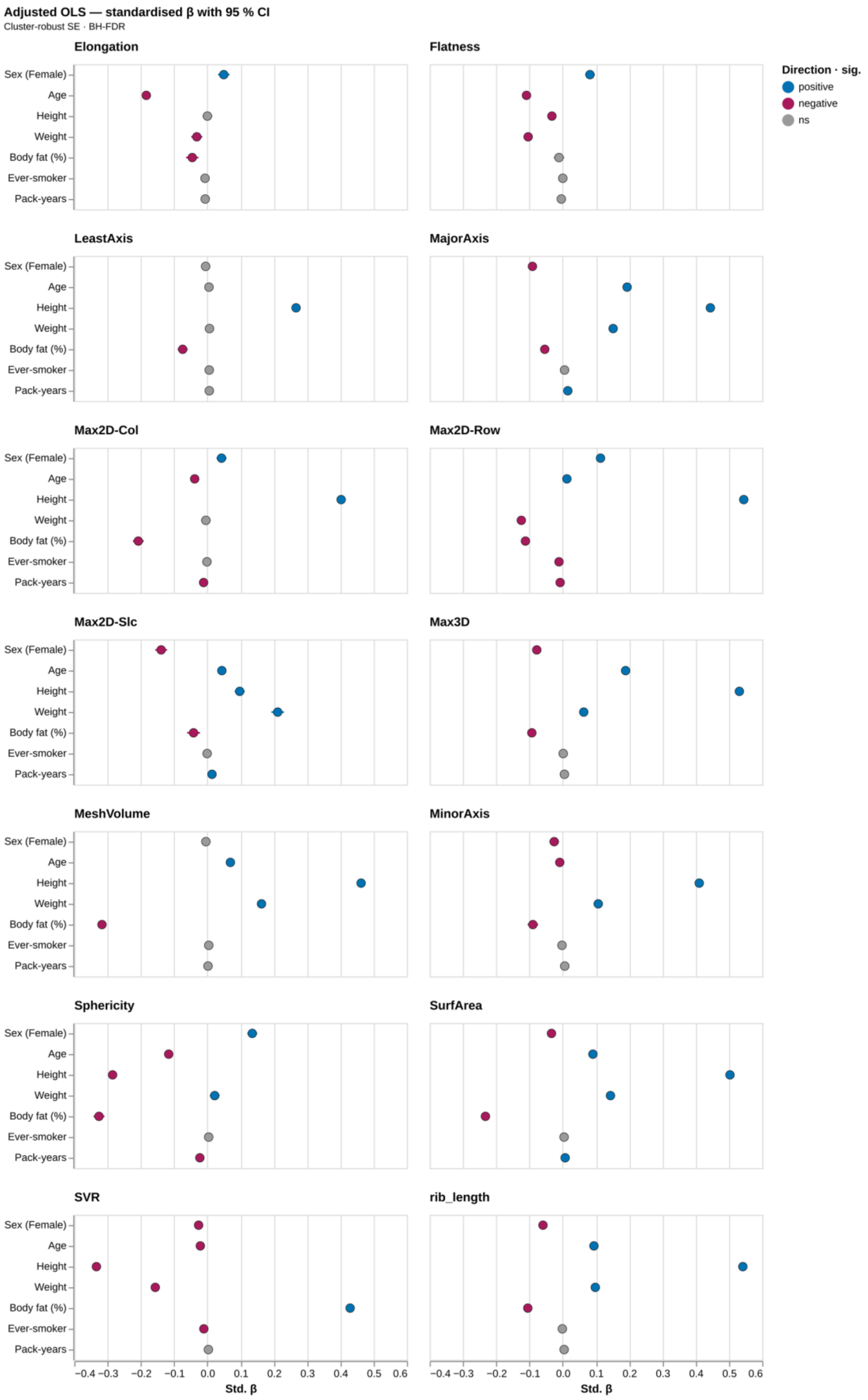


*Supplementary Figure 11:* ***Adjusted OLS standardised β forest plot.*** *95% confidence intervals on the standardised slope per (predictor, descriptor) pair from the adjusted model.*

#### S3.3 Unadjusted (marginal) standardised β heatmap

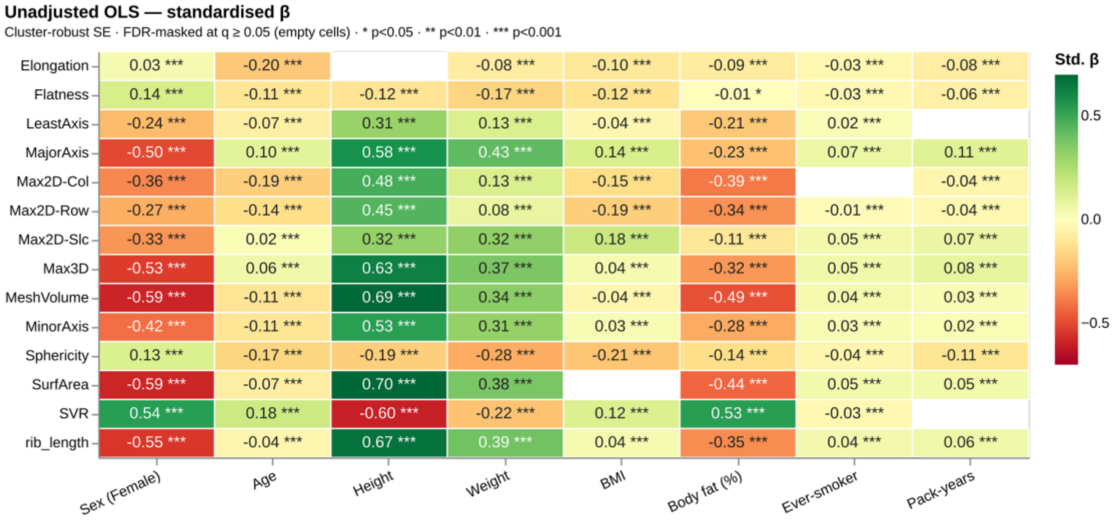


*Supplementary Figure 12:* ***Unadjusted (marginal) standardised β heatmap.*** *Per-predictor marginal slope for each (shape descriptor × predictor) pair, including BMI (unadjusted layer only) – the total-association counterpart to the adjusted and targeted maps.*

#### S3.4 Per-descriptor rib-position maps

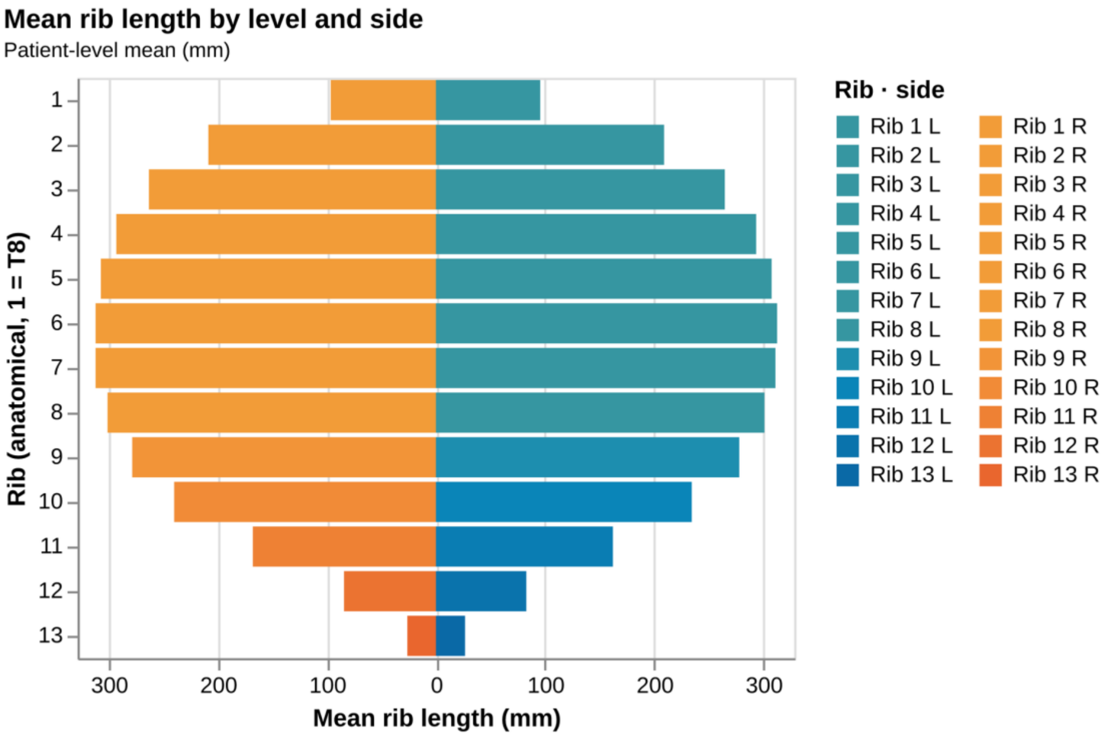

*Supplementary Figure 13:* ***Mean rib length by anatomical level and side.***

### S4 PC score associations

#### S4.1 Adjusted PC association map

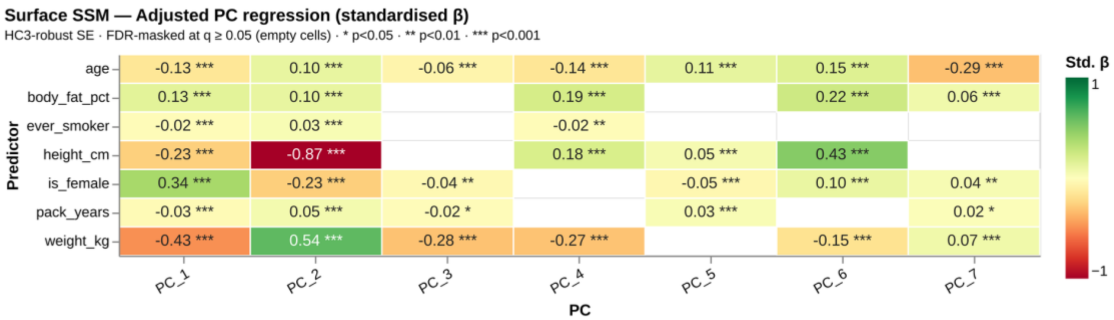


*Supplementary Figure 14:* ***Adjusted (multivariable) associations with the leading PC scores.*** *Standardised β from the per-PC adjusted (HC3) OLS against the seven covariates – the mutually-adjusted counterpart to the main-text targeted map.*

#### S4.2 PC β-vector field

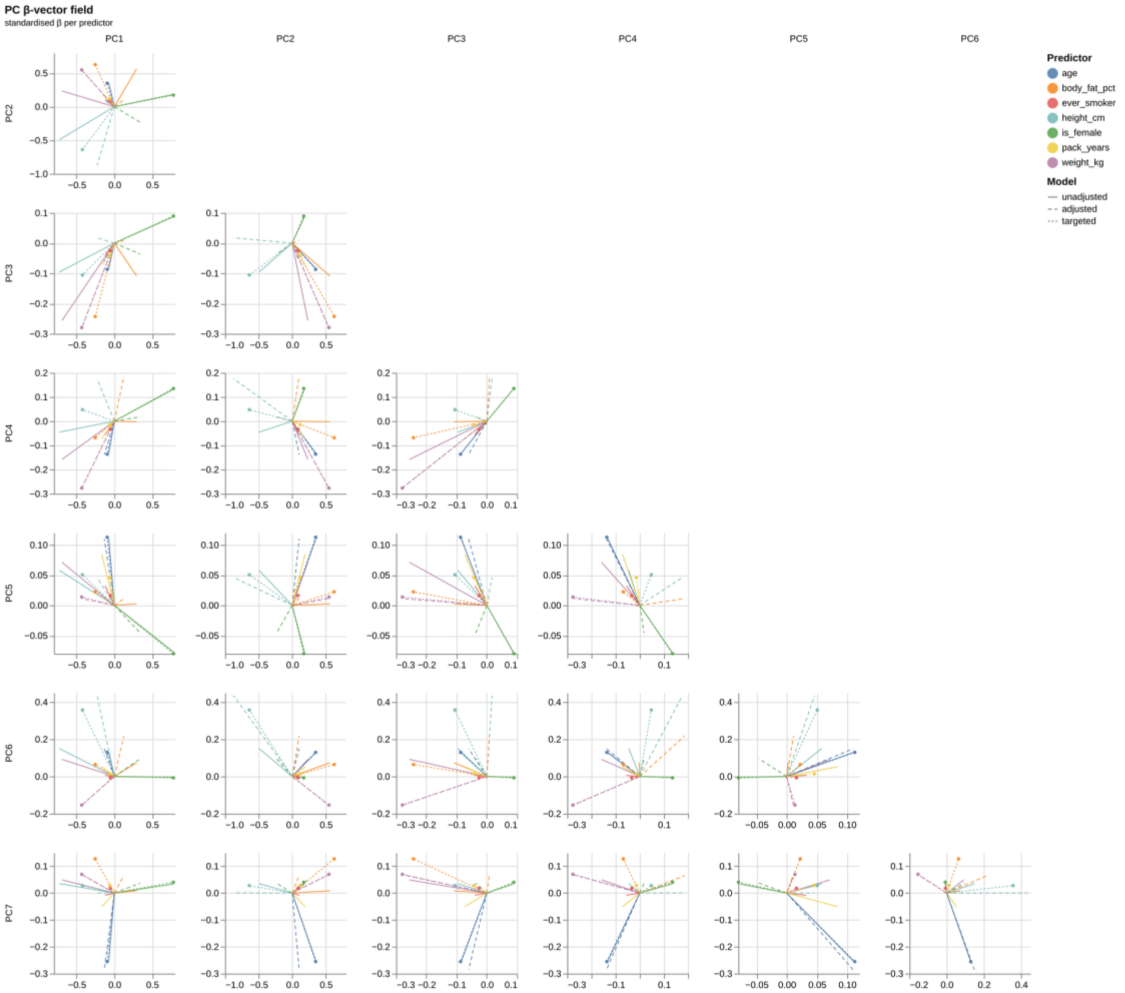


*Supplementary Figure 15:* ***Per-PC β-vector field.*** *Each predictor's standardised β as an arrow in PC space; solid arrows are the unadjusted (marginal) effect, dashed arrows the adjusted effect. Complements the magnitude ranking of the heatmaps with effect direction.*

#### S4.3 Marginal and adjusted PC sex pair-plots

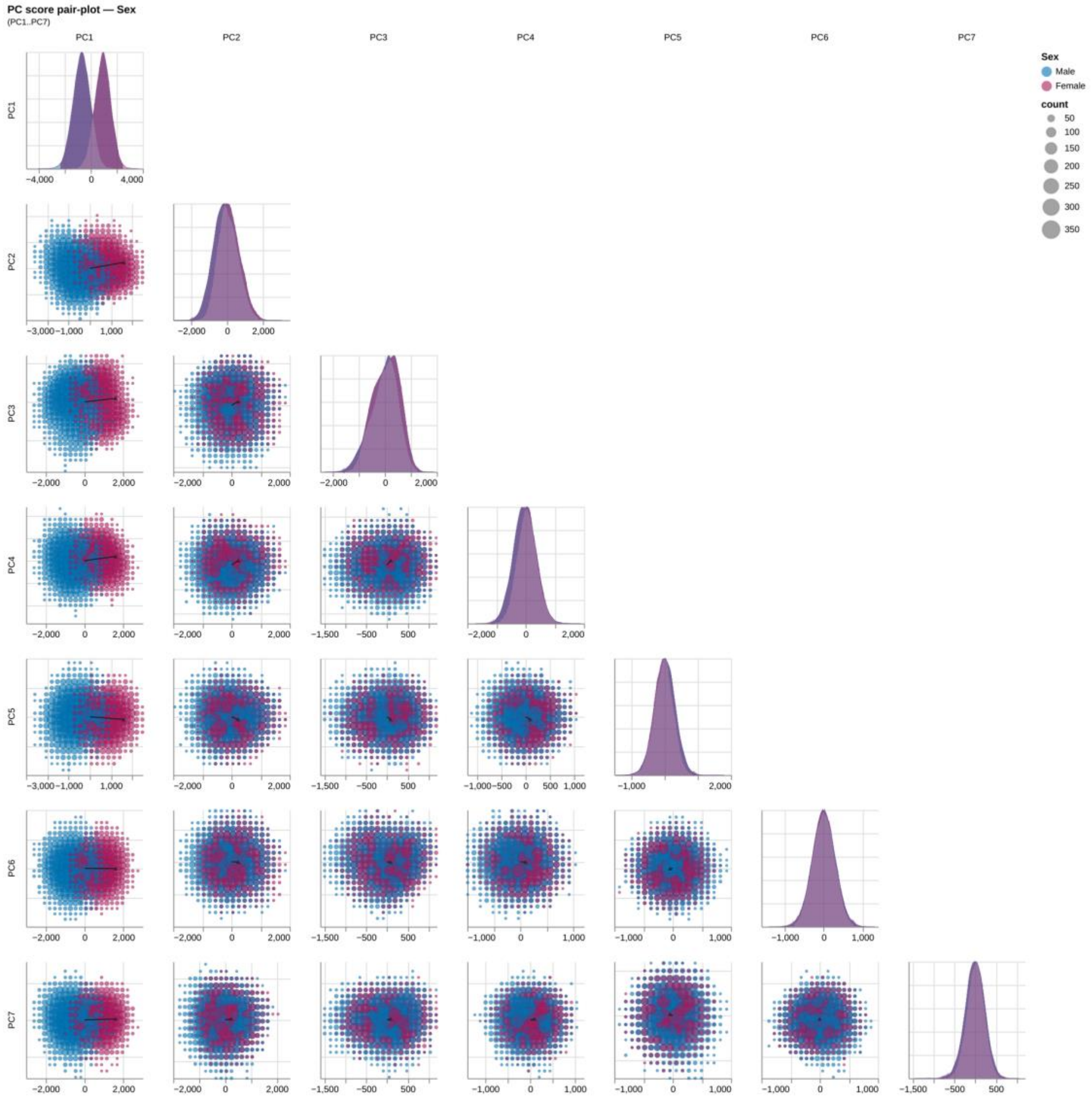


*Supplementary Figure 16:* ***PC score pair-plot by sex (marginal).*** *The unadjusted by-sex pair-plot; companion to the targeted view in the main text.*

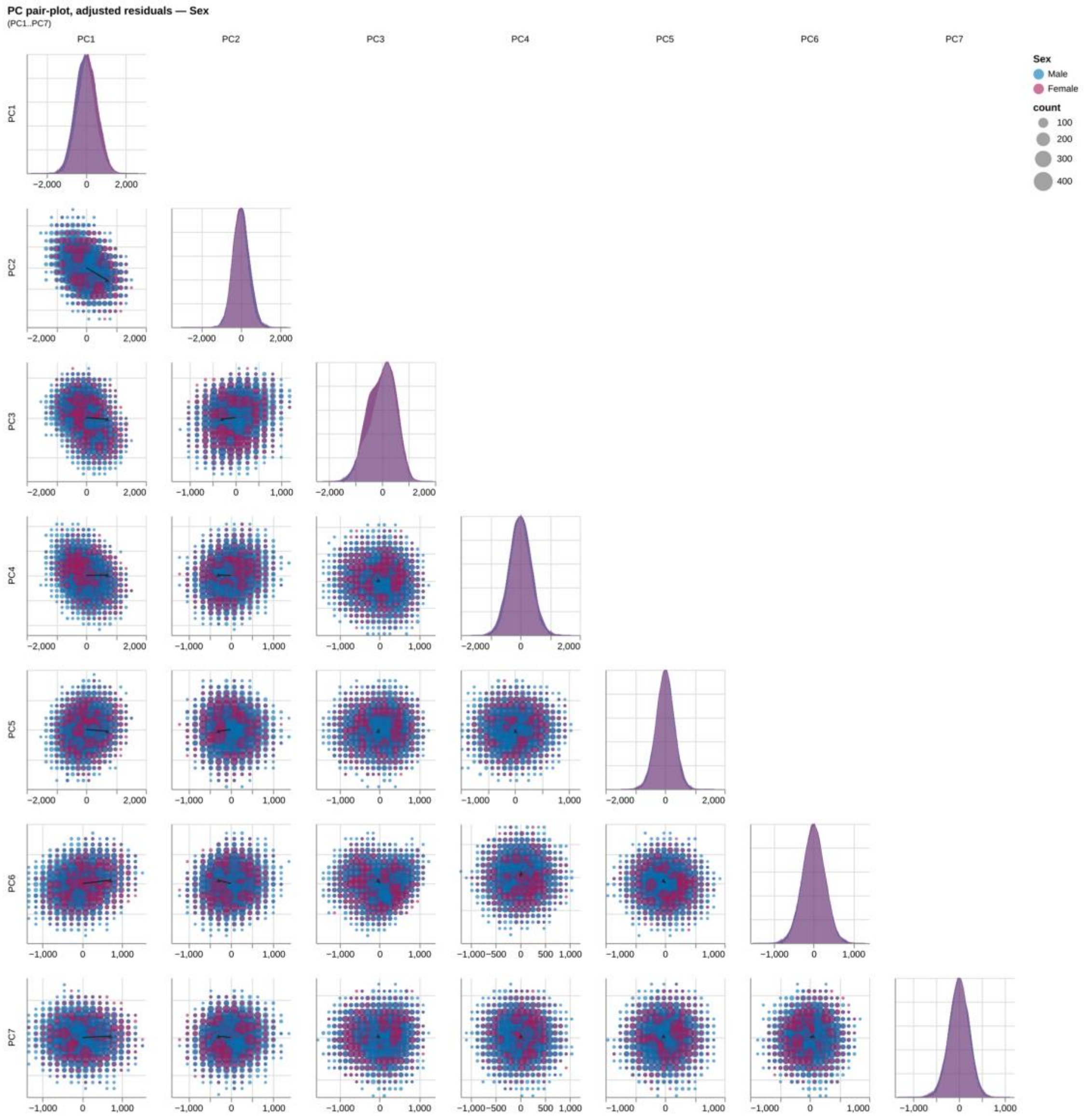


*Supplementary Figure 17:* ***PC score pair-plot by sex (adjusted).*** *PC scores residualised on all covariates except sex; the PC1 separation shrinks relative to the marginal and targeted views, visualising the body-composition mediation of the sex difference.*

#### S4.4 PC scores pair-plot by body fat

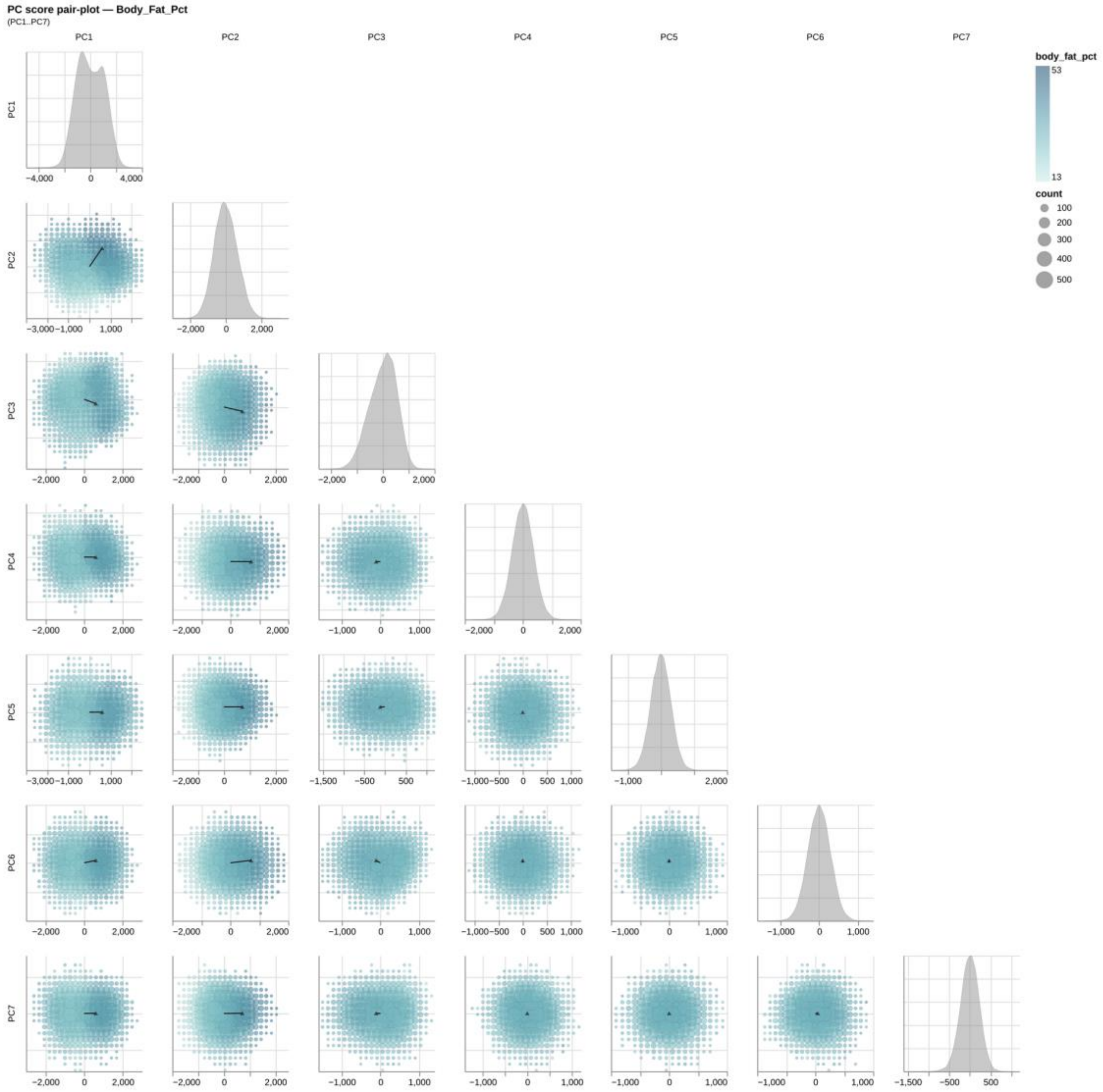


*Supplementary Figure 18:* ***PC score pair-plot coloured by body-fat percentage.*** *Off-diagonal bins coloured by mean body-fat percentage, showing the body-composition gradient aligning with the leading modes.*

#### S4.5 Per-rib PC loadings

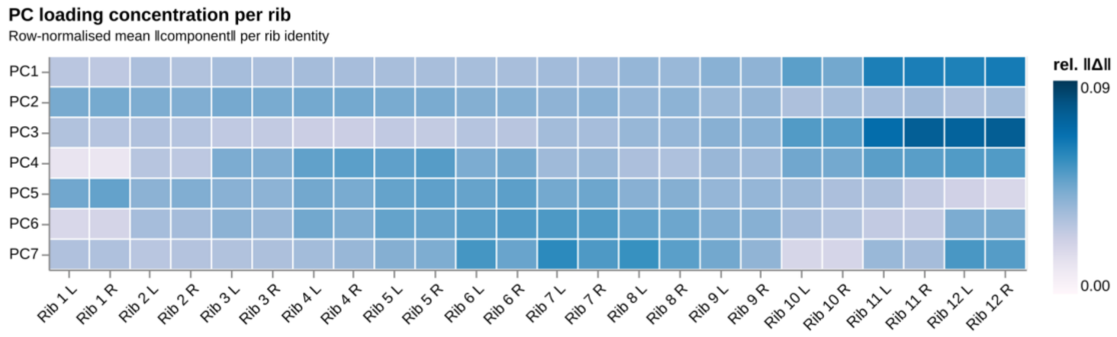


*Supplementary Figure 19:* ***Per-rib loadings of the leading principal components.*** *Loadings of each PC onto each of the 24 rib positions.*

##### S4.6 PC scores pair-plot by smoking status

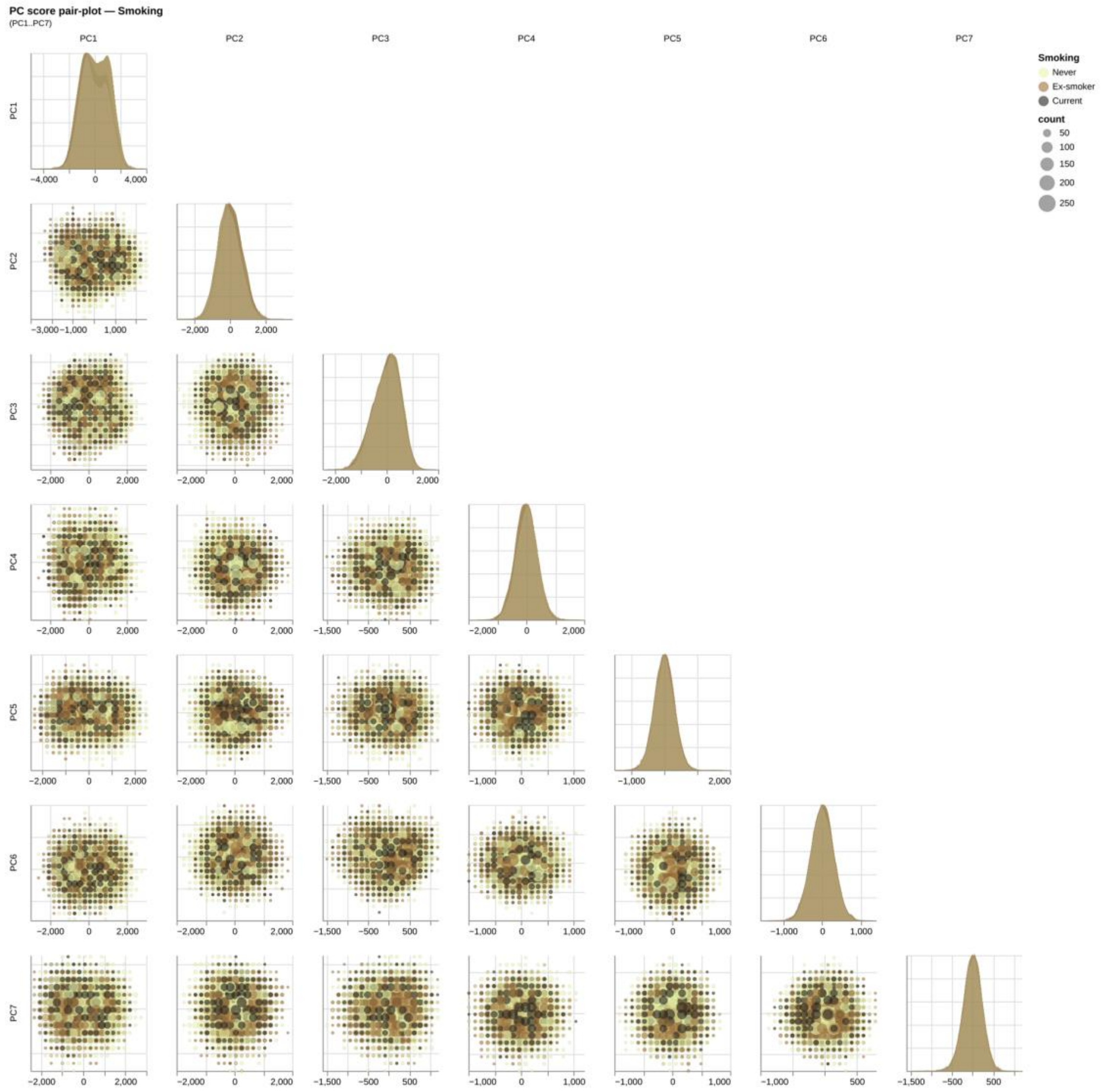


*Supplementary Figure 20:* ***PC score pair-plot stratified by smoking status.*** *Companion view to the by-sex pair-plot in the main text. Smoking is shown here as the descriptive three-level variable (Never / Ex-smoker / Current); the association models use the binary ever-smoker contrast. Marginal density panels show the per-PC distribution per smoking-status group.*

#### S5 Anatomical interpretation of principal modes

##### S5.1 PC-anchored anatomical maps (PC4 onwards)

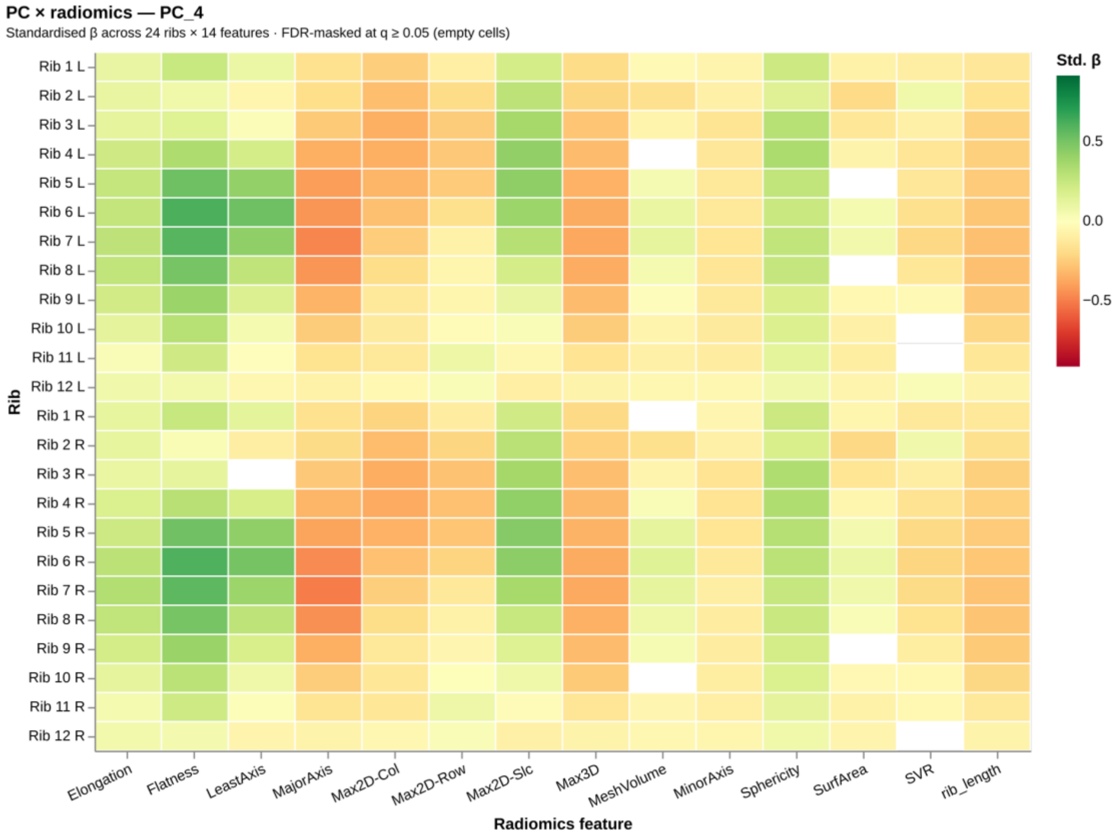
PC × radiomics — PC_4
Standardised β across 24 ribs × 14 features · FDR-masked at q ≥ 0.05 (empty cells)
Rib 1 L
Rib 2 L
Rib 3 L
Rib 4 L
Rib 5 L
Rib 6 L
Rib 7 L
Rib 8 L
Rib 9 L
Rib 10 L
Rib 11 L
Rib 12 L
Rib 1 R
Rib 2 R
Rib 3 R
Rib 4 R
Rib 5 R
Rib 6 R
Rib 7 R
Rib 8 R
Rib 9 R
Rib 10 R
Rib 11 R
Rib 12 R
Rib
Elongation
Flatness
LeastAxis
MajorAxis
Max2D-Col
Max2D-Row
Max2D-Slc
Max3D
MeshVolume
MinorAxis
Sphericity
SurfArea
SVR
rib_length
Radiomics feature
Std. β
0.5
0.0
−0.5

*(a) PC4*

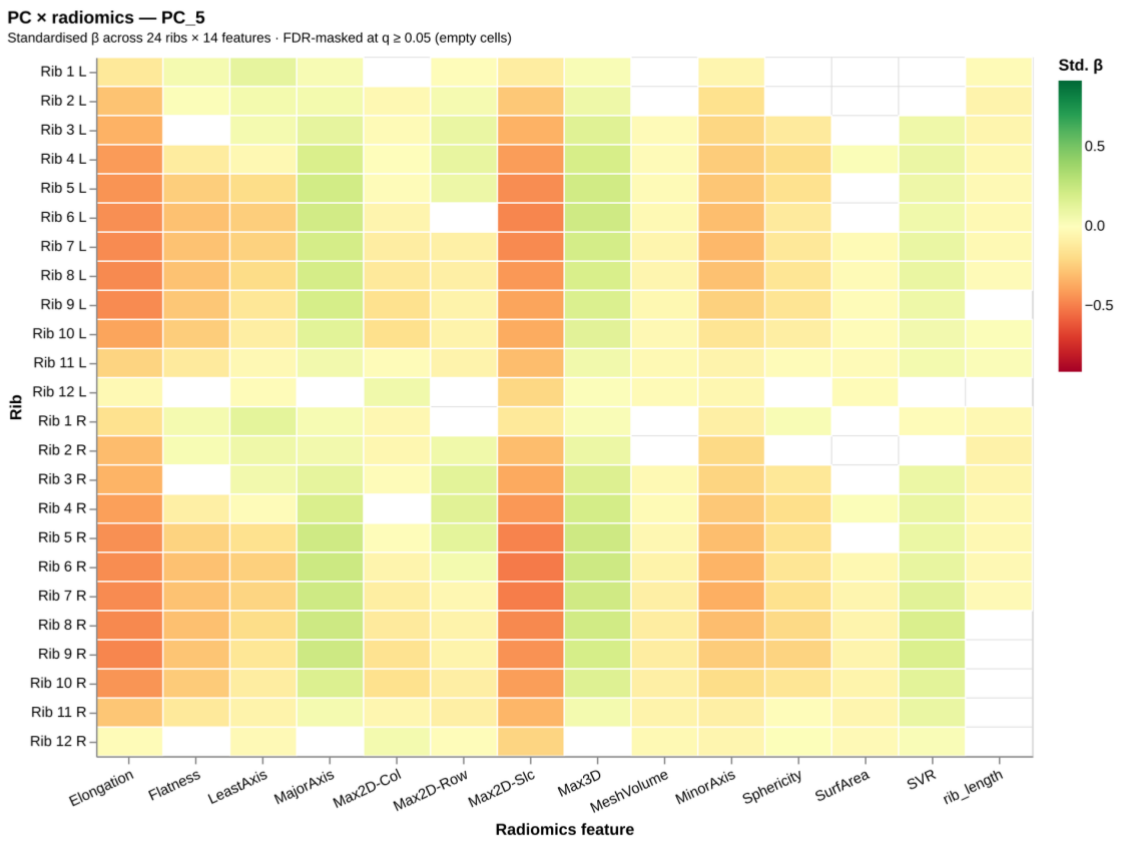
PC × radiomics — PC_5
Standardised β across 24 ribs × 14 features · FDR-masked at q ≥ 0.05 (empty cells)
Rib 1 L
Rib 2 L
Rib 3 L
Rib 4 L
Rib 5 L
Rib 6 L
Rib 7 L
Rib 8 L
Rib 9 L
Rib 10 L
Rib 11 L
Rib 12 L
Rib 1 R
Rib 2 R
Rib 3 R
Rib 4 R
Rib 5 R
Rib 6 R
Rib 7 R
Rib 8 R
Rib 9 R
Rib 10 R
Rib 11 R
Rib 12 R
Rib
Elongation
Flatness
LeastAxis
MajorAxis
Max2D-Col
Max2D-Row
Max2D-Slc
Max3D
MeshVolume
MinorAxis
Sphericity
SurfArea
SVR
rib_length
Radiomics feature
Std. β
0.5
0.0
−0.5

*(b) PC5*

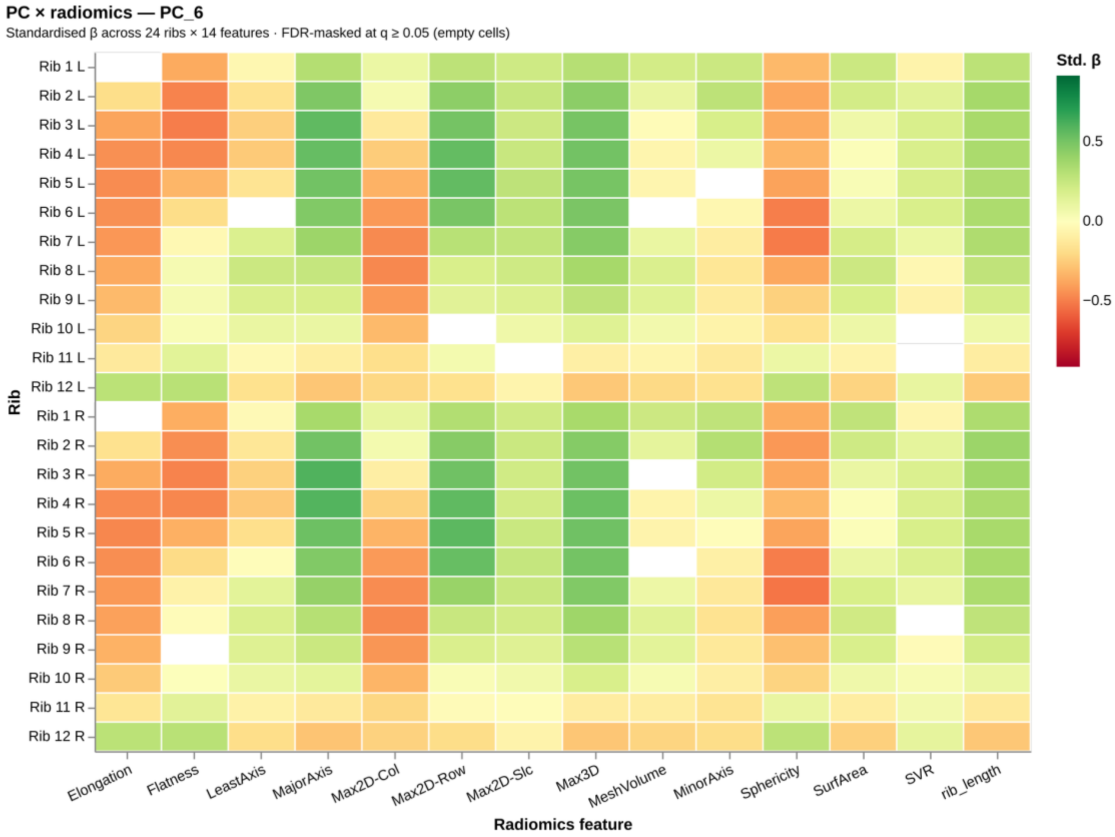
PC × radiomics — PC_6
Standardised β across 24 ribs × 14 features · FDR-masked at q ≥ 0.05 (empty cells)
Std. β
0.5
0.0
−0.5
Rib
Rib 1 L
Rib 2 L
Rib 3 L
Rib 4 L
Rib 5 L
Rib 6 L
Rib 7 L
Rib 8 L
Rib 9 L
Rib 10 L
Rib 11 L
Rib 12 L
Rib 1 R
Rib 2 R
Rib 3 R
Rib 4 R
Rib 5 R
Rib 6 R
Rib 7 R
Rib 8 R
Rib 9 R
Rib 10 R
Rib 11 R
Rib 12 R
Elongation
Flatness
LeastAxis
MajorAxis
Max2D-Col
Max2D-Row
Max2D-Slc
Max3D
MeshVolume
MinorAxis
Sphericity
SurfArea
SVR
rib_length
Radiomics feature

*(c) PC6*

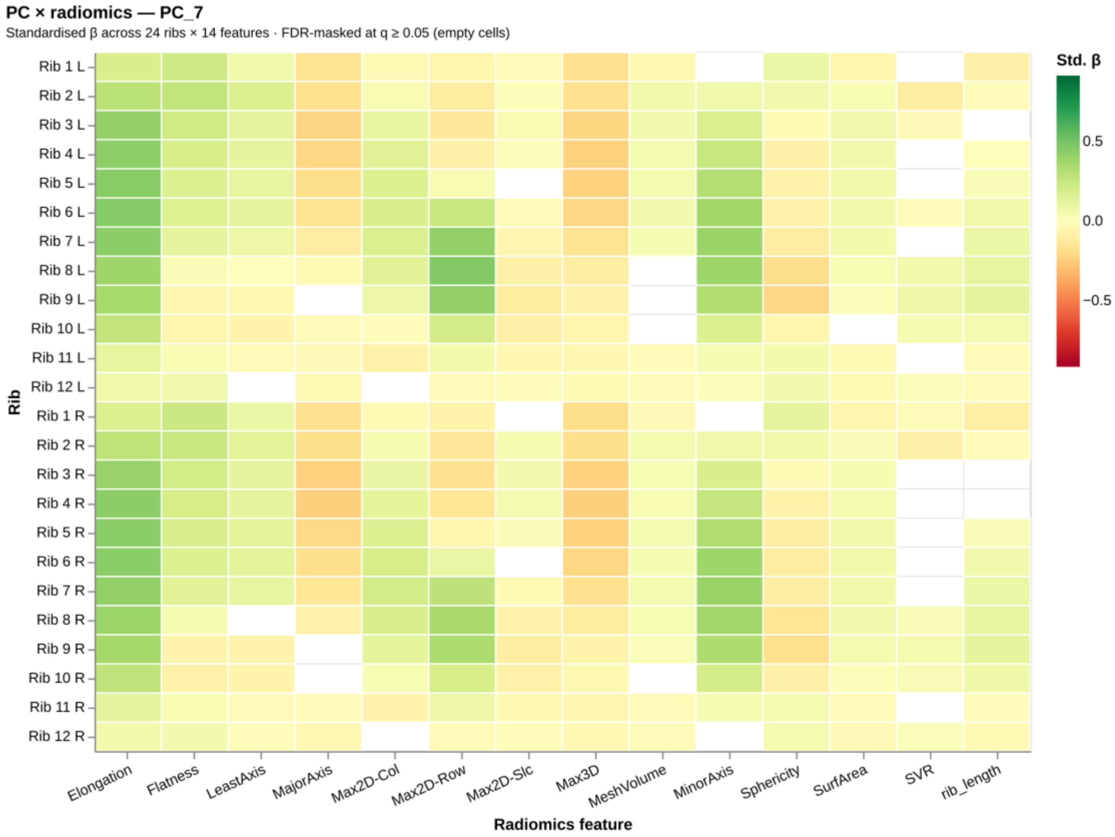


(d) PC7

*Supplementary Figure 21:* ***Per-PC anatomical cross-walk (PC4 onwards).*** *Standardised slope of every (rib position × shape descriptor) pair on each PC score (cohort-z-scored), rendered as 24-row × 14-column heatmaps. Print shows PC4–PC7; the interactive supplement browses all 28 modes. PC1–PC3 are in the main-text anatomical maps.*

#### S5.2 Descriptor-anchored anatomical maps

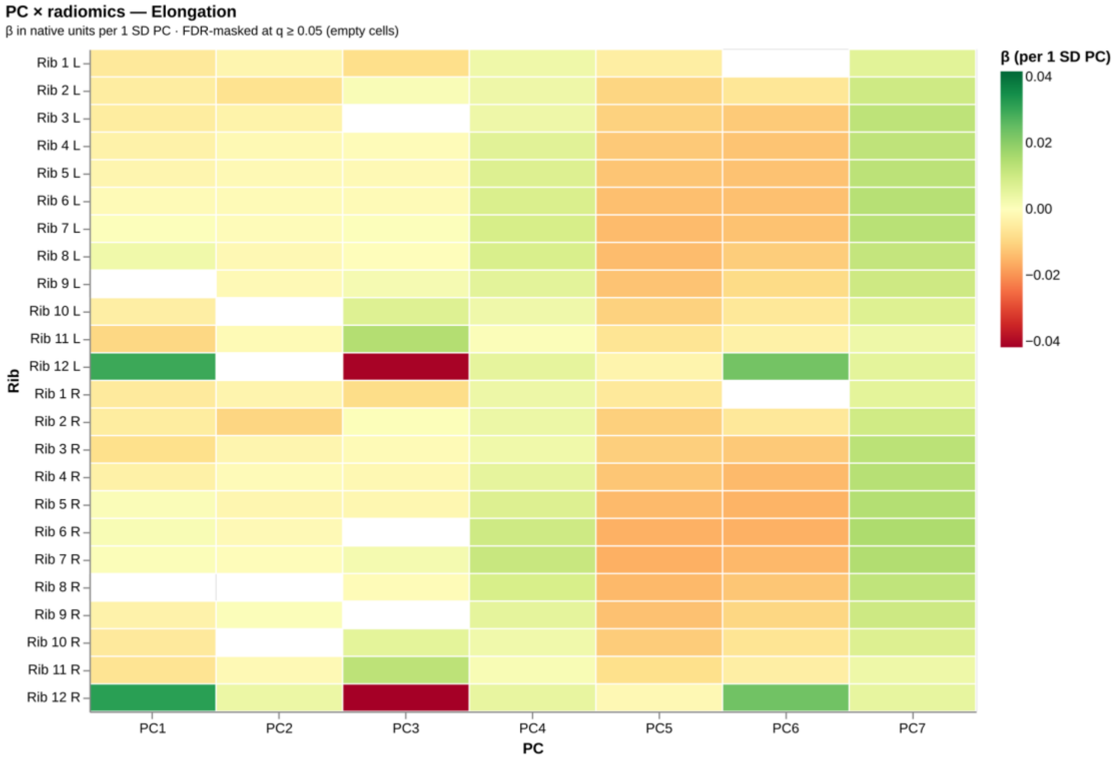


*(a) Elongation*

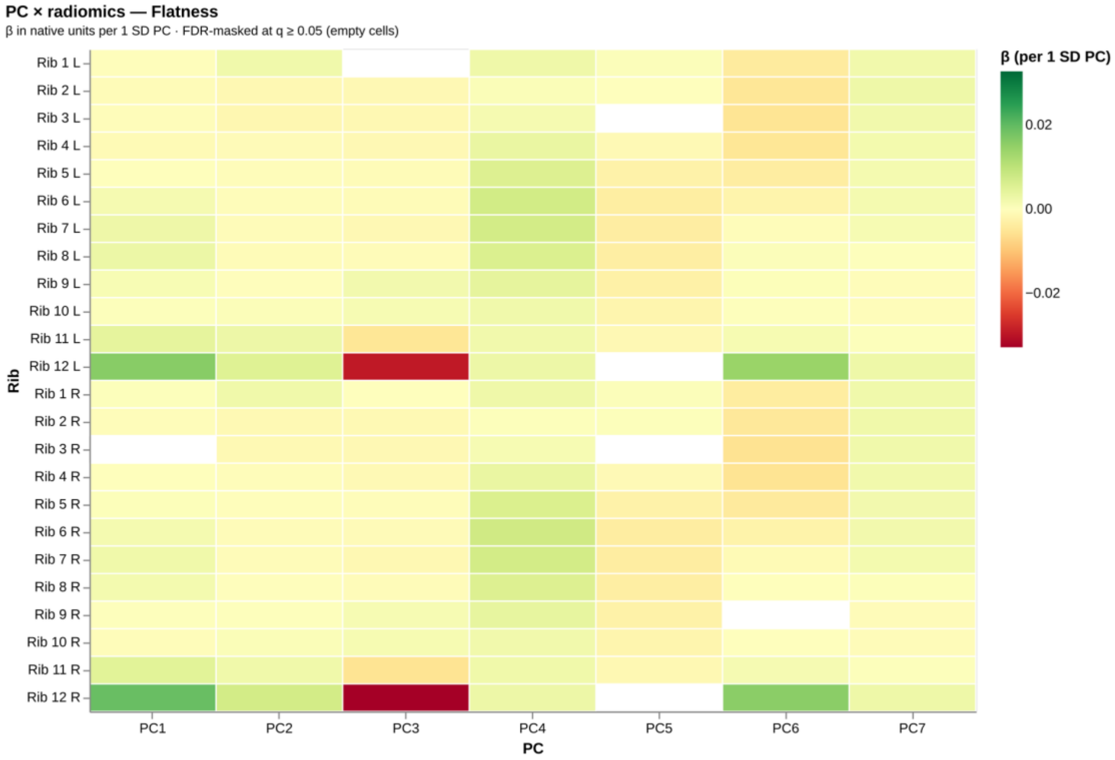

PC × radiomics — Flatness
β in native units per 1 SD PC · FDR-masked at q ≥ 0.05 (empty cells)
β (per 1 SD PC)
0.02
0.00
−0.02
Rib 1 L
Rib 2 L
Rib 3 L
Rib 4 L
Rib 5 L
Rib 6 L
Rib 7 L
Rib 8 L
Rib 9 L
Rib 10 L
Rib 11 L
Rib 12 L
Rib 1 R
Rib 2 R
Rib 3 R
Rib 4 R
Rib 5 R
Rib 6 R
Rib 7 R
Rib 8 R
Rib 9 R
Rib 10 R
Rib 11 R
Rib 12 R
Rib
PC1
PC2
PC3
PC4
PC5
PC6
PC7
PC


*(b) Flatness*

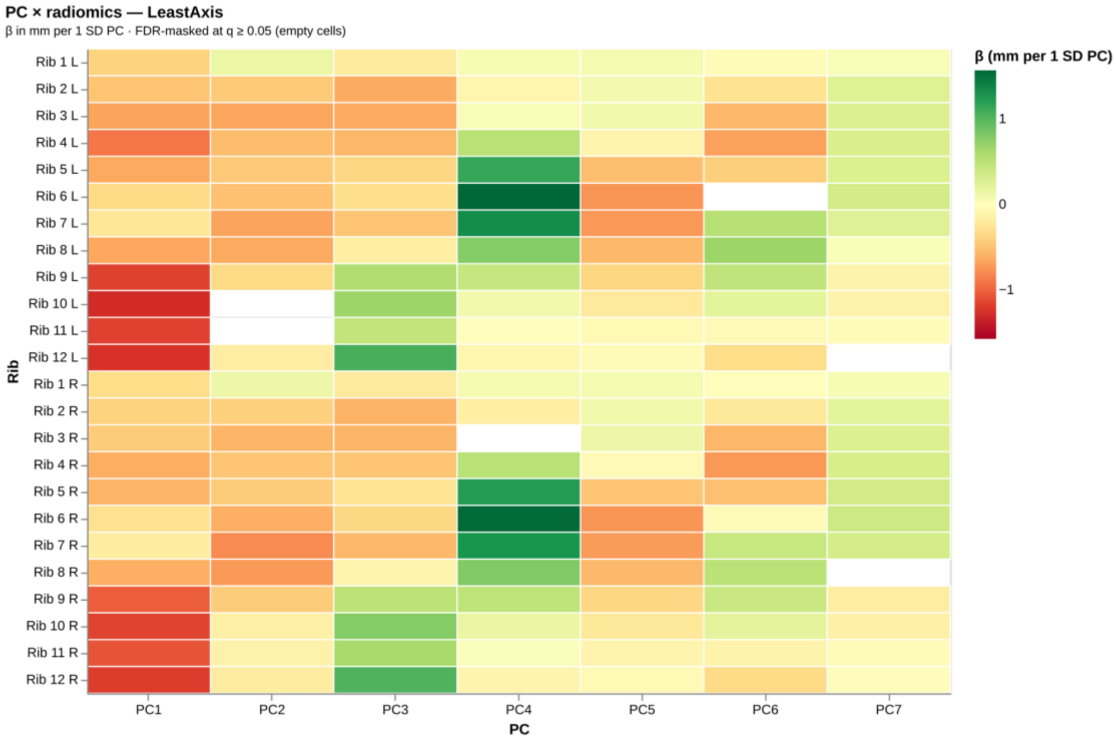

PC × radiomics — LeastAxis
β in mm per 1 SD PC · FDR-masked at q ≥ 0.05 (empty cells)
β (mm per 1 SD PC)
1
0
−1
Rib 1 L
Rib 2 L
Rib 3 L
Rib 4 L
Rib 5 L
Rib 6 L
Rib 7 L
Rib 8 L
Rib 9 L
Rib 10 L
Rib 11 L
Rib 12 L
Rib 1 R
Rib 2 R
Rib 3 R
Rib 4 R
Rib 5 R
Rib 6 R
Rib 7 R
Rib 8 R
Rib 9 R
Rib 10 R
Rib 11 R
Rib 12 R
Rib
PC1
PC2
PC3
PC4
PC5
PC6
PC7
PC

*(c) LeastAxis*

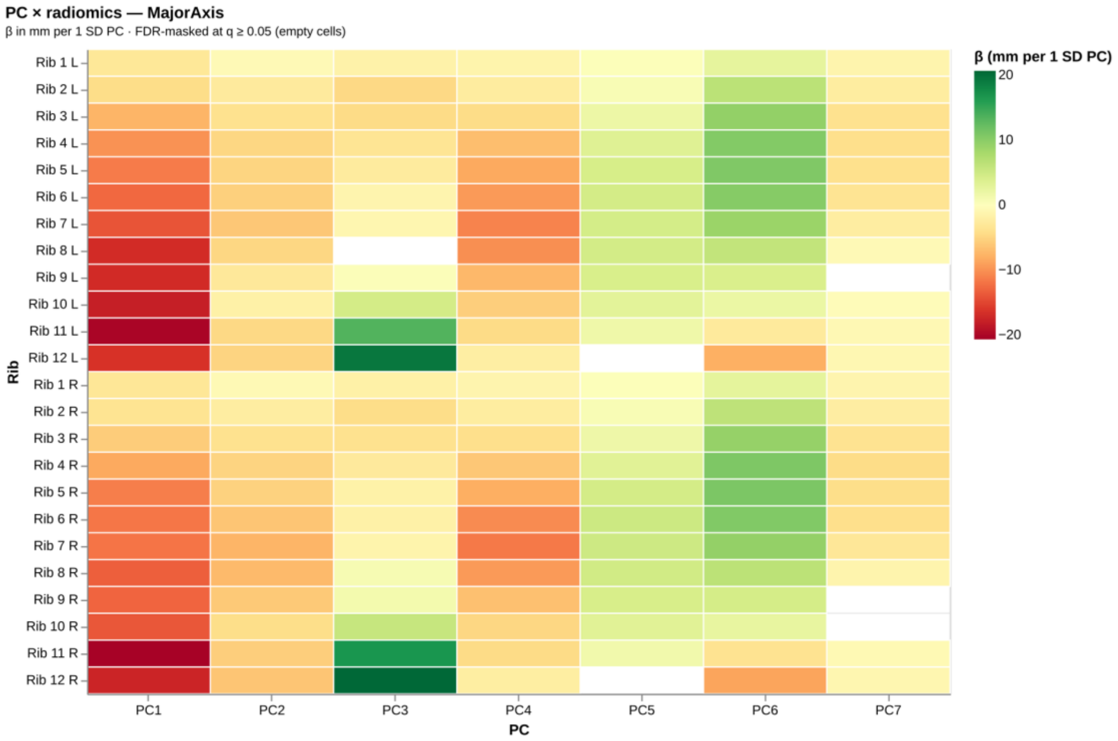


*(d) MajorAxis*

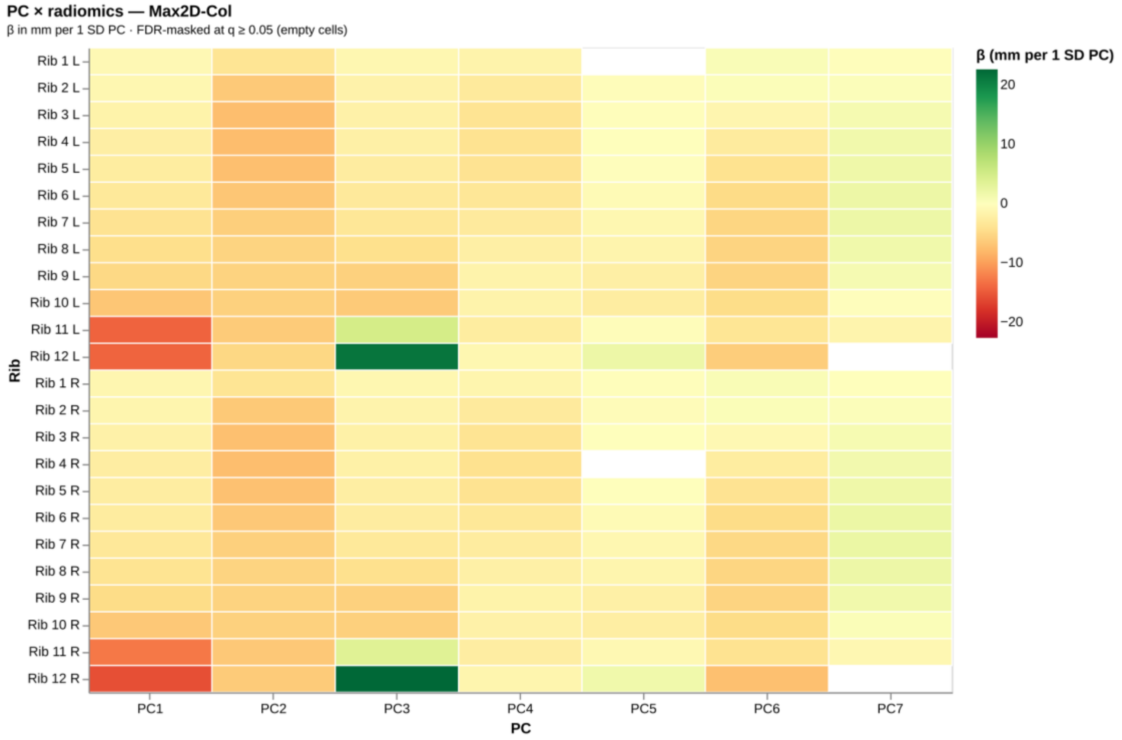
PC × radiomics — Max2D-Col
β in mm per 1 SD PC · FDR-masked at q ≥ 0.05 (empty cells)
Rib 1 L
Rib 2 L
Rib 3 L
Rib 4 L
Rib 5 L
Rib 6 L
Rib 7 L
Rib 8 L
Rib 9 L
Rib 10 L
Rib 11 L
Rib 12 L
Rib 1 R
Rib 2 R
Rib 3 R
Rib 4 R
Rib 5 R
Rib 6 R
Rib 7 R
Rib 8 R
Rib 9 R
Rib 10 R
Rib 11 R
Rib 12 R
Rib
PC1
PC2
PC3
PC4
PC5
PC6
PC7
PC
β (mm per 1 SD PC)
20
10
0
−10
−20

*(e) Max2D-Col*

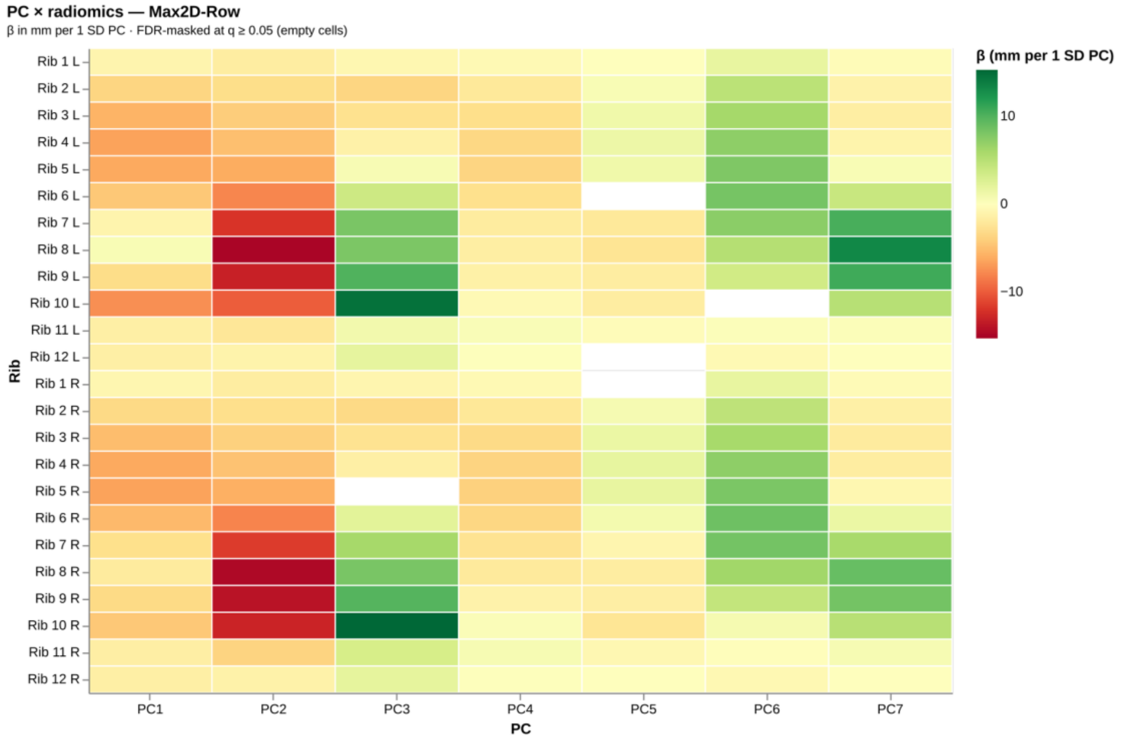
PC × radiomics — Max2D-Row
β in mm per 1 SD PC · FDR-masked at q ≥ 0.05 (empty cells)
Rib 1 L
Rib 2 L
Rib 3 L
Rib 4 L
Rib 5 L
Rib 6 L
Rib 7 L
Rib 8 L
Rib 9 L
Rib 10 L
Rib 11 L
Rib 12 L
Rib 1 R
Rib 2 R
Rib 3 R
Rib 4 R
Rib 5 R
Rib 6 R
Rib 7 R
Rib 8 R
Rib 9 R
Rib 10 R
Rib 11 R
Rib 12 R
Rib
PC1
PC2
PC3
PC4
PC5
PC6
PC7
PC
β (mm per 1 SD PC)
10
0
−10

*(f) Max2D-Row*

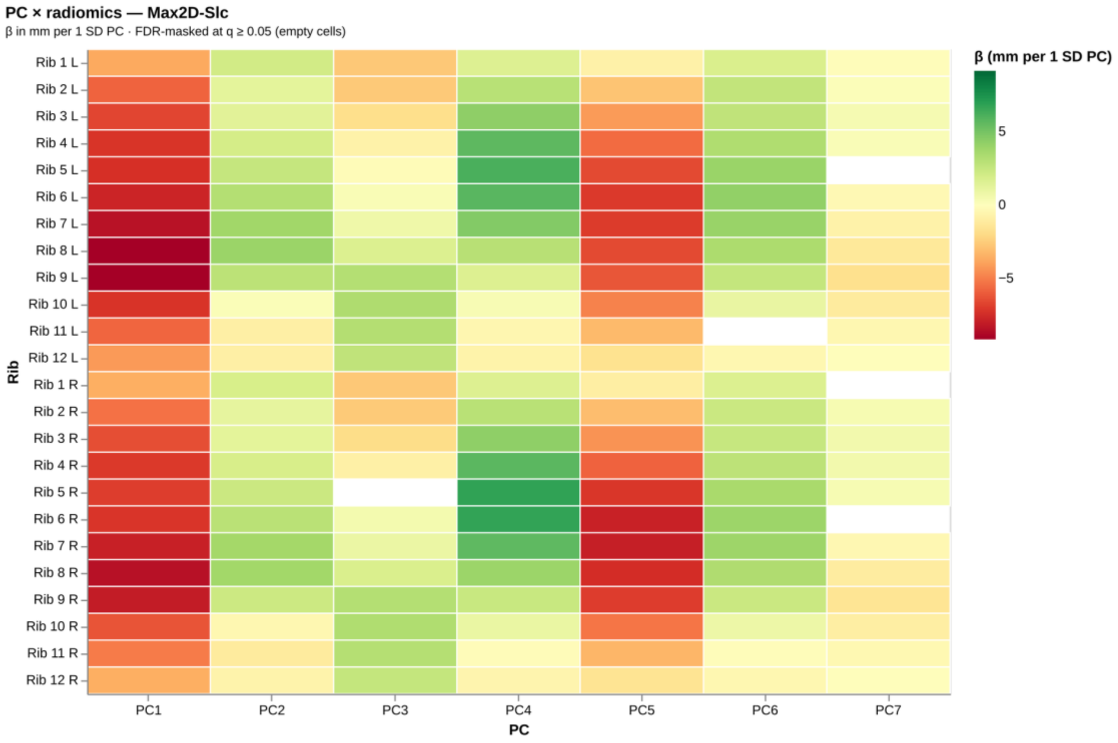


*(g) Max2D-Slc*

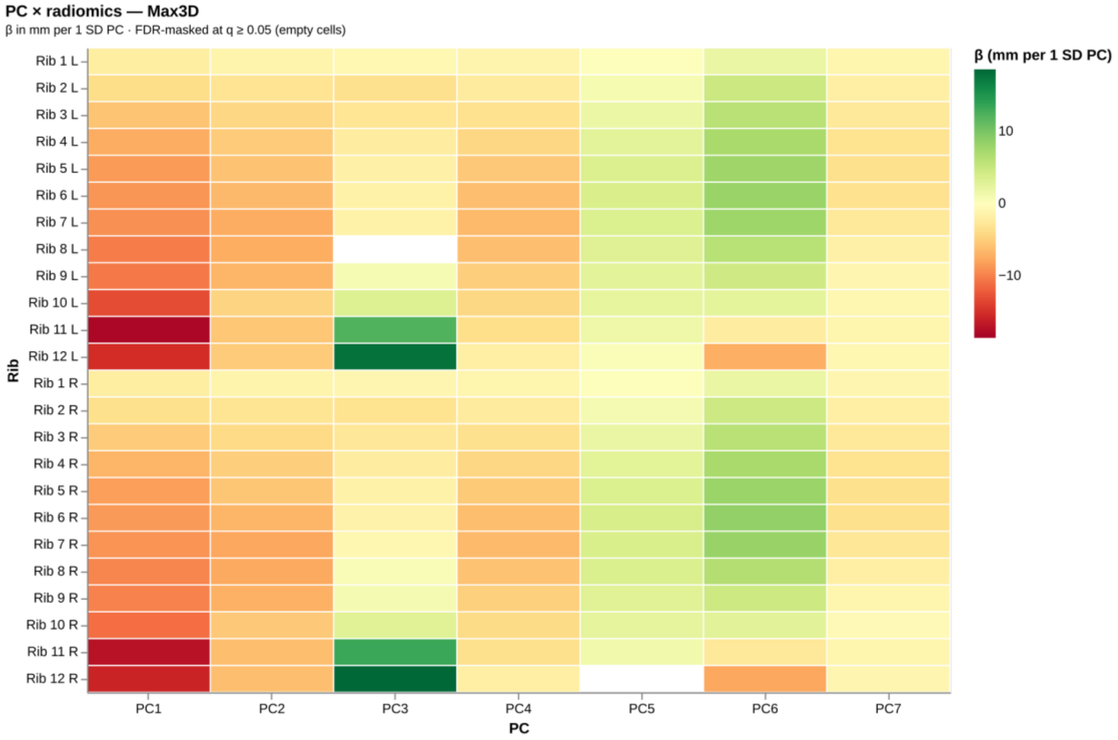

PC × radiomics — Max3D
β in mm per 1 SD PC · FDR-masked at q ≥ 0.05 (empty cells)
β (mm per 1 SD PC)
10
0
−10
Rib 1 L
Rib 2 L
Rib 3 L
Rib 4 L
Rib 5 L
Rib 6 L
Rib 7 L
Rib 8 L
Rib 9 L
Rib 10 L
Rib 11 L
Rib 12 L
Rib 1 R
Rib 2 R
Rib 3 R
Rib 4 R
Rib 5 R
Rib 6 R
Rib 7 R
Rib 8 R
Rib 9 R
Rib 10 R
Rib 11 R
Rib 12 R
Rib
PC1
PC2
PC3
PC4
PC5
PC6
PC7
PC


*(h) Max3D*

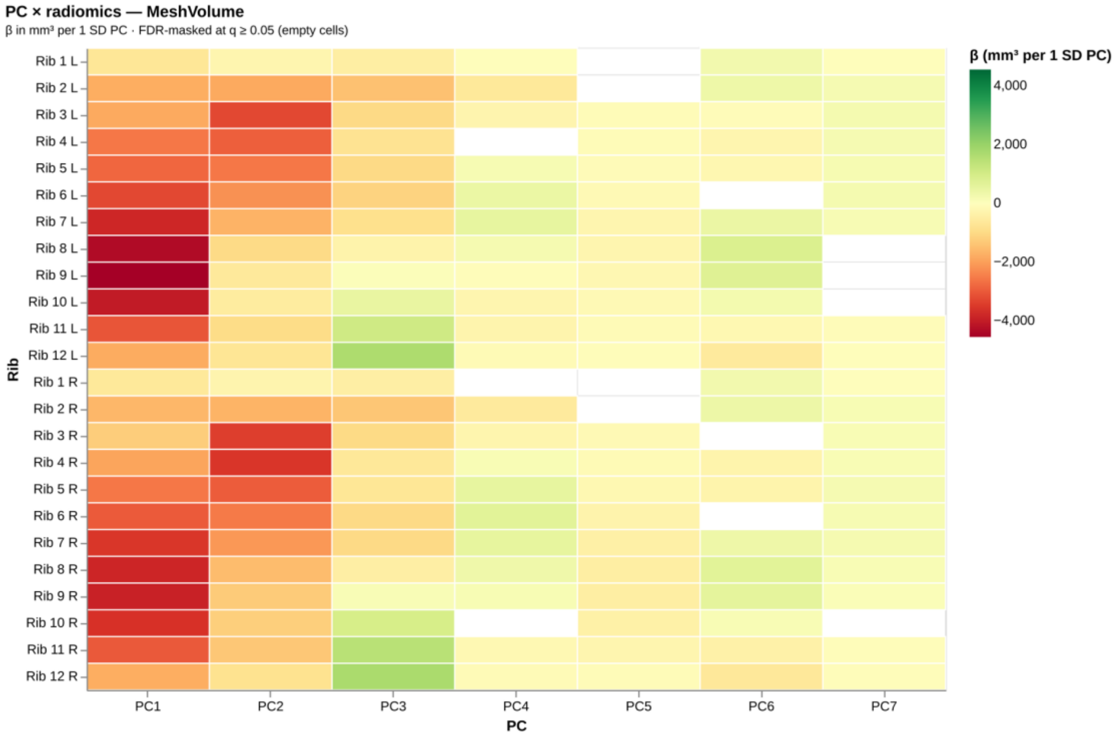

PC × radiomics — MeshVolume
β in mm³ per 1 SD PC · FDR-masked at q ≥ 0.05 (empty cells)
β (mm³ per 1 SD PC)
4,000
2,000
0
−2,000
−4,000
Rib 1 L
Rib 2 L
Rib 3 L
Rib 4 L
Rib 5 L
Rib 6 L
Rib 7 L
Rib 8 L
Rib 9 L
Rib 10 L
Rib 11 L
Rib 12 L
Rib 1 R
Rib 2 R
Rib 3 R
Rib 4 R
Rib 5 R
Rib 6 R
Rib 7 R
Rib 8 R
Rib 9 R
Rib 10 R
Rib 11 R
Rib 12 R
Rib
PC1
PC2
PC3
PC4
PC5
PC6
PC7
PC

*(i) MeshVolume*

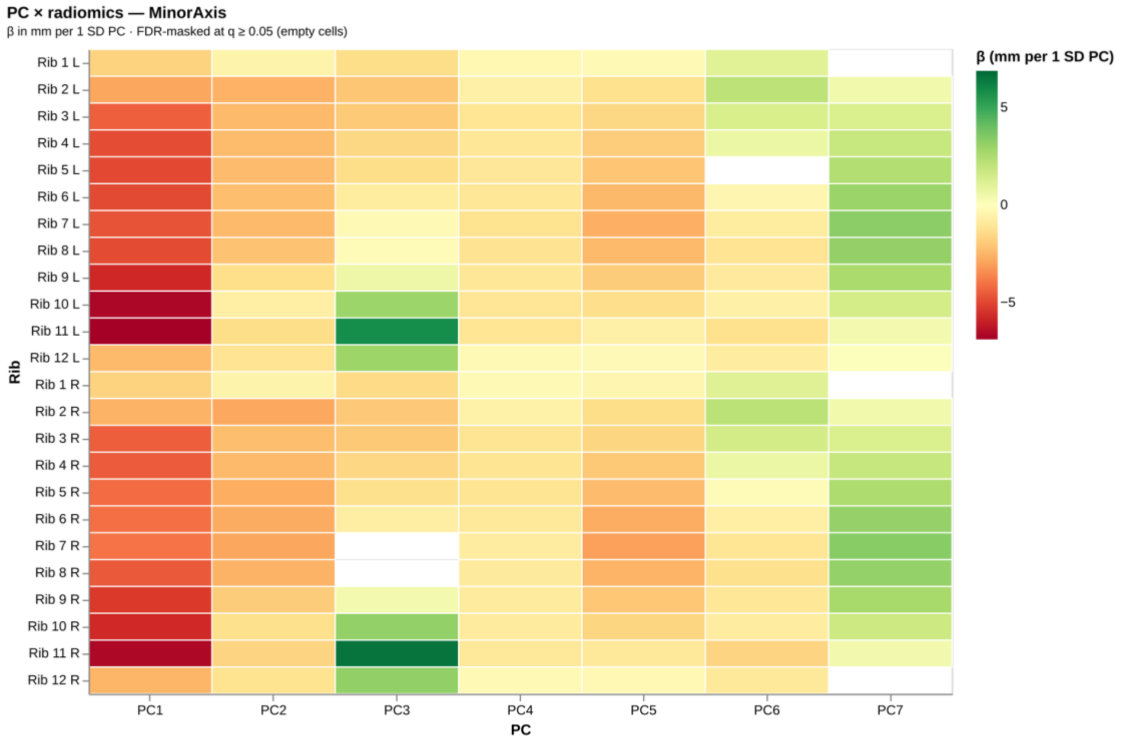


*(j) MinorAxis*

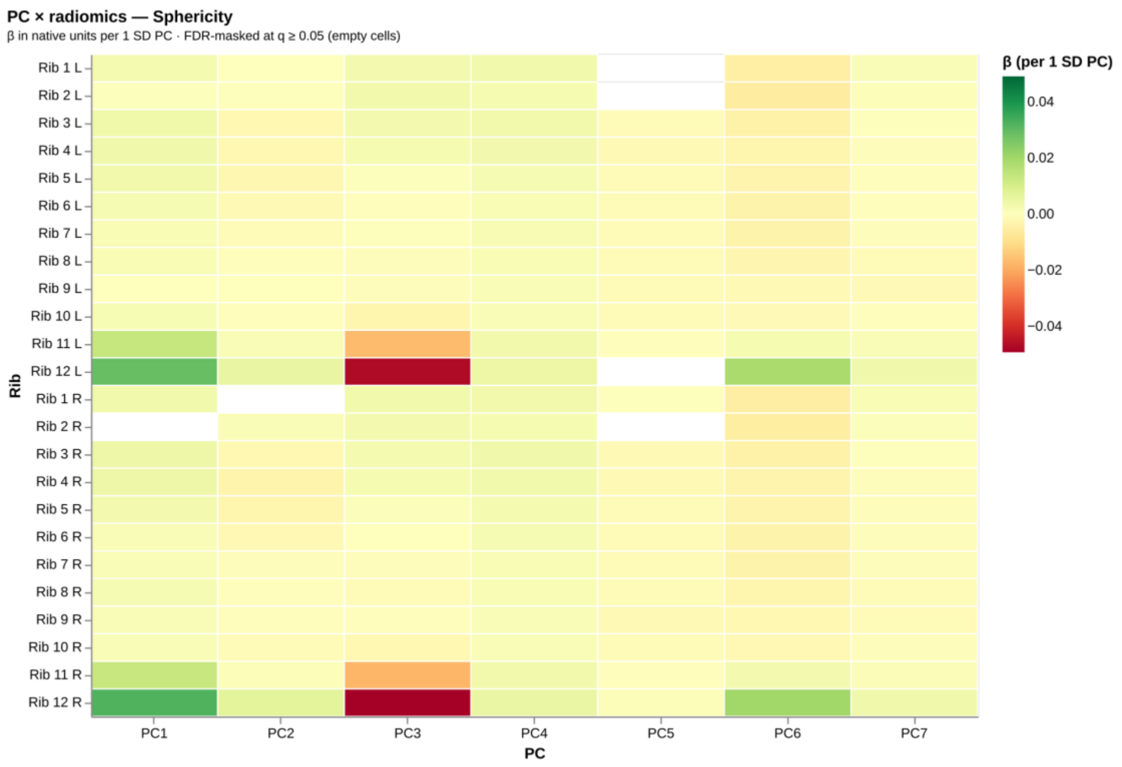


*(k) Sphericity*

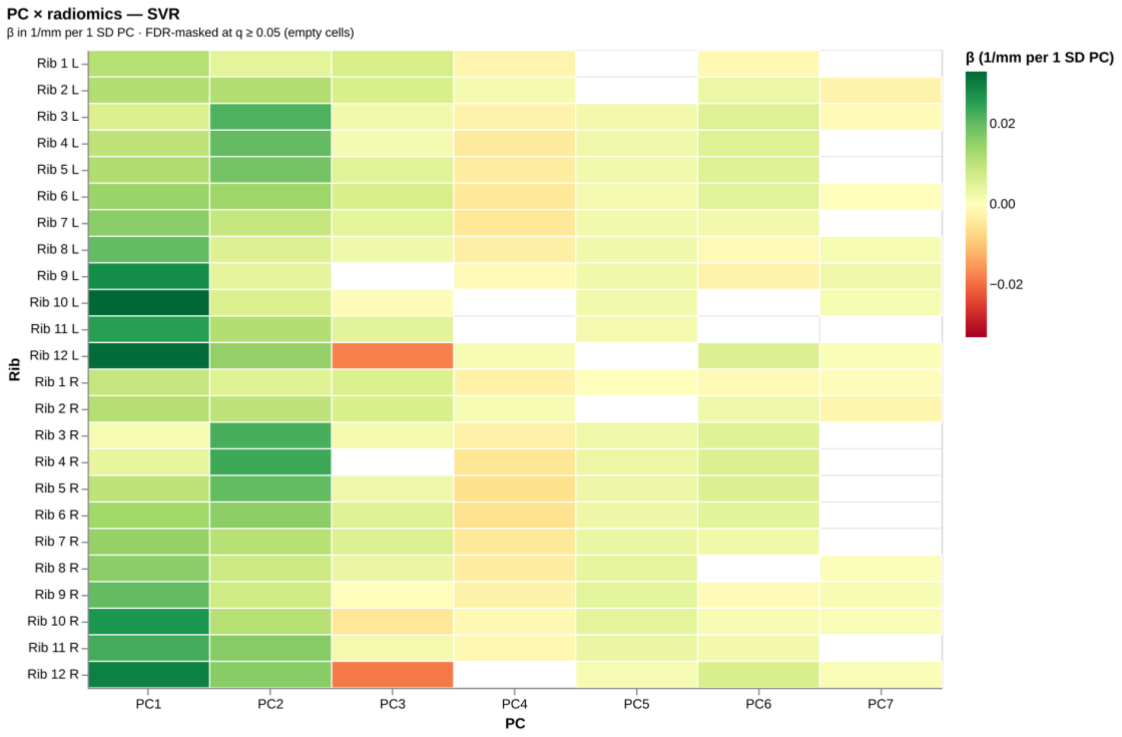

*(l) SVR*

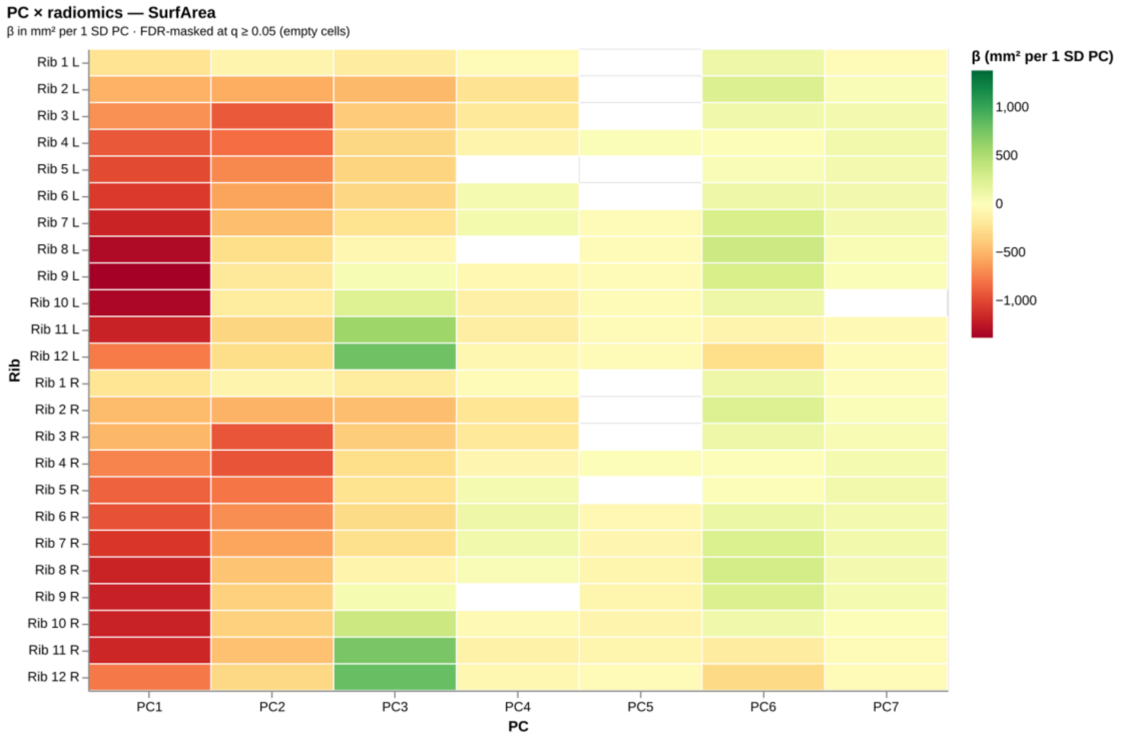


*(m) SurfArea*

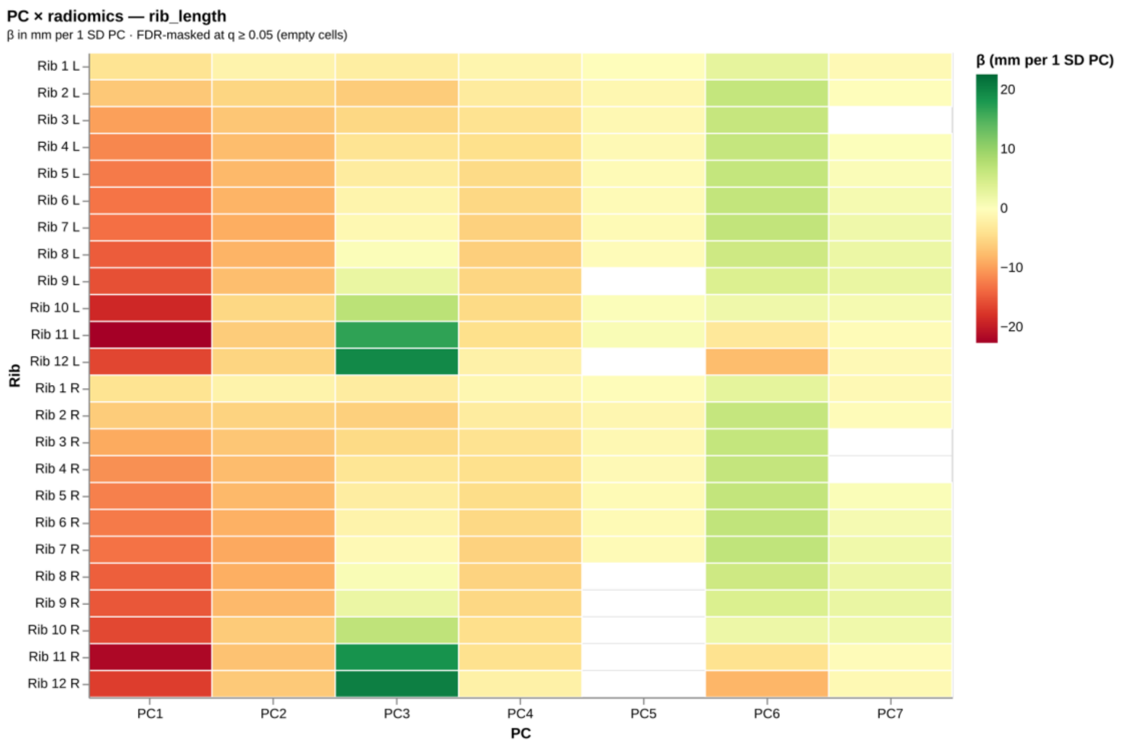


*(n) rib_length*

*Supplementary Figure 22:* ***Per-descriptor anatomical cross-walk.*** *Standardised slope of one shape descriptor at every rib position on each PC score, one panel per descriptor. Print shows all 14 descriptors; the interactive supplement browses the same set.*
